# Beyond BMI: an interpretable integrated body composition index from low-dose chest CT for all-cause mortality risk stratification: a multicentre study

**DOI:** 10.64898/2026.08.05.26359437

**Authors:** Jirong Yi, Krishna K. Patel, Robert J.H. Miller, Anna M. Marcinkiewicz, Assiata Kamagate, Aakash Shanbhag, Waseem Hijazi, Mark Lemley, Jianhang Zhou, Joanna X. Liang, Giselle Ramirez, Shiva Mostafavi, Meghana Urs, Clemens P. Spielvogel, Leandro Slipczuk, Mark Travin, Erick Alexanderson, Isabel Caraval-Juarez, René R. S. Packard, Mouaz Al-Mallah, Terrence D. Ruddy, Andrew J. Einstein, Attila Feher, Edward J. Miller, Wanda Acampa, Stacey Knight, Viet T Le, Steve Mason, Vinicius F. Calsavara, Panithaya Chareonthaitawee, Samuel Wopperer, Alan C. Kwan, Lixia Wang, Debiao Li, Elliot K. Fishman, Felipe Lopez-Ramirez, Daniel S. Berman, Jacek Kwiecinski, Damini Dey, Marcelo Di Carli, Piotr J. Slomka

## Abstract

**Background:** Body composition is recognized as a major determinant of health outcomes, but its multidimensional nature makes clinical adoption challenging. We sought to develop and validate a body composition index (BCI) for all-cause mortality risk assessment, integrating variables of six body composition tissues.

**Methods:** We analyzed 28509 consecutive patients undergoing myocardial perfusion imaging with routine low-dose chest CT attenuation correction (CTAC) scans acquired during myocardial perfusion imaging (MPI) at 12 centers across four countries. An artificial intelligence-based BCI was developed in a cohort of 15037 patients’ CTACs by integrating the CT-derived metrics of bone, skeletal muscle, and four adipose tissue compartments, coronary artery calcium score, and basic demographic variables (age, sex, BMI). The performance of BCI for mortality prediction was validated in an internal cohort of 6444 patients and an external cohort of 7028 patients by prognosis, calibration, net benefit, and explainability. Model-based simulation of tissue metrics modification was performed to evaluate estimated mortality risk reduction.

**Findings:** During a median of 3.5 (IQR [1.9, 5.1]) years, 4697 (16%) patients died. In the external testing cohort, the BCI demonstrated excellent discrimination for mortality (area under receiver operating characteristic curve 0.78 (95% CI [0.76, 0.79]) and Harrell’s concordance index 0.75 [0.73, 0.76]), calibration, and net benefit overall and across pre-specified subgroups stratified by patient characteristics and imaging protocols. Visceral adipose tissue attenuation was the most influential body composition measure, followed by skeletal muscle volume. Simulated improvement in body composition was associated with significant mortality risk reduction.

**Interpretation:** An index combining six body composition measures obtained opportunistically from routine chest CT provides robust mortality risk stratification. By converting complex body composition information into a single interpretable score, the BCI can facilitate clinical implementation of opportunistic CT biomarkers and guide individualized preventive strategies.

**Funding:** National Institute of Health

**Research in context:** *Evidence before this study:* We searched PubMed and Google Scholar on July 7, 2026 for English language studies using the terms (“mortality prediction” OR “death prediction”) AND (“integrated body composition analysis” OR “body composition metrics integration”) AND (“predictive model” OR “model development”), and identified 41 publications describing predictive models incorporating body composition metrics. Recent advances in artificial intelligence (AI) have enabled increasingly granular quantification of body composition from CT, with multiple tissues and tissue-specific metrics showing independent prognostic value for all-cause mortality (ACM) prediction. However, reported associations of body composition metrics with mortality risk were highly inconsistent or even contradictory across studies. This heterogeneity likely reflected the differences in study design or the inherent complexity of body composition analysis, such as the interaction among variables derived from multiple tissues. Thus, it is difficult to interpret body composition results reliably in clinical practice. We hypothesized that integrating multiple body composition metrics could achieve comprehensive and reliable prediction of mortality across patient populations.

*Added value of this study:* In this large international, multicenter longitudinal cohort study of 28509 consecutive patients with suspected or known coronary artery disease (CAD) undergoing myocardial perfusion imaging across 12 sites in four countries, we developed and externally validated an artificial intelligence-based body composition index (BCI) for all-cause mortality prediction. The BCI integrated chest CT-derived metrics of six body composition tissue compartments – bone, skeletal muscle (SM), subcutaneous adipose tissue (SAT), intramuscular adipose tissue (IMAT), visceral adipose tissue (VAT), and epicardial adipose tissue (EAT), together with demographics (age, sex, BMI) and CT-derived coronary artery calcium (CAC) score, all derived automatically from routine low-dose attenuation correction CT scans without additional imaging or laboratory tests. In an external validation cohort of 7028 patients, the BCI achieved robust discrimination for mortality prediction, substantially outperforming a baseline model using demographics and calcium score alone, and exceeded the discriminative performance of prior body composition models. Performance was consistent across subgroups defined by sex, age, body mass index, cardiometabolic risk factors, imaging modality, and acquisition protocol, supporting generalizability across clinical settings. The BCI demonstrated excellent calibration and net clinical benefit. Explainability analysis identified visceral adipose tissue attenuation and skeletal muscle volume as the dominant contributors to mortality risk, offering tissue-specific therapeutic targets that complement generic weight-based metrics. Model-based simulations suggested that improvements in these specific compartments could reduce estimated mortality risk more effectively than equivalent changes in BMI alone, pointing toward precision therapeutic targets beyond weight reduction. Our study improved over previous literatures on body composition-based predictive models in one or more of the following ways: (1) we considered six types of body composition tissues while existing studies considered only three or less tissues, thus significantly higher granularity and improved ability to capture refined tissue interactions; (2) our model was rigorously validated in external sites, showing cross-site generalizability while most previous work lacked external validation; (3) our model achieved significant prediction performance improvement over previous body composition predictive models, offering better clinical outcomes prediction and facilitating the suitability for widespread clinical adoption; (4) our study was conducted with a large multi-center cohort across multiple countries, achieving statistical power and demonstrating reliability towards patients characteristics and imaging acquisition protocols while previous studies involved small single-center cohorts; (5) our model required only CT scans and basic demographics which were widely available in clinical routine while the existing models required dedicated laboratory tests or specialized imaging modalities, enhancing the applicability in routine clinical practice.

*Implications of all the available evidence:* BCI offers an explainable and clinically deployable tool for improved prediction of mortality risk from existing chest CT scans. Its use could inform updates to existing risk models, guide precision preventive strategies, and support development of therapeutics targeting specific body composition phenotypes.

## Introduction

Opportunistic body composition analysis from existing CT scans is an emerging approach for improving all-cause mortality (ACM) risk stratification, as artificial intelligence (AI) has made automated quantification of CT-derived body composition increasingly feasible at scale.^1–4^ Multiple tissue-specific measures from skeletal muscle, bone, and adipose tissue compartments, have demonstrated independent prognostic value beyond established cardiometabolic risk factors.^5–8^ Yet, translating these findings into reliable clinical tools has proven difficult. Reported associations between individual body composition metrics and mortality have been highly inconsistent across studies, and in some cases directionally opposite, reflecting the inherent contextual dependence of these metrics on patient characteristics.^5–8^ The multidimensional nature of body composition data compounds this difficulty as no single measure captures the full complexity of tissue distribution,^1, 3, 9, 10^ and optimally combining multiple metrics is challenging during routine clinical interpretation.

Existing body composition prediction models have several limitations that have hindered clinical translation. They considered three or fewer body composition tissues, thus with limited granularity and less effective in capturing the multidimensional interactions.^4, 11, 12^ Most were developed in small or single-center cohorts without external validation, limiting confidence in their generalizability across clinical settings.^4, 11, 12^ Multiple models also relied on dedicated biomarkers or laboratory tests not routinely available from CT, reducing their practicality.^13–15^ They achieved modest prediction performance, leaving room for substantial improvement.^13–15^

To address these gaps, we developed and externally validated an AI-based body composition index (BCI) for all-cause mortality (ACM) risk stratification. The BCI was derived from routine low-dose attenuation correction CT scans acquired during myocardial perfusion imaging and integrates quantitative CT-derived metrics from six tissue compartments, coronary artery calcium (CAC), and basic demographic parameters (sex, age, and BMI) into a single clinically interpretable index. We hypothesized that this integration would achieve improved and more reliable prediction of mortality risk.

## Methods

### Study population

This retrospective study considered 31170 consecutive patients with suspected or known coronary artery disease (CAD) who underwent single photon emission computed tomography/CT (SPECT/CT) or positron emission tomography/CT (PET/CT) myocardial perfusion imaging (MPI) between 2007 and 2022 across 12 centers from four countries participating in the REgistry of Fast Myocardial Perfusion Imaging with NExt Generation SPECT (REFINE SPECT) or the REgistry of Flow and Perfusion Imaging for Artificial Intelligence with PET (REFINE PET) (Supplementary Figure 1; Supplementary Methods).^16, 17^ Following previous studies, we excluded patients who lacked CT attenuation correction (CTAC) scans, or did not have coverage of the T5–T11 vertebral levels on CTAC, with 28509 patients finally included for analysis.^1, 18^ Incomplete clinical variables were imputed due to the small missing rate, such as 0.2% for body mass index (BMI). The study protocol adhered to the Declaration of Helsinki and was approved by the institutional review boards at each participating center, with overall study granted by the IRB at Cedars-Sinai Medical Center (Los Angeles, California).

### CTAC imaging acquisition parameters

All CTAC scans were low-dose, non-contrast, and not electrocardiographically gated. The scans were acquired with different hybrid scanners from multiple vendors (General Electric, Philips, and Siemens). The range of slice thickness, tube current, and tube voltage were 2.0-5.0 mm, 10-595 mAs, and 80-140 kVp, respectively (Supplementary Table 1).

### CTAC imaging measures

Body composition tissues were automatically segmented and quantified from a sub-volume between vertebrae T5 and T11 of CTAC scans using a previously validated approach.^1, 18^ This study considered three types of volumetric metrics of six tissues (subcutaneous adipose tissue (SAT), skeletal muscle (SM), intramuscular adipose tissue (IMAT), bone, epicardial adipose tissue (EAT), and visceral adipose tissue (VAT)) including attenuation (mean of CT attenuation value, in Hounsfield unit (HU)), heterogeneity (standard deviation (SD) of CT attenuation value, in HU), volume index (volume divided by height squared, in cm^3^/m^2^). The ratio of SM volume to total adipose tissue (TAT) volume (sum of SAT, IMAT, EAT, and VAT) and the ratio of ectopic adipose tissue volume (IMAT, EAT, and VAT) to TAT volume were also used.^19^ Coronary artery calcium (CAC) score was derived from CTAC by a previously validated model.^20^

### Clinical outcome

Baseline demographic and medical information was obtained from the registries (Supplementary Methods).^16, 17^ MPI and medical history variables were used for adjustment in risk stratification evaluation in external testing cohort, including imaging modality (SPECT/CT vs PET/CT), left ventricle ejection fraction (LVEF), stress total perfusion deficit (TPD), hypertension, diabetes mellitus, dyslipidemia, family history of CAD, and smoking status. The primary endpoint of this study was ACM (Supplementary Methods).

### BCI model development and external testing scheme

We developed a support vector machine (SVM)-based BCI model for integrating body composition metrics, basic demographics (sex ,age, BMI) and CT-derived CAC score using 15037 patients from ten sites by formulating a supervised binary classification task for all-cause mortality event prediction (Supplementary Methods).^21^ Model performance was rigorously validated using an external testing cohort consisting of 7028 cases from two additional sites and an internal cohort of 6444 cases (Supplementary Figure 2). The search space for key hyperparameters and the final selected hyperparameters were determined from model development data (Supplementary Table 2). We followed similar protocols to develop different models tailored to various clinical scenarios. Explainability of BCI model was investigated using the SHapley Additive exPlanations (SHAP) values.^21^

### Simulated body composition improvement and mortality risk reduction

To illustrate the potential effect of targeted changes in body composition metrics on predicted mortality risk reduction, we conducted a model-based simulation using the BCI. For each body composition metric, we fixed all other model variables at their mean values in the external validation cohort and varied the selected metric incrementally across its observed physiological range. The corresponding BCI was calculated at each step. An analogous simulation was performed for BMI to compare the estimated reduction in predicted mortality risk associated with changes in individual body composition metrics versus BMI. The analyses were repeated within subgroups stratified by patient characteristics.

### Statistical analysis

Continuous variables were summarized as medians with interquartile ranges (IQR: Q1–Q3), and categorical variables were presented as frequencies with percentages. Comparisons of continuous variables were conducted using the Kruskal-Wallis rank sum test. For categorical variables, Pearson’s chi-squared test or Fisher’s exact test was applied.

The primary evaluation metrics were area under receiver operating characteristic (ROC) curve (AUC) for discriminative prediction of death event, and Harrell’s concordance index (C-index) for prognostic prediction of death. We also assessed the robustness and the generalizability of BCI in subpopulations stratified by patient characteristics and imaging acquisition protocols. The net clinical benefit and the calibration performance of BCI model were investigated using the decision curve analysis and the calibration plots. The race-related evaluation of the model in internal testing cohort was presented since the external testing cohort consisted of predominantly White patients with patients from other races < 2%. The DeLong test was used to assess statistical significance of difference in AUC analysis.

Kaplan-Meier curves were used for prognostic evaluation with the log-rank test for assessing the difference between groups. Cox proportional hazard regression models were fitted for ACM with results presented as hazard ratios (HRs) with corresponding 95% confidence intervals (CIs). We used cutoffs corresponding to negative predictive value (NPV) of 98% and positive predictive value (PPV) of 40% determined from development cohort to categorize patients into low-risk, intermediate-risk and high-risk groups for risk stratification by Kaplan-Meier survival analysis. These thresholds were selected to reliably identify low-risk and high-risk individuals, consistent with established approaches in biomarker-based risk stratification.^22^ The linearity and the proportionality assumptions of Cox model were verified by visually inspecting the Martingale residual and the Schoenfeld residual plots, respectively. Youden index-based cutoff was used for prediction pattern analysis. All statistical tests were two-sided, with a significance level of 5%, i.e., p-value <0.05 considered statistically significant. Analyses were conducted using R version 4.3.2 (RStudio). An overview of our study design is shown in Figure 1.

**Figure 1:**
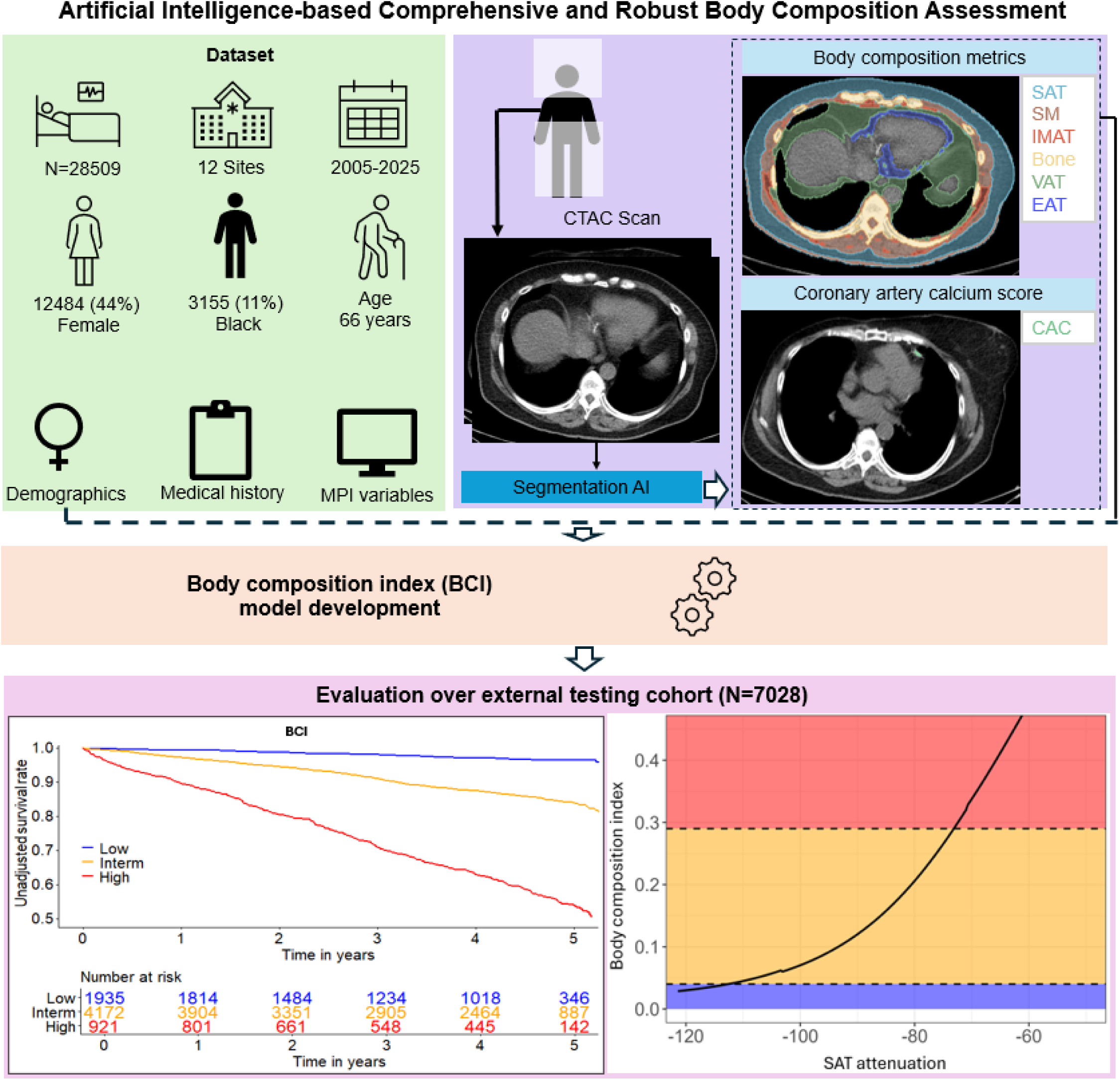
Overview of study design. CAC – coronary artery calcium, MPI – myocardial perfusion imaging, CTAC – computed tomography attenuation correction, AI – artificial intelligence, SAT – subcutaneous adipose tissue, SM – skeletal muscle, IMAT – intramuscular adipose tissue, VAT – visceral adipose tissue, EAT – epicardial adipose tissue.

## Results

### Study population

The final cohort consisted of 28509 patients with median age 66 [58, 74] years and 12484 (44%) females (Table 1). During the follow up of 3.54 [1.92, 5.11] years, 4697 (16%) patients died. In external testing cohort (N=7028), median age was 66 [57, 74] years with 3069 (44%) females and median follow up of 4.36 [2.30, 4.89] years, during which 949 (14%) patients died. Patients who died were older (73 [64, 81] vs 65 [57, 72]) and had higher VAT attenuation (-77 [-83, -70] vs -82 [-86, -77]), both p<0.05 (Supplementary Table 3). Similar patterns were observed in the development and internal testing cohorts (Supplementary Table 3-4).

**Table 1:**
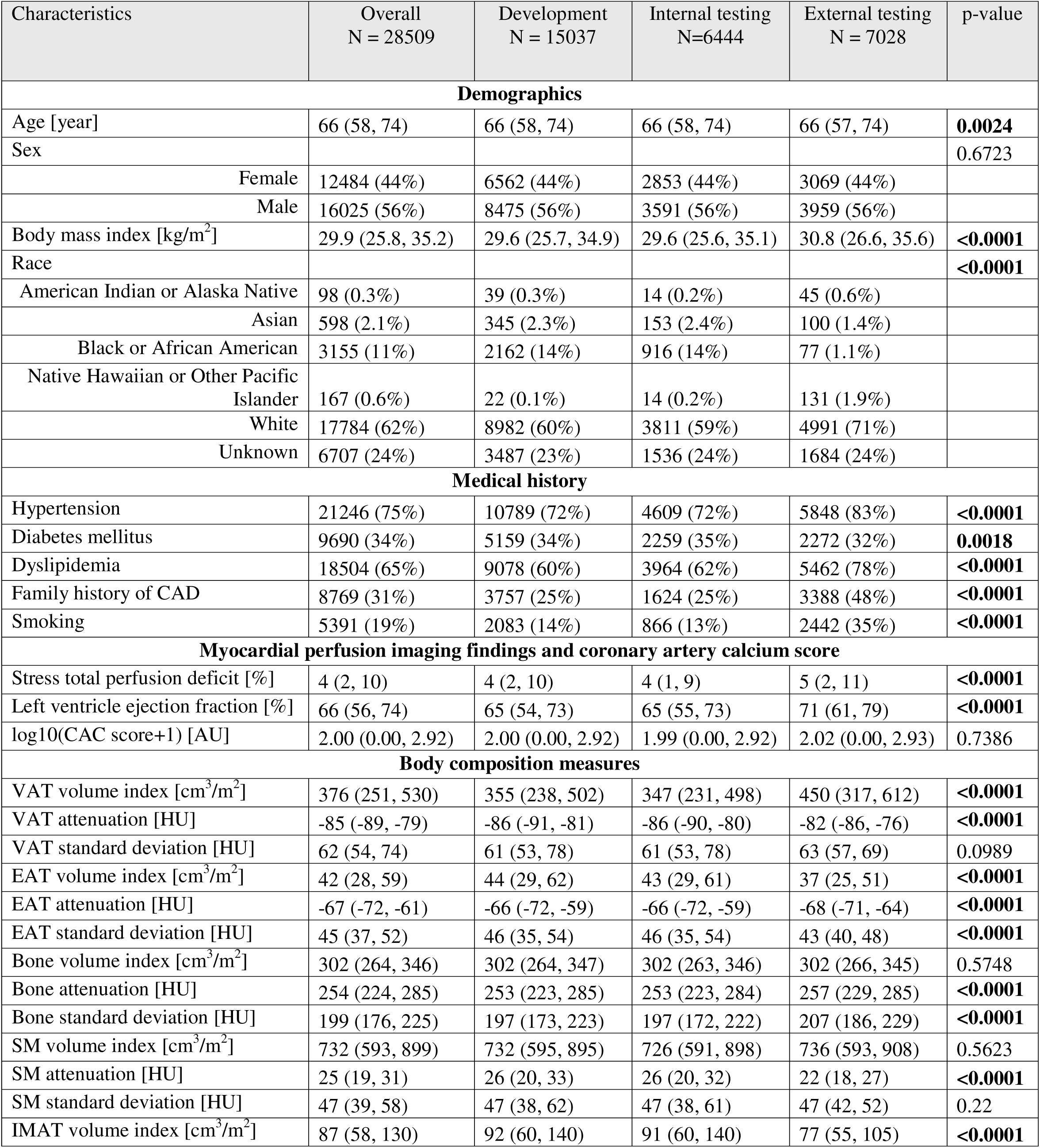
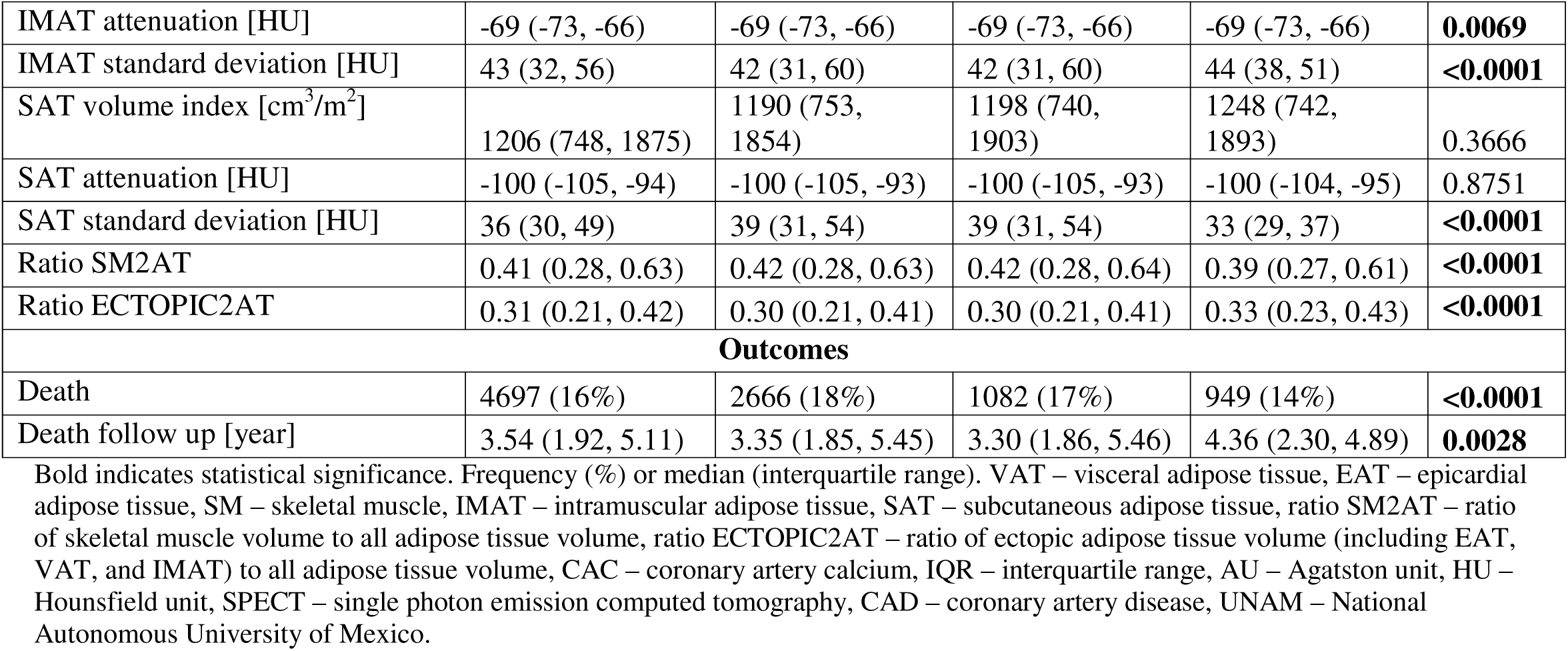
Baseline characteristics of the development, internal and external testing cohorts.

| Characteristics | Overall<br>N = 28509 | Development<br>N = 15037 | Internal testing<br>N=6444 | External testing<br>N = 7028 | p-value |
| --- | --- | --- | --- | --- | --- |
| **Demographics** |  |  |  |  |  |
| Age [year] | 66 (58, 74) | 66 (58, 74) | 66 (58, 74) | 66 (57, 74) | **0.0024** |
| Sex |  |  |  |  | 0.6723 |
| Female | 12484 (44%) | 6562 (44%) | 2853 (44%) | 3069 (44%) |  |
| Male | 16025 (56%) | 8475 (56%) | 3591 (56%) | 3959 (56%) |  |
| Body mass index [kg/m<sup>2</sup>] | 29.9 (25.8, 35.2) | 29.6 (25.7, 34.9) | 29.6 (25.6, 35.1) | 30.8 (26.6, 35.6) | **<0.0001** |
| Race |  |  |  |  | **<0.0001** |
| American Indian or Alaska Native | 98 (0.3%) | 39 (0.3%) | 14 (0.2%) | 45 (0.6%) |  |
| Asian | 598 (2.1%) | 345 (2.3%) | 153 (2.4%) | 100 (1.4%) |  |
| Black or African American | 3155 (11%) | 2162 (14%) | 916 (14%) | 77 (1.1%) |  |
| Native Hawaiian or Other Pacific Islander | 167 (0.6%) | 22 (0.1%) | 14 (0.2%) | 131 (1.9%) |  |
| White | 17784 (62%) | 8982 (60%) | 3811 (59%) | 4991 (71%) |  |
| Unknown | 6707 (24%) | 3487 (23%) | 1536 (24%) | 1684 (24%) |  |
| **Medical history** |  |  |  |  |  |
| Hypertension | 21246 (75%) | 10789 (72%) | 4609 (72%) | 5848 (83%) | **<0.0001** |
| Diabetes mellitus | 9690 (34%) | 5159 (34%) | 2259 (35%) | 2272 (32%) | **0.0018** |
| Dyslipidemia | 18504 (65%) | 9078 (60%) | 3964 (62%) | 5462 (78%) | **<0.0001** |
| Family history of CAD | 8769 (31%) | 3757 (25%) | 1624 (25%) | 3388 (48%) | **<0.0001** |
| Smoking | 5391 (19%) | 2083 (14%) | 866 (13%) | 2442 (35%) | **<0.0001** |
| **Myocardial perfusion imaging findings and coronary artery calcium score** |  |  |  |  |  |
| Stress total perfusion deficit [%] | 4 (2, 10) | 4 (2, 10) | 4 (1, 9) | 5 (2, 11) | **<0.0001** |
| Left ventricle ejection fraction [%] | 66 (56, 74) | 65 (54, 73) | 65 (55, 73) | 71 (61, 79) | **<0.0001** |
| log10(CAC score+1) [AU] | 2.00 (0.00, 2.92) | 2.00 (0.00, 2.92) | 1.99 (0.00, 2.92) | 2.02 (0.00, 2.93) | 0.7386 |
| **Body composition measures** |  |  |  |  |  |
| VAT volume index [cm<sup>3</sup>/m<sup>2</sup>] | 376 (251, 530) | 355 (238, 502) | 347 (231, 498) | 450 (317, 612) | **<0.0001** |
| VAT attenuation [HU] | -85 (-89, -79) | -86 (-91, -81) | -86 (-90, -80) | -82 (-86, -76) | **<0.0001** |
| VAT standard deviation [HU] | 62 (54, 74) | 61 (53, 78) | 61 (53, 78) | 63 (57, 69) | 0.0989 |
| EAT volume index [cm<sup>3</sup>/m<sup>2</sup>] | 42 (28, 59) | 44 (29, 62) | 43 (29, 61) | 37 (25, 51) | **<0.0001** |
| EAT attenuation [HU] | -67 (-72, -61) | -66 (-72, -59) | -66 (-72, -59) | -68 (-71, -64) | **<0.0001** |
| EAT standard deviation [HU] | 45 (37, 52) | 46 (35, 54) | 46 (35, 54) | 43 (40, 48) | **<0.0001** |
| Bone volume index [cm<sup>3</sup>/m<sup>2</sup>] | 302 (264, 346) | 302 (264, 347) | 302 (263, 346) | 302 (266, 345) | 0.5748 |
| Bone attenuation [HU] | 254 (224, 285) | 253 (223, 285) | 253 (223, 284) | 257 (229, 285) | **<0.0001** |
| Bone standard deviation [HU] | 199 (176, 225) | 197 (173, 223) | 197 (172, 222) | 207 (186, 229) | **<0.0001** |
| SM volume index [cm<sup>3</sup>/m<sup>2</sup>] | 732 (593, 899) | 732 (595, 895) | 726 (591, 898) | 736 (593, 908) | 0.5623 |
| SM attenuation [HU] | 25 (19, 31) | 26 (20, 33) | 26 (20, 32) | 22 (18, 27) | **<0.0001** |
| SM standard deviation [HU] | 47 (39, 58) | 47 (38, 62) | 47 (38, 61) | 47 (42, 52) | 0.22 |
| IMAT volume index [cm<sup>3</sup>/m<sup>2</sup>] | 87 (58, 130) | 92 (60, 140) | 91 (60, 140) | 77 (55, 105) | **<0.0001** |
| IMAT attenuation [HU] | -69 (-73, -66) | -69 (-73, -66) | -69 (-73, -66) | -69 (-73, -66) | **0.0069** |
| IMAT standard deviation [HU] | 43 (32, 56) | 42 (31, 60) | 42 (31, 60) | 44 (38, 51) | **<0.0001** |
| SAT volume index [cm<sup>3</sup>/m<sup>2</sup>] | 1206 (748, 1875) | 1190 (753, 1854) | 1198 (740, 1903) | 1248 (742, 1893) | 0.3666 |
| SAT attenuation [HU] | -100 (-105, -94) | -100 (-105, -93) | -100 (-105, -93) | -100 (-104, -95) | 0.8751 |
| SAT standard deviation [HU] | 36 (30, 49) | 39 (31, 54) | 39 (31, 54) | 33 (29, 37) | **<0.0001** |
| Ratio SM2AT | 0.41 (0.28, 0.63) | 0.42 (0.28, 0.63) | 0.42 (0.28, 0.64) | 0.39 (0.27, 0.61) | **<0.0001** |
| Ratio ECTOPIC2AT | 0.31 (0.21, 0.42) | 0.30 (0.21, 0.41) | 0.30 (0.21, 0.41) | 0.33 (0.23, 0.43) | **<0.0001** |
| **Outcomes** |  |  |  |  |  |
| Death | 4697 (16%) | 2666 (18%) | 1082 (17%) | 949 (14%) | **<0.0001** |
| Death follow up [year] | 3.54 (1.92, 5.11) | 3.35 (1.85, 5.45) | 3.30 (1.86, 5.46) | 4.36 (2.30, 4.89) | **0.0028** |

Bold indicates statistical significance. Frequency (%) or median (interquartile range). VAT – visceral adipose tissue, EAT – epicardial adipose tissue, SM – skeletal muscle, IMAT – intramuscular adipose tissue, SAT – subcutaneous adipose tissue, ratio SM2AT – ratio of skeletal muscle volume to all adipose tissue volume, ratio ECTOPIC2AT – ratio of ectopic adipose tissue volume (including EAT, VAT, and IMAT) to all adipose tissue volume, CAC – coronary artery calcium, IQR – interquartile range, AU – Agatston unit, HU – Hounsfield unit, SPECT – single photon emission computed tomography, CAD – coronary artery disease, UNAM – National Autonomous University of Mexico.

### Mortality prediction

The integrated BCI achieved excellent prediction performance for all-cause mortality (AUC 0.78 [0.76, 0.79]; Figure 2 and Supplementary Table 5). The discriminative prediction was reliable across time windows (Figure 2) and across patient subgroups (Table 2; Supplementary Table 6). Mortality risk increased monotonically with BCI decile, with the highest decile carrying a 20-fold higher risk relative to the lowest (unadjusted HR 20.0 [12.1, 33.0], p<0.05; Figure 3). Higher BCI was reliably associated with increased mortality risk overall (unadjusted HR 1.9 and adjusted HR 1.73 for per SD increase of BCI, p<0.05) and in patient subgroups (Supplementary Table 7). BCI achieved similar performance in patient subgroups (Supplementary Table 7; Supplementary Figure 3).

**Figure 2:**
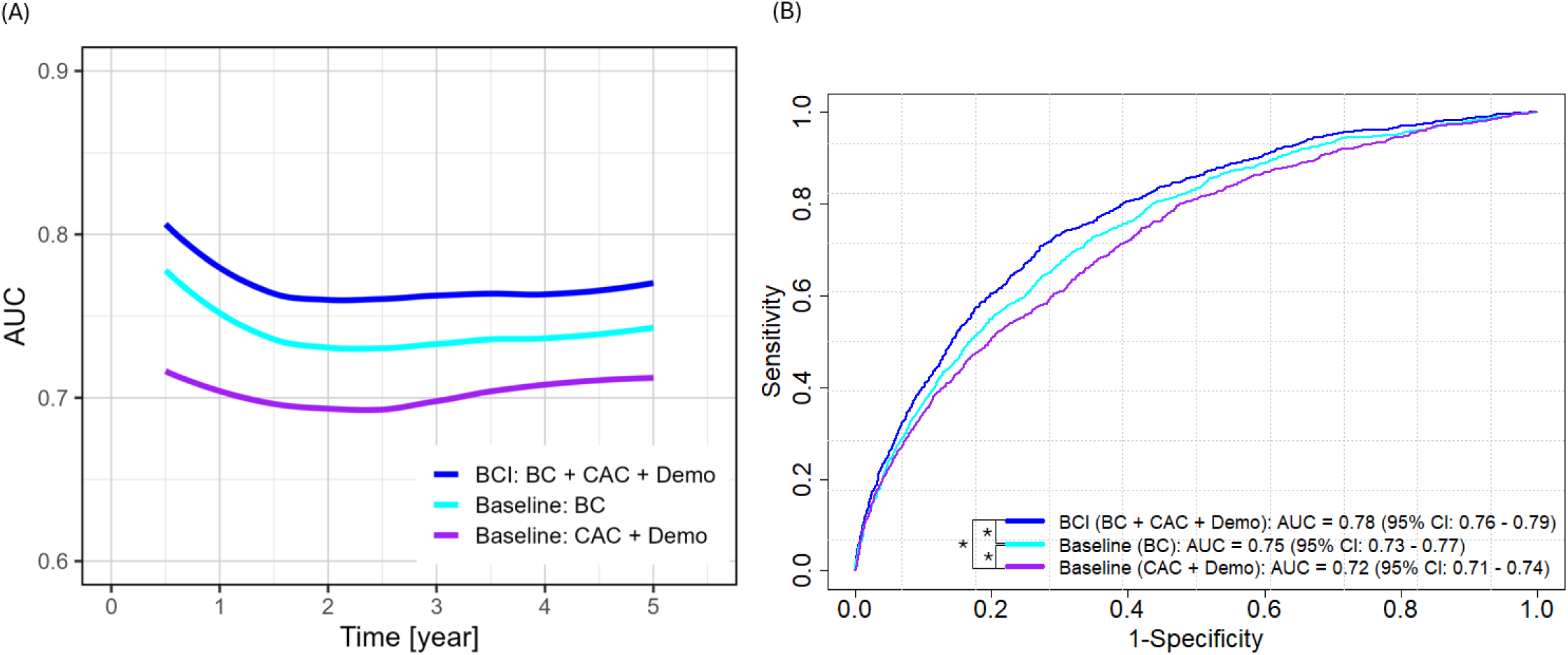
Prediction performance of body composition index (BCI) in external testing cohort. (A): time-dependent area under receiver operating characteristics curve (AUC) for mortality event prediction; (B): AUC for mortality event prediction during entire follow up time. The baseline models (cyan and purple) used only CT-derived body composition (BC) metrics or only basic demographic (Demo) variables (sex, age, and BMI) and CT-derived coronary artery calcium (CAC) score. The final model **BCI (blue)** integrated BC metrics, CAC score, and Demo. * p<0.05.

**Figure 3.**
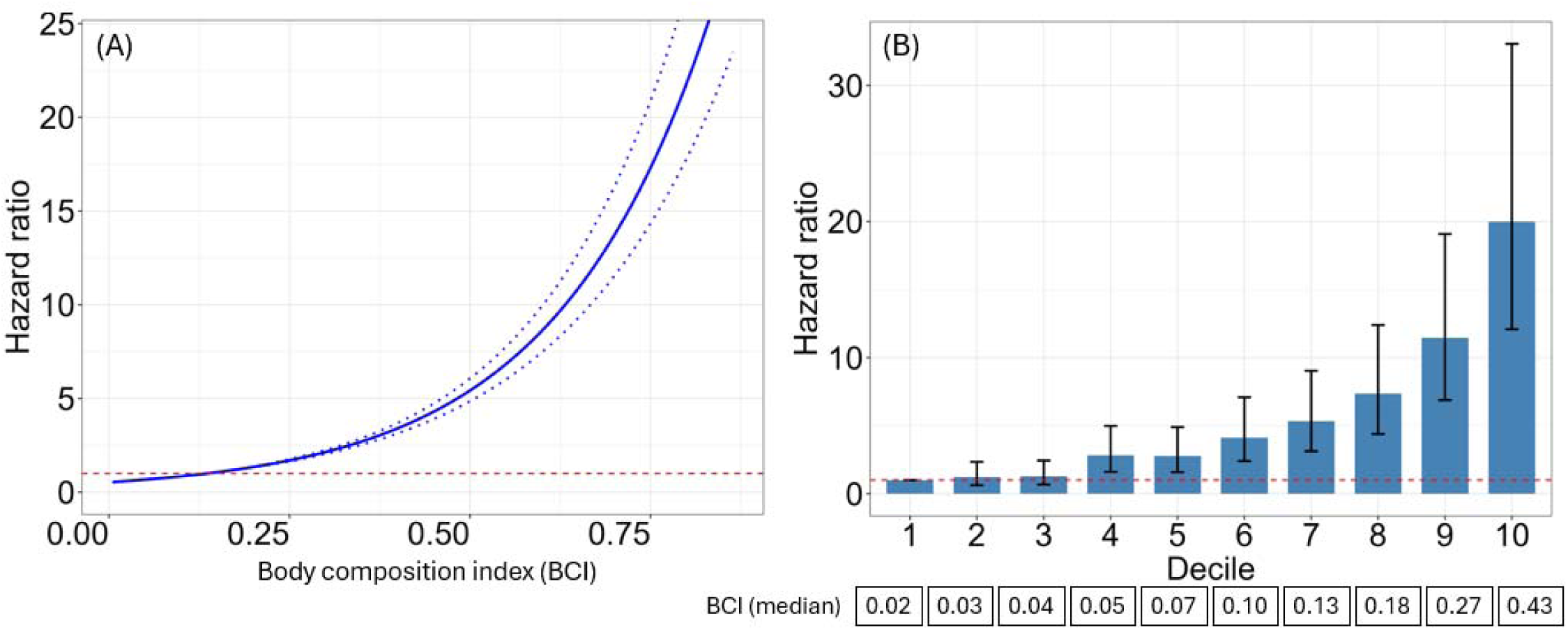
Relation between unadjusted hazard ratio and body composition index (BCI) in external testing cohort. (A): the population mean was used as reference for calculating hazard ratio; (B): the first decile was used as reference for calculating hazard ratio. The dashed lines in (A) and the error bars in (B) indicated 95% confidence interval. Red dashed lines corresponded to hazard ratio 1. BCI integrated body composition metrics, coronary artery calcium (CAC) score, and demographics. Red dashed horizontal line corresponds to hazard ratio 1. CAC – coronary artery calcium score, MPI – three myocardial perfusion variables.

**Table 2.**
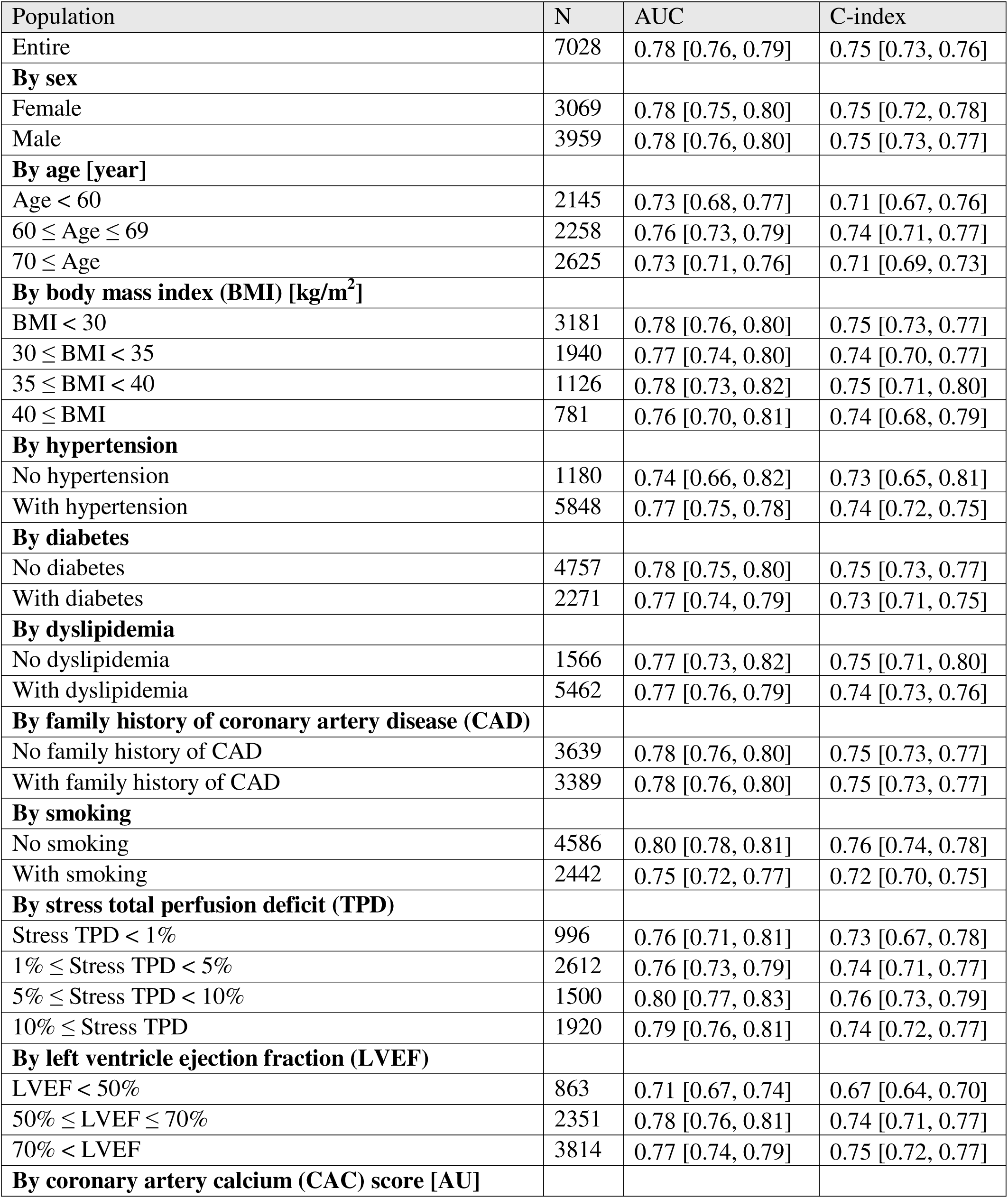
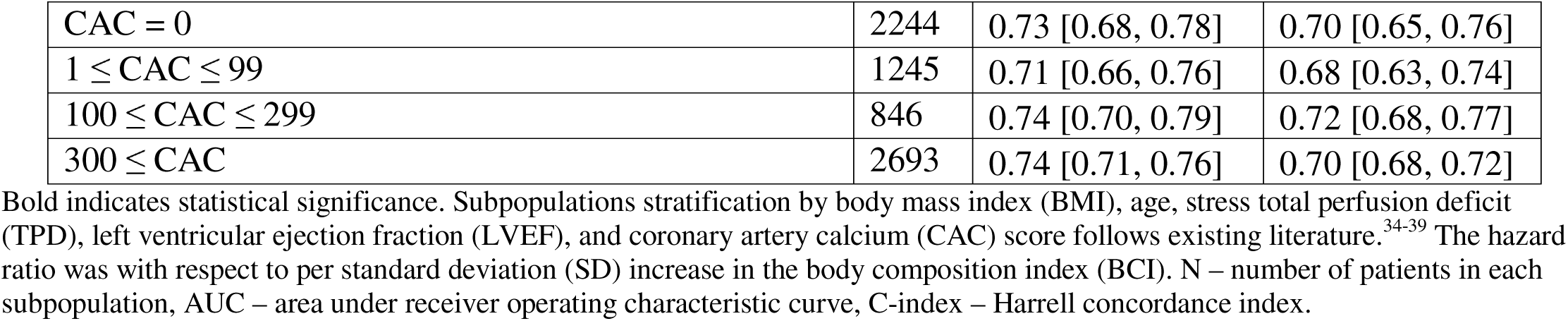
Reliable discriminative prediction performance of the body composition index (BCI), integrating CT-derived body composition metrics, coronary artery calcium score, and demographic variables, in the external testing cohort overall and across subgroups defined by patient characteristics and imaging acquisition protocol.

When integrating body composition metrics alone (adipose tissues, bone, skeletal muscle), the model showed reliably strong predictive value in external cohort (unadjusted HR 1.79 [1.71, 1.86], AUC 0.75 [0.73, 0.77], and C-index 0.72 [0.71, 0.74]) as well as its subgroups stratified by patient characteristics and imaging acquisition protocols (Supplementary Tables 5 and 7).

### Model evaluation

#### Explainability

Adipose tissue attenuation and SM volume index were strong contributors to BCI with higher adipose tissue attenuation elevating mortality risk while higher SM volume index reducing risk (Figure 4). Attenuation was the most influential metric from VAT, SAT, EAT, and bone, while volume index was the strongest predictor from IMAT and SM.

**Figure 4:**
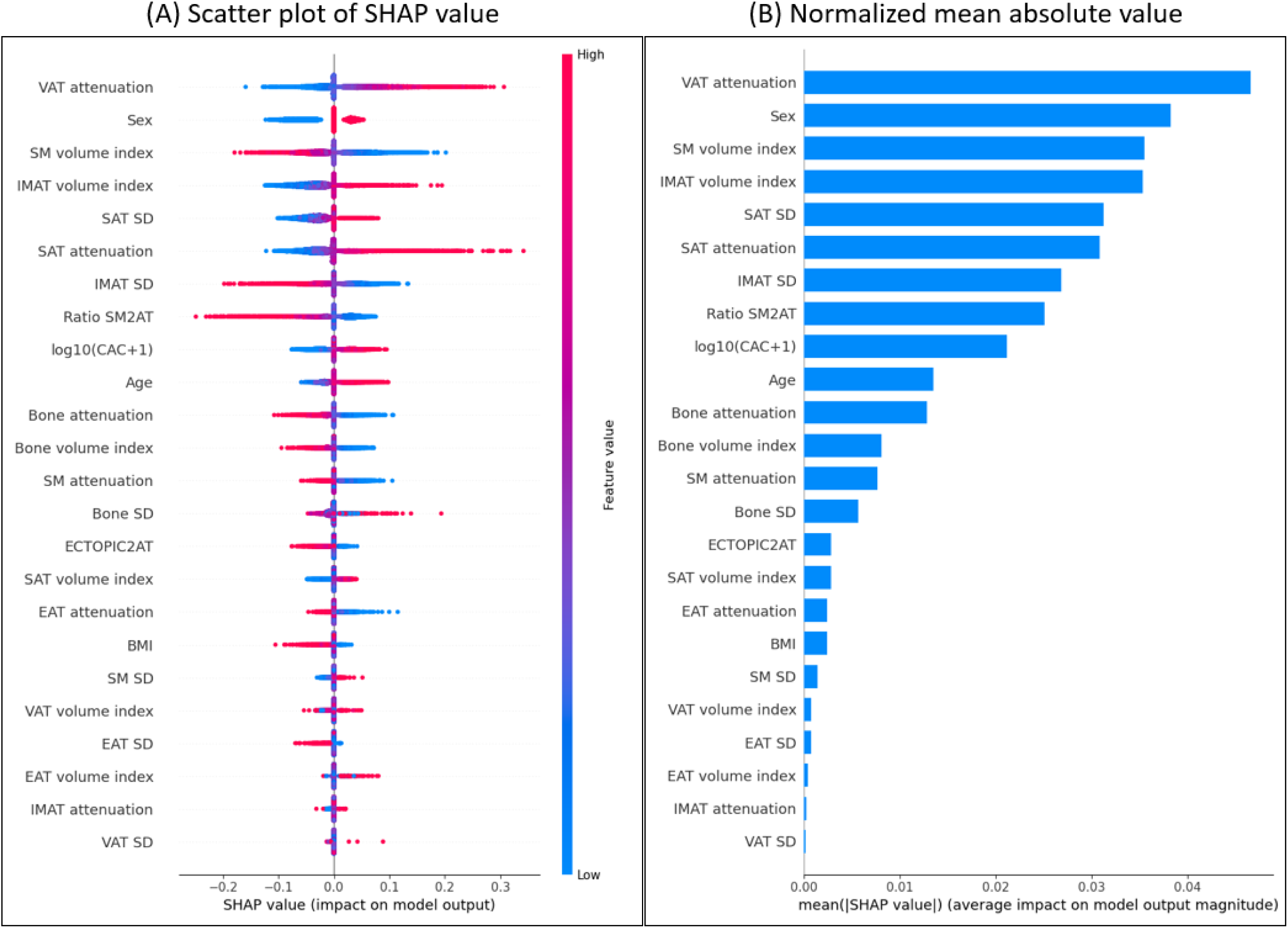
Feature importance of body composition index in the external testing cohort. (A): Beeswarm of SHAP value; (B): bar plot of SHAP value. Body composition index integrated body composition metrics, coronary artery calcium score, and demographics. SHAP – SHapley Additive exPlanations. VAT – visceral adipose tissue, SM – skeletal muscle, IMAT – intramuscular adipose tissue, SAT – subcutaneous adipose tissue, SD – standard deviation, SM2AT – ratio of skeletal muscle volume to all adipose tissue volume, CAC – coronary artery calcium, ECTOPIC2AT – ratio of ectopic adipose tissue volume (including EAT, VAT, and IMAT) to all adipose tissue volume, EAT – epicardial adipose tissue, BMI – body mass index.

When predictive value-based cutoffs were applied, high-risk patients by BCI had significantly higher VAT attenuation (-74 [-80, -68] in high-risk vs -86 [-89, -82] in low-risk patients, p<0.05) (Supplementary Table 8). VAT attenuation and SM volume index remained strong mortality risk predictors in these risk groups (Supplementary Figure 4).

#### Calibration

The BCI model was better calibrated than the baseline model which integrated demographics and CAC score only (calibration slopes 0.92 vs 1.39 and intercepts 0.0 vs -0.29; Supplementary Figure 5).

#### Prediction patterns

When the Youden index threshold is applied, individuals misclassified by the BCI as false positives nonetheless demonstrated clinically meaningful risk profiles. Specifically, false positives had body composition measures and ages comparable to true positives, including high VAT attenuation and advanced age, both of which are associated with increased mortality risk (Supplementary Table 9).

### Potential for clinical deployment

#### Risk stratification pathway

When the BCI was categorized into low-risk, intermediate-risk, and high-risk groups with predictive value-based thresholds, high BCI was associated with a 17-fold elevated mortality risk compared to the low-risk group (HR 17.11, p<0.05; Table 3; Figure 5) in the external testing cohort. The association was reliable across sex (HRs 19.4 in females vs 16.11 in males, all p<0.05) and other pre-specified subgroups (Table 3; Supplementary Table 10; Supplementary Figure 6). BCI identified 1935 (28%) patients as low-risk and 921 (13%) patients as high-risk in the external testing cohort (Figure 6), with similar distributions and predictive values in different subpopulations (Supplementary Figure 7; Supplementary Table 11).

**Figure 5:**
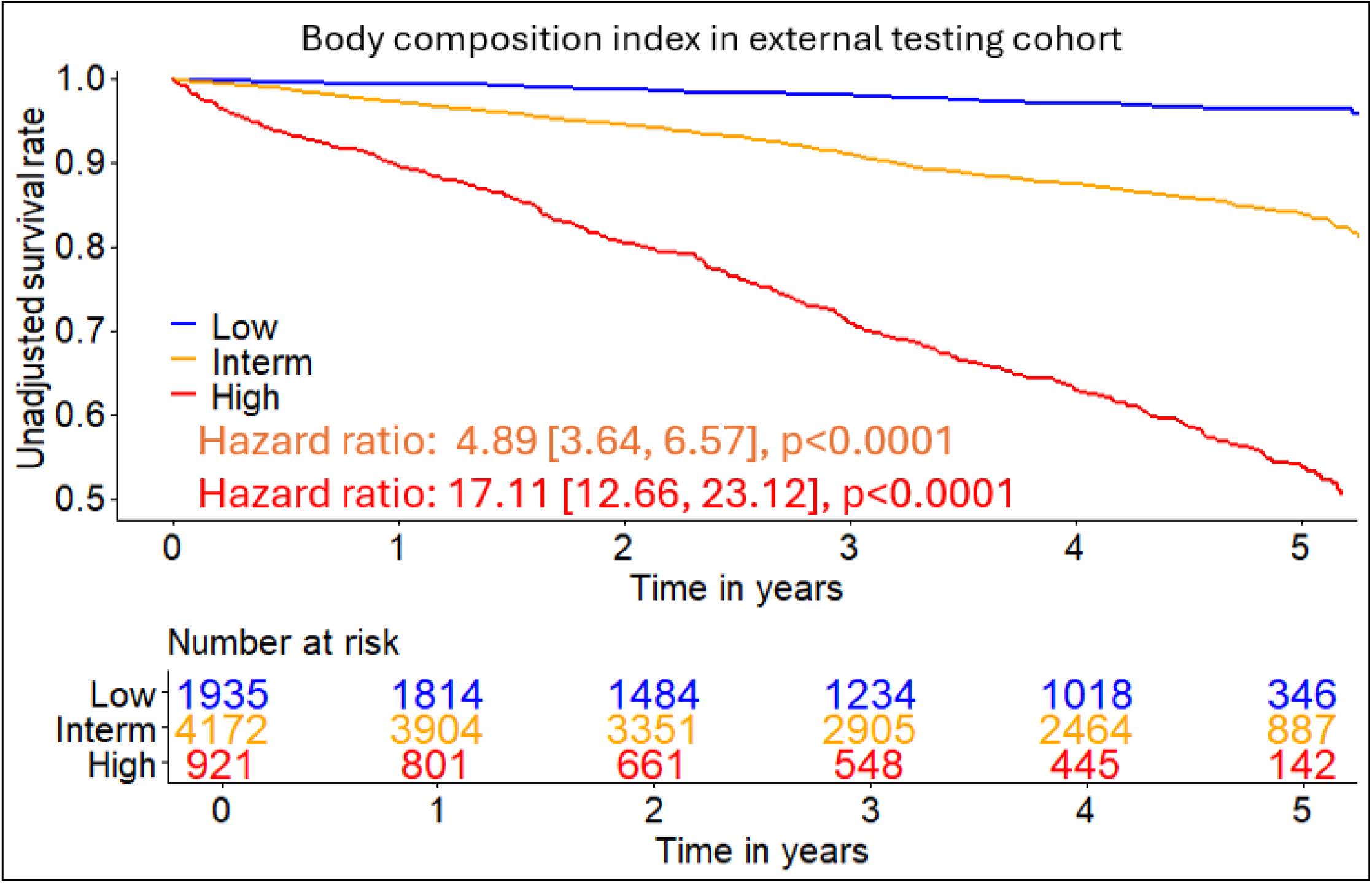
**Kaplan-Meier curve of body composition index for all-cause mortality risk stratification in the external testing cohort**. Body composition index integrated body composition metrics, coronary artery calcium score, and demographics.

**Figure 6:**
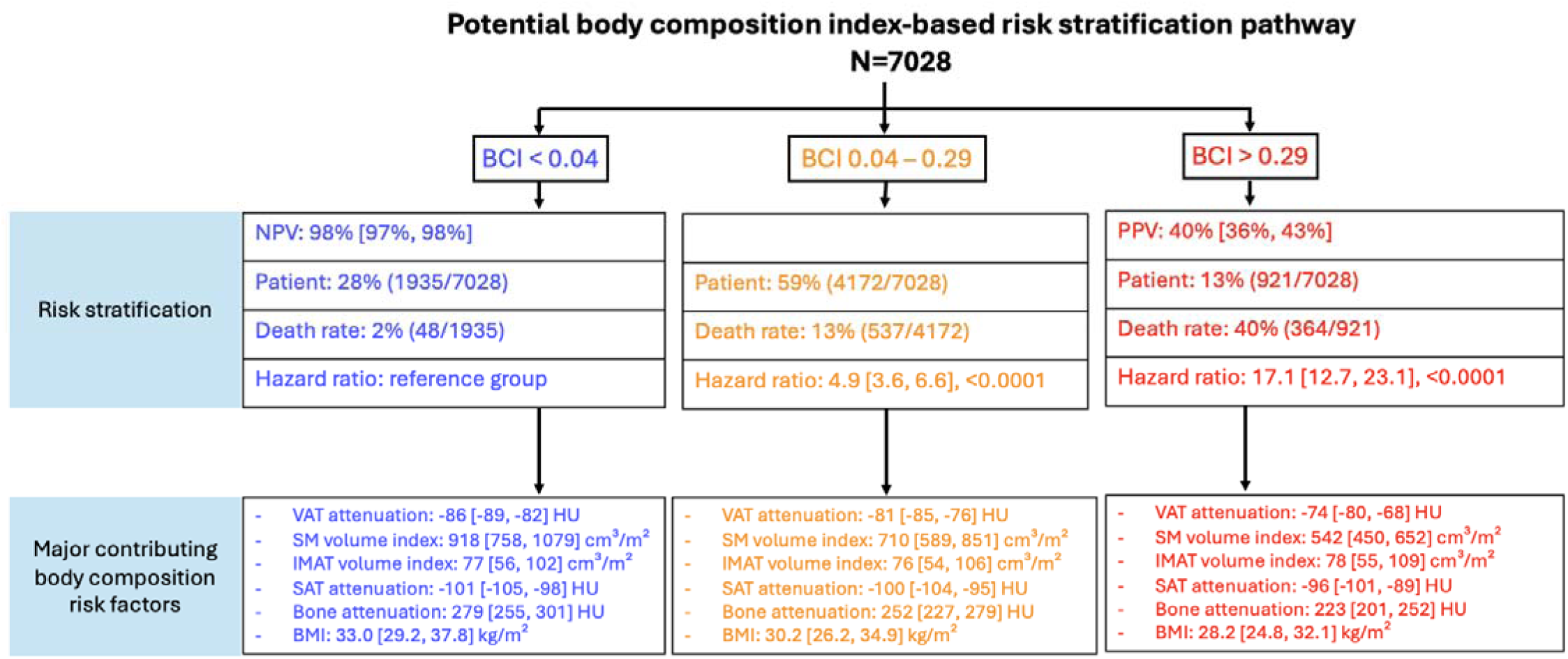
Proposed body composition index-based clinical risk stratification pathway. Body composition index integrated body composition metrics, coronary artery calcium score, and demographics. BCI – body composition index, NPV – negative predictive value, PPV – positive predictive value, VAT – visceral adipose tissue, SM – skeletal muscle, IMAT – intramuscular adipose tissue, SAT – subcutaneous adipose tissue, BMI – body mass index, HU – Hounsfield unit.

**Table 3:**
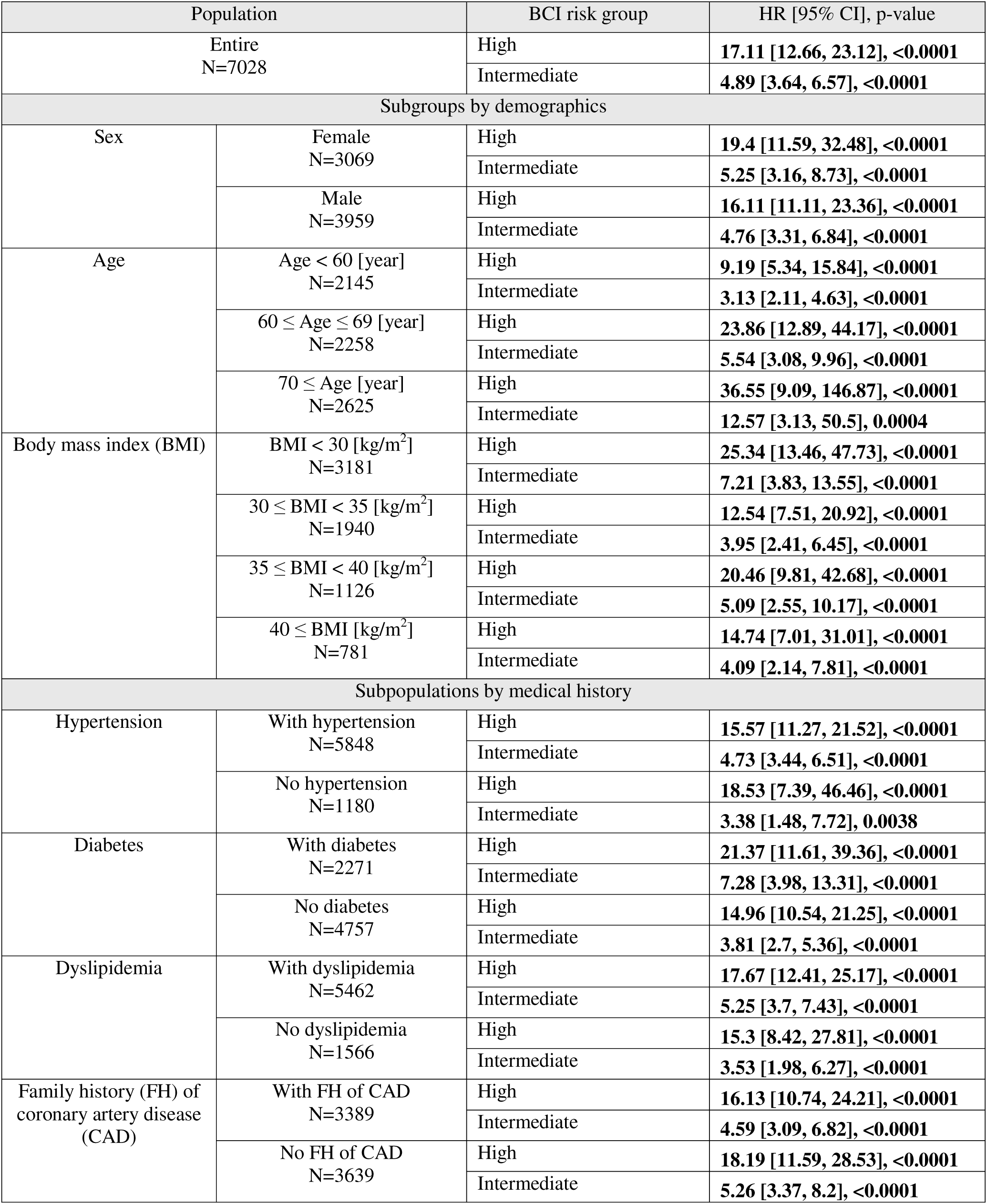
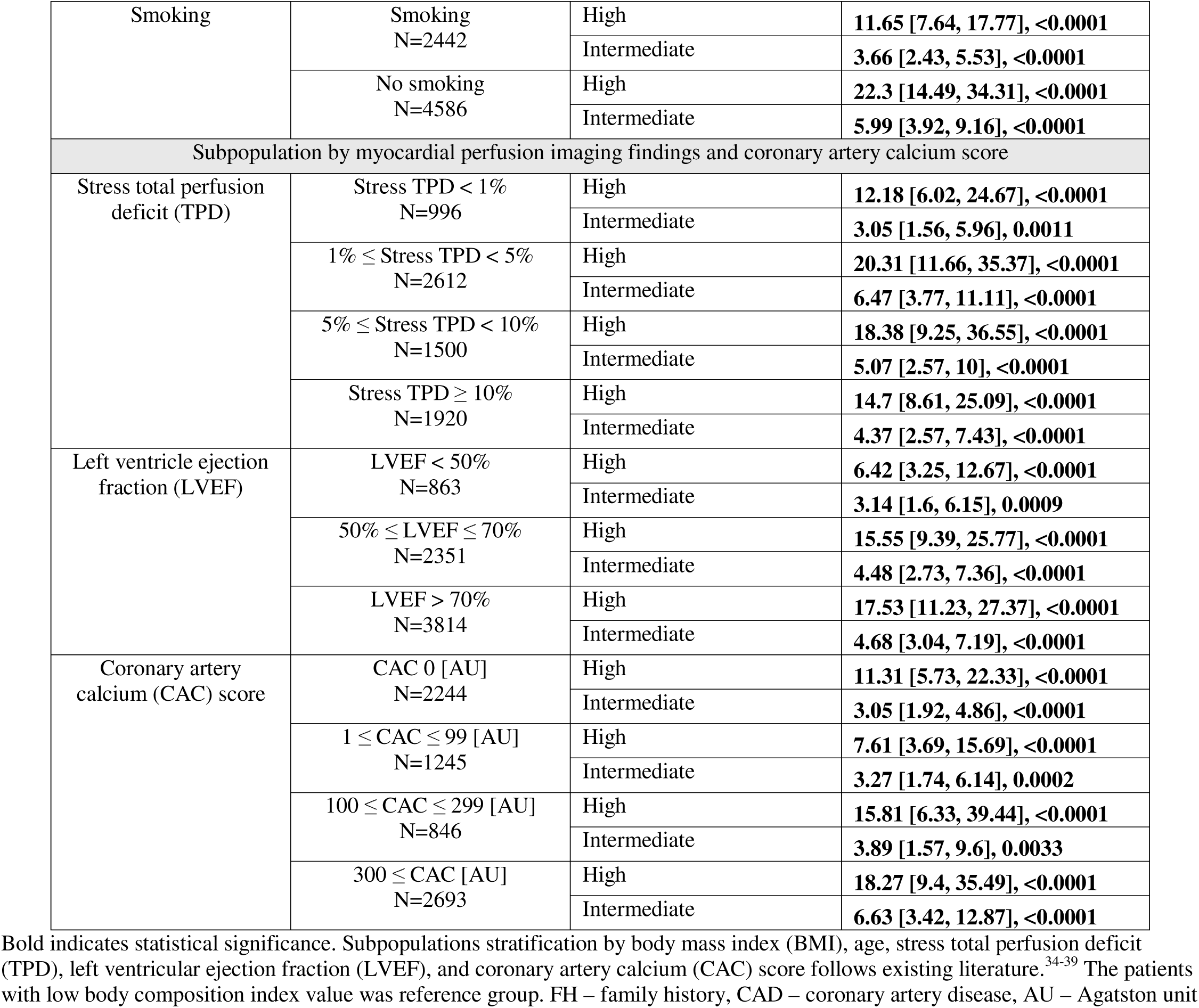
Hazard ratio (HR) for all-cause mortality prediction by body composition index (BCI) risk category in the overall and pre-specified subgroups within the external testing cohort. BCI integrates CT-derived body composition metrics, coronary artery calcium score, and demographic variables.

#### Net clinical benefit

The BCI demonstrated superior net clinical benefit compared with the baseline model (demographics + CAC score) across a full range of decision thresholds in external testing cohort (Supplementary Figure 8), with reliable net benefits observed across patient subgroups (Supplementary Figure 3).

#### Simulation of body composition improvement

Increase in SM volume index and reduction in SAT attenuation were associated with substantial decreases in estimated BCI and mortality risk (Figure 7). A simulated reduction in SAT attenuation from -60 HU by 100% to -120 was associated with a reduction in BCI from high-risk range (> 0.29) to the low-risk range (< 0.04) while a simulated increase in BMI from 25 kg/m^2^ by 100% to 50 kg/m^2^ achieved almost no reduction in BCI from 0.09 to 0.07 (Figure 8). Similar results were also observed in patient subgroups (Supplementary Figures 9-11). Two illustrative patient cases with distinct body composition profiles had dramatically different outcomes (Figure 8).

**Figure 7:**
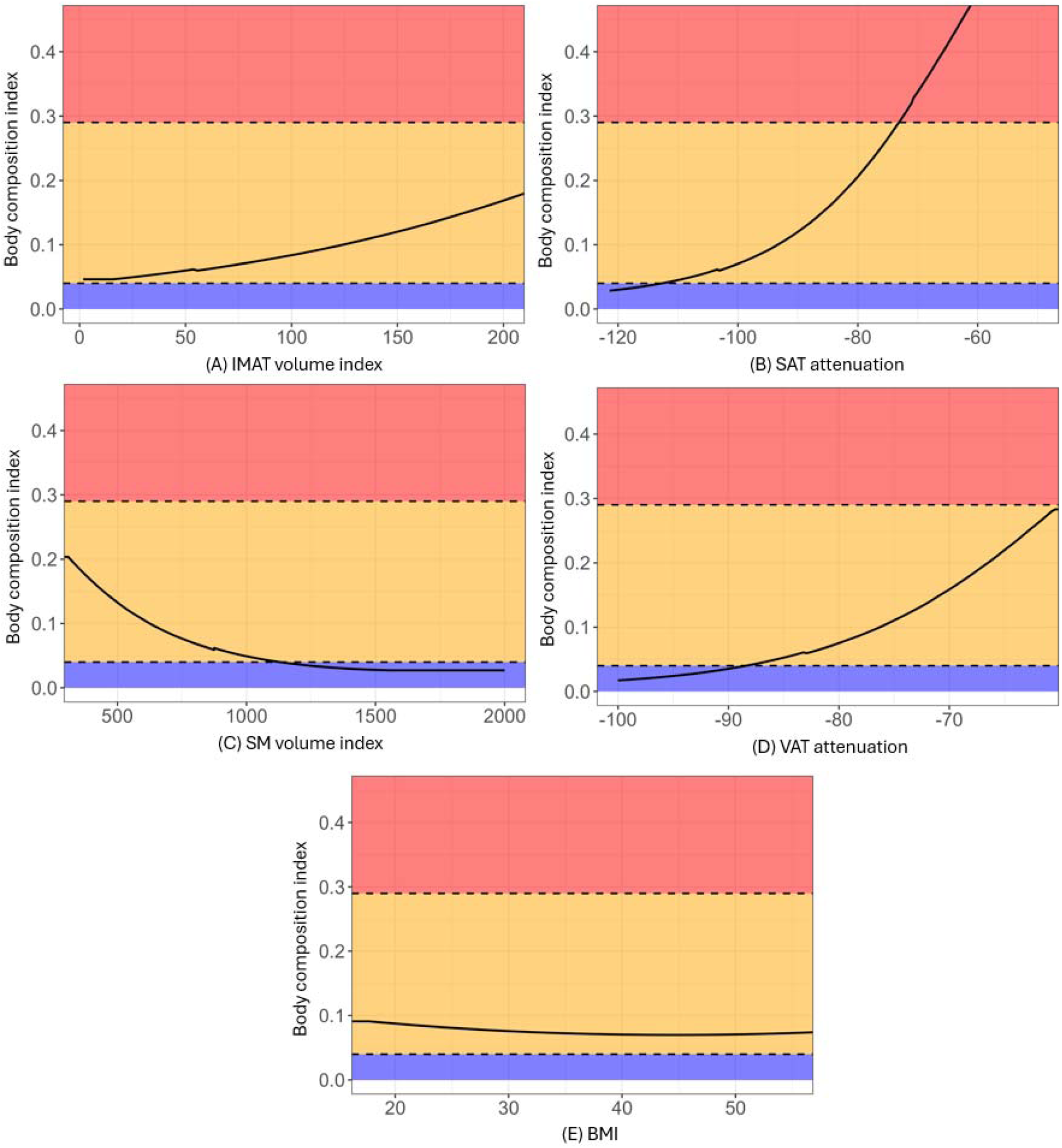
Model-based simulation of estimated body composition index (BCI) reduction through targeted improvement in body composition metric. BCI decreased significantly when body composition metric changed (A-D) but only slightly when BMI changed (E). Red, yellow, and blue regions correspond to high-risk, intermediate-risk, and high-risk groups, respectively. Body composition index (BCI) was estimated as a function of simulated modification in body composition metric. All other body composition metrics were held at subgroup means. Two horizontal dashed lines indicate BCI thresholds of 0.04 and 0.29, corresponding to the low-risk (BCI <0.04), intermediate-risk (0.04 ≤ BCI ≤ 0.29), and high-risk (BCI >0.29) categories. This is a model-based simulation; results should be interpreted as hypothesis-generating and do not imply causal therapeutic efficacy. IMAT – intramuscular adipose tissue, SAT – subcutaneous adipose tissue, SM – skeletal muscle, VAT – visceral adipose tissue, BMI – body mass index, HU – Hounsfield unit.

**Figure 8:**
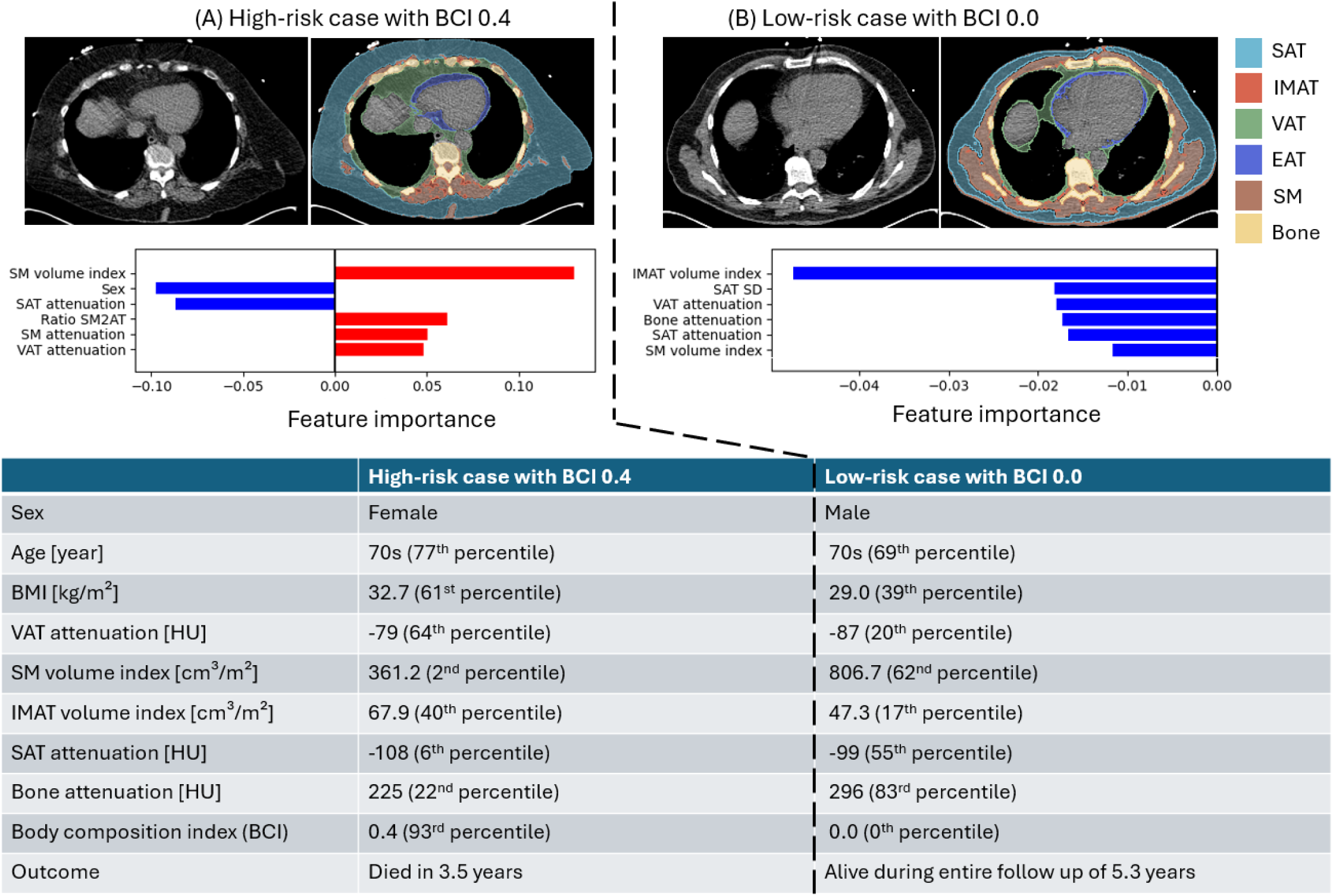
Comparison of patient cases demonstrating high-risk and low-risk body composition index (BCI) profiles. Top panels: CT slice and body composition segmentation; Middle panels: bar plot of SHAP feature importance of top six most significant variables; Bottom panels: body composition metrics. BCI integrated body composition metrics, coronary artery calcium score, and demographics. Red indicated the variable increased the mortality risk while blue indicated the variable decreased the mortality risk. BCI – body composition index, SHAP – SHapley Additive exPlanations. EAT – epicardial adipose tissue, VAT – visceral adipose tissue, SM – skeletal muscle, IMAT – intramuscular adipose tissue, SAT – subcutaneous adipose tissue, ratio SM2AT – ratio of skeletal muscle volume to adipose tissue volume, BMI – body mass index, SD – standard deviation, HU – Hounsfield unit.

#### Race-specific analysis

The prediction performance of BCI was robust to race (AUCs 0.80 [0.76, 0.84] in Black vs 0.82 [0.80, 0.83] in White, p>0.05; unadjusted HRs 1.66 [1.48, 1.88] in Black vs 2.14 [2.01, 2.27] in White, all p<0.05) (Supplementary Figure 12; Supplementary Tables 12-13). When categorized, high BCI was associated with higher mortality risk (HRs 8.08 [3.72, 17.53] in Black vs 10.16 [6.07, 16.98] in White, respectively, all p<0.05) (Supplementary Figure 13).

## Discussion

This study demonstrates that an AI-derived body composition index, developed and externally validated from routine low-dose attenuation correction CT scans, achieves excellent and reliable mortality risk stratification in consecutive patients with suspected or known coronary artery disease. By integrating CT-derived metrics from six tissue compartments with CAC score and demographic variables, the BCI resolves the interpretive complexity and delivers reliable prediction across a broad spectrum of patient profiles. Patients in the high-risk BCI category carried a significantly higher mortality risk than those in the low-risk category. Modifying tissue-specific metrics achieves significantly more risk reduction than generic weight reduction. These results represent a critical step toward making body composition analysis clinically actionable at the point of care, using what is already acquired during routine chest CT.

Prior studies have established that multiple body composition components carry independent prognostic value, but have also highlighted a consistent challenge: associations between individual metrics and mortality are highly variable or even opposite across studies.^5–8, 23^ Traditional non-ML risk stratification approaches had attempted to account for patient-level contextual effects through modifier-specific thresholds or sex-stratified models (for example: sex-stratified prediction models^24–27^). While these approaches represent important progress, they are limited in their ability to capture the multidimensional and interactive nature of body composition data, and have largely been developed in general lower-risk populations without established cardiovascular disease.^1, 3, 10, 23–29^ The AI-based BCI implicitly captures non-linear relationships and interaction effects between tissue metrics and demographic variables, enabling it to handle the multidimensional body composition data. The resulting strong prediction performance in external testing cohort and pre-specified subgroups, substantially exceeds that of prior body composition models,^12, 13^ and complements the existing guideline recommended cardiovascular risk models, which target primary prevention population without known cardiovascular disease.^24–27^

Our explainability analysis identifies adipose tissue attenuation as dominant contributors to BCI, followed by SM volume. Higher VAT attenuation, reflecting denser, less lipid-rich, more metabolically dysfunctional visceral fat could be a marker of fibrosis and inflammation.^30^ Skeletal muscle volume reflects overall musculoskeletal reserve and its independent contribution underscores the prognostic importance of sarcopenia in this population.^9^ Notably, BMI ranked below 14 other body composition metrics in feature importance, consistent with the well-described limitations of BMI as a surrogate for body composition in populations with established disease.^31^

Model-based simulations demonstrated that reducing adipose tissue density was associated with substantially greater estimated BCI reduction than equivalent changes in BMI, particularly in male patients with multiple cardiometabolic comorbidities. Similarly, increasing skeletal muscle volume produced meaningful estimated risk reduction that BMI modification could not replicate. These findings suggest that interventions targeting specific body composition compartments may have greater prognostic relevance than strategies focused solely on body weight. Whether VAT attenuation is a modifiable therapeutic target or primarily a marker of advanced adipose tissue dysfunction warrants further investigation.^32^ However, skeletal muscle mass is readily modifiable through lifestyle interventions, particularly resistance training and other muscle-building activities, supporting its potential as a practical therapeutic target.^33^

Several limitations warrant consideration. The study population was derived from imaging registries focused on cardiometabolic risk, and residual confounders such as cancer or cancer treatment may influence body composition association with mortality, not fully captured by our model.^7^ Our body composition metrics were quantified from chest CT between vertebrae T5 and T11, thus only a standardized but estimation of the whole-body composition. The cutoffs for risk stratification were derived using thresholds corresponding to specific NPV and PPV from development data. Site- and population-specific cutoffs and calibration may offer even better risk stratification. Due to the retrospective nature of this study, additional prospective validation and cohort studies in different populations will be important for validating its generalizability and clinical adoption.

In conclusion, the externally validated BCI offers a clinically deployable, explainable, and scalable tool for mortality risk stratification from existing chest CT scans. Its ability to consolidate multidimensional body composition data into a single interpretable and reliable score improved risk stratification. It also has the potential to inform updates to existing risk models and support the development and monitoring of therapeutics targeting specific body composition phenotypes.

## Contributors

JY co-designed the study, developed the algorithms/models, conducted data processing, experiments, analysis, and co-wrote the manuscript. KP, RM and AM co-wrote the manuscript and contributed materials, clinical expertise, and technical expertise. PJS co-designed the study, provided the overall guidance and study funding, co-wrote the manuscript, and contributed materials, clinical expertise, and technical expertise. AK, AS, WH, ML, JZ, JXL, GR, SM, MKU, CPS, LS, MT, EA, IC, RRSP, MA, TDR, AJE, AF, EJM, WA, SK, VTL, SM, VFC, PC, SW, ACK, LW, DL, EKF, FL, DSB, JK, DD, and MDC contributed materials, clinical expertise, and technical expertise. All authors critically revised the manuscript and contributed to its formation. JY and KP contributed equally as co-first authors. JY, KP, RM, AM, and AS directly accessed and verified the data in the study. All authors had full access to all the data in the study, accept the final responsibility to submit for publication and take responsibility for the contents of the manuscript. JY and PJS were responsible for the decision to submit the manuscript.

## Data sharing

To the extent allowed by data sharing agreements and IRB protocols, the patient data from this manuscript will be shared upon written request to the corresponding author at. All implementations of the proposed body composition index model primarily utilized Python version 3.11.5 and open-source libraries such as scikit-learn (version 1.5.2). All statistical analyses were performed using RStudio 4.3.2, with open-source libraries such as tdROC (version 2.0) (Supplementary Methods). The source statistical analysis codes will be released at https://github.com/qimagingAI and can be accessed from the date of publication. All experiments were conducted on a desktop system running Windows 11 Pro 64-bit, equipped with 128GB RAM, an AMD Ryzen 9 7950X 16-Core Processor, and an NVIDIA RTX 4090 24GB GPU.

## Acknowledgements

This research was supported in part by grant R35HL161195 from the National Heart, Lung, and Blood Institute (NHLBI) and R01EB034586 from the National Institute of Biomedical Imaging and Bioengineering (PI: Piotr Slomka). The content is solely the responsibility of the authors and does not necessarily represent the official views of the National Institutes of Health.

