## Supplemental Material for "Beyond BMI: an interpretable integrated body composition index from low-dose chest CT for all-cause mortality risk stratification: a multicentre study"

**Supplementary appendix**

**Supplementary Methods**

**Data collection, heterogeneity, and harmonization**

For sites participating in the REgistry of Fast myocardial perfusion Imaging with NExt generation SPECT (REFINE SPECT)—including Yale University, Columbia University, University of Calgary, and University of Ottawa—de-identified data from patients with suspected or known coronary artery disease (CAD) who underwent SPECT/CT myocardial perfusion imaging (MPI) with prognostic follow-up were transferred to the core laboratory at Cedars-Sinai Medical Center using Health Insurance Portability and Accountability Act (HIPAA)-compliant methods, including secure Box servers and encrypted physical media.^1, 2^

At each site, patient identifiers (e.g., medical record numbers, names, and imaging dates) were removed using dedicated de-identification software, which generated a cross-referenced list linking anonymized patient codes to the original data. SPECT/CT images for each patient were collected in Digital Imaging and Communications in Medicine (DICOM) format. When multiple scans were available for a given patient, only the earliest scan was included. Image reconstruction was performed using vendor-recommended iterative algorithms applied to list-mode data, optimized for each scanner at the participating sites.^1, 2^

All image data underwent quality control review by experienced core laboratory technologists. Myocardial perfusion quantification was performed using the Quantitative Perfusion SPECT (QPS) software, which generated metrics including stress total perfusion deficit (TPD) and left ventricular ejection fraction (LVEF).^1-4^ The resulting MPI variables were then merged in R Studio with clinical datasets containing demographics, medical history, outcomes, and related information. Clinical data were standardized by distributing a master template spreadsheet to all participating sites to ensure uniform data structure; no further harmonization was applied.^1, 2^

For sites participating in the REgistry of Flow and perfusion imaging for artificial INtelligEnce with PET (REFINE PET)—including Cedars-Sinai Medical Center, Mayo Clinic, Brigham and Women’s Hospital, University of Kansas Medical Center, Montefiore Einstein Medical Center, Intermountain Healthcare, National Autonomous University of Mexico, and University of Naples Federico II—patient data were collected, transferred, processed, and harmonized in an analogous manner, using zipped DICOM (ZCM) files and the dedicated Quantitative PET (QPET) software.^3-6^

**Clinical outcomes**

The primary clinical outcome in this study was all-cause mortality (ACM), which was ascertained using country-specific approaches.^1, 2, 5^ For the eight U.S. sites—Yale University, Columbia University, Cedars-Sinai Medical Center, Mayo Clinic, Brigham and Women’s Hospital, University of Kansas Medical Center, Montefiore Einstein Medical Center, and Intermountain Healthcare—ACM was determined through the National Death Index or the Social Security Death Index.^1, 2, 5^ At the University of Ottawa, ACM was established by chart review at the site or through physician records available in the OACIS Clinical Information System.^1, 2, 5^ For the University of Calgary, ACM was obtained from Alberta Vital Statistics.^1, 2, 5^ At the University of Naples Federico II, ACM was identified using hospital electronic medical records.^1, 2, 5^ For the Mexican site, ACM was determined through chart review or direct patient contact.^1, 2, 5^

**Body composition index**

A support vector machine (SVM) model was developed to derive the body composition index (BCI) which was validated in both an internal cohort and an external cohort.^7^ The SVM model integrated 20 body composition metrics, three demographic variables, and one CT-derived coronary artery calcium (CAC) score. Demographics capture baseline hazard and modify the effects of body composition, while CAC quantifies atherosclerotic burden that body composition alone cannot.^8^ Adding these to BCI produces a more complete, clinically aligned risk signal that improves discrimination, calibration, and classification over a composition-only score while allowing adaptation in multiple clinical specialties with chest CT available. We trained the SVM model through a binary classification task where the death outcome during the entire follow up time was used as the binary label.

**Data preprocessing**

To mitigate the impact of potential outliers, the 0.5th and 99.5th percentiles of the numerical features were calculated using the model development dataset, and feature values were truncated within these bounds. The truncated numerical features were then normalized to a [0, 1] range. Subsequently, the mean and standard deviation of each numerical feature were computed from the model development dataset and used to standardize the features, ensuring zero mean and unit variance.

**External testing scheme**

To assess the generalizability of the SVM model across different sites, we implemented an external testing scheme (Supplementary Figure 1). Patient cases (N=7028) from one positron emission tomography (PET) site—Intermountain Healthcare (ITMT)—and one single-photon emission computed tomography (SPECT) site— University of Ottawa (OTTAWA)—were used as external testing data to evaluate the discriminative and prognostic performance of BCI. Within the remaining cases from seven PET sites—Cedars-Sinai Medical Center (CSMC), Mayo Clinic, Brigham and Women’s Hospital (BWH), Montefiore Einstein Medical Center (MEMC), University of Kansas Medical Center (KUMC), National Autonomous University of Mexico (UNAM), and University of Naples Federico II (UNFII)—and three SPECT sites—Yale University, Columbia University, and University of Calgary, 30% (6444) of them were randomly sampled to create an internal testing cohort and the other 70% (15037) were used as model development cohort.

**Hyperparameter tuning and selection**

The development dataset was randomly and evenly divided into five non-overlapping folds. For each of the five iterations, the fold was used for validation, while the remaining four folds were used for training the model. During hyperparameter tuning (Supplementary Figure 1-A), each candidate hyperparameter configuration was used to train a model on the training data and evaluated by the area under the receiver operating characteristic curve (AUC) on the corresponding validation fold. The average AUC across all five validation folds was used to identify the optimal hyperparameter configuration. The search space and the final selected hyperparameters are detailed in Supplementary Table 1.

**Model training**

During model training (Supplementary Figure 1-B), the development dataset was randomly split into training (80%) and validation (20%) subsets. The training data, along with the optimally selected hyperparameters, was used to train a SVM model, while the validation set was used to select the model for predicting on the internal and the external testing cohorts.

**Model testing**

During testing (Supplementary Figure 1-C), the trained model was applied to the internal and the external testing cohorts separately. The predictions over the internal and the external testing cohorts were used to separately evaluate the performance of BCI generated by the model.

**Model variants**

To facilitate the adoption of our proposed BCI in clinical setting with different data availabilities, we also developed variants of our original SVM model and BCI such as: (1) a BCI which integrated only body composition metrics, thus applicable in any clinical setting where body composition metrics could be obtained such as from computed tomography (CT) or magnetic resonance imaging (MRI);^9, 10^ (2) a BCI which integrated body composition metrics, CT-based CAC score, and demographics, thus adoptable in any clinical scenarios where chest CT were available. We also developed other risk index models for validating the effectiveness of integrating different variables such as: (1) a risk index model which integrated body composition metrics, CT-based CAC score, demographics, medical history, and myocardial perfusion variables; (2) a risk index model incorporating all non-body composition metrics such as demographics, medical histories, CT-based CAC score, and MPI variables. In addition, a baseline risk prediction was developed using demographical variables and CAC score.

**Mortality risk reduction by modifying body composition metric**

Within the external testing cohort of 7028 patients, we divided patients into subgroups by sex, smoking status, hypertension, diabetes, and dyslipidemia, thus 32 subgroups in total. Within each subgroup, we calculated the mean value for each predictor to obtain a synthetic patient. Then, we evaluated how the BCI can be reduced with respect to a particular body composition metric by modifying its value while keeping the values of other predictors fixed.

**Statistical analysis**

All statistical analyses were conducted in [R (version 4.3.2)](https://cran.r-project.org/bin/windows/base/old/) using RStudio. The following open-source R packages were utilized. Baseline characteristics were summarized using [gtsummary (version 2.1.0)](https://cran.r-project.org/web/packages/gtsummary/index.html). Martingale residual plots were generated with [survminer (version 0.5.0)](https://cran.r-project.org/web/packages/survminer/index.html). Proportional hazards assumptions were evaluated using Schoenfeld residuals implemented in [survival (version 3.8-3)](https://cran.r-project.org/web/packages/survival/index.html) and survminer. Kaplan–Meier curves were generated using [adjustedCurves (version 0.11.2)](https://cran.r-project.org/package=adjustedCurves). Cox proportional hazards regression models were fitted using [survivalAnalysis (version 0.3.0)](https://www.rdocumentation.org/packages/survivalAnalysis/versions/0.4.0) and survival. The area under the receiver operating characteristic curve (AUC) and corresponding 95% confidence intervals (CIs) were calculated using [pROC (version 1.18.5)](https://cran.r-project.org/package=pROC). Harrell’s concordance index (C-index) and its 95% CI were computed using survival and [boot (version 1.3-28.1)](https://cran.r-project.org/package=boot). Time-dependent AUC curves were generated using [tdROC (version 2.0)](https://cran.r-project.org/package=tdROC). Calibration curves were constructed using [CalibrationCurves (version 2.0.4)](https://cran.r-project.org/package=CalibrationCurves). Decision curve analysis was performed using [rmda (version 1.6)](https://cran.r-project.org/web/packages/rmda/readme/README.html).To promote transparency, reproducibility, and future research, all statistical analysis code will be made publicly available at <https://github.com/qimagingAI>.

**Supplementary Table 1. Overview of myocardial perfusion imaging acquisition parameters.**

| Modality | Site | kVp | Tube current  (mAs) | Breathing mode | Spacing (mm) | | | Dimensionality | | |
| --- | --- | --- | --- | --- | --- | --- | --- | --- | --- | --- |
|  |  |  |  |  | x | y | z | x | y | z |
| SPECT/CT | Yale | 120 | 20-150 | Normal breathing | 0.98 | 0.98 | 2.5-5.0 | 512 | 512 | 27-134 |
|  | Calgary | 120 | 20-300 | End-expiratory breath hold | 0.49-0.98 | 0.49-0.98 | 2.5-5.0 | 512 | 512 | 45-157 |
|  | Columbia | 90-120 | 31-125 | Normal breathing | 1.17 | 1.17 | 3.0 | 512 | 512 | 54-166 |
|  | Ottawa | 130 | 24-53 | End-expiratory breath hold | 0.98 | 0.98 | 5.0 | 512 | 512 | 28-84 |
| PET/CT | CSMC | 100 | 11-13 | Normal breathing | 1.37 | 1.37 | 3.0 | 512 | 512 | 10-75 |
|  | MEMC | 120 | 54-163 | Normal breathing | 1.12-1.17 | 1.12-1.17 | 3.0 | 512 | 512 | 60-63 |
|  | KUMC | 120-140 | 50-244 | Normal breathing | 0.70-0.98 | 0.70-0.98 | 3.75 | 512 | 512 | 71 |
|  | UNAM | 80-120 | 33-595 | End-expiratory breath hold | 0.98-1.37 | 0.98-1.37 | 3.0 | 512 | 512 | 82-132 |
|  | BWH | 80-140 | 10-300 | Normal breathing | 0.98-1.37 | 0.98-1.37 | 2.5-5.0 | 512 | 512 | 32-71 |
|  | Mayo | 120-140 | 20-180 | Normal breathing | 1.37 | 1.37 | 3.75 | 512 | 512 | 41-47 |
|  | ITMT | 120-130 | 16-407 | Normal breathing | 1.37-1.52 | 1.37-1.52 | 2 | 512 | 512 | 75-111 |
|  | UNFII | 120-140 | 15-260 | Normal breathing | 0.70-1.17 | 0.70-1.17 | 3.0-3.75 | 512 | 512 | 59-89 |

Sites highlighted by yellow were used as external testing cohort. Yale – Yale University, Calgary – University of Calgary, Columbia – Columbia University, Ottawa – Ottawa University, CSMC – Cedars-Sinai Medical Center, MEMC – Montefiore Einstein Medical Center, KUMC – University of Kansas Medical Center, UNAM – National Autonomous University of Mexico, BWH – Brigham and Women’s Hospital, Mayo – Mayo Clinic, ITMT – Intermountain Healthcare, UNFII – University of Naples Federico II.

**Supplementary Table 2. Hyperparameter search space of support vector machine (SVM) for developing body composition index (BCI) model.**

| Hyperparameter | Searching space | Selected value |
| --- | --- | --- |
| “**C**”: regularization parameter in C-SVM with smaller value for stronger regularization and less overfitting models | 0.1, 1, 10, 100 | 1 |
| “**gamma**”: kernel coefficient | ‘scale’, 0.01, 0.001 | 0.01 |
| “**kernel**”: kernel type | ‘rbf’ | ‘rbf’ |
| “**class_weight**”: weights specification for different classes | ‘balanced’ | ‘balanced’ |
| “**eval_metric**”: evaluation metric for hyperparameter selection and model selection | “auc” | “auc” |
| “**seed**”: random seed for data splitting | 42 | 42 |

All other hyperparameters used default values.

**Supplementary Table 3. Internal and external testing cohort patients stratified by all-cause mortality.**

| Characteristics | Overall  N = 13472 | Internal testing  cohort  N=6444 | | External testing  Cohort  N=7028 | |
| --- | --- | --- | --- | --- | --- |
|  |  | Alive  N=5362 | Death  N=1082 | Alive  N=6079 | Death  N=949 |
| **Demographics** | | | | | |
| Age [year] | 66 (57, 74) | 65 (57, 73) | 73 (64, 81) | 65 (57, 72) | 73 (64, 81) |
| Sex |  |  |  |  |  |
| Female | 5922 (44%) | 2437 (45%) | 416 (38%) | 2697 (44%) | 372 (39%) |
| Male | 7550 (56%) | 2925 (55%) | 666 (62%) | 3382 (56%) | 577 (61%) |
| BMI [kg/m^2^] | 30.2 (26.0, 35.3) | 30.0 (25.8, 35.5) | 27.8 (24.4, 32.9) | 31.1 (26.8, 35.8) | 29.5 (25.1, 34.3) |
| Race |  |  |  |  |  |
| American Indian | 59 (0.4%) | 10 (0.2%) | 4 (0.4%) | 37 (0.6%) | 8 (0.8%) |
| Asian | 253 (1.9%) | 137 (2.6%) | 16 (1.5%) | 89 (1.5%) | 11 (1.2%) |
| Black | 993 (7.4%) | 775 (14%) | 141 (13%) | 71 (1.2%) | 6 (0.6%) |
| Pacific Islander | 145 (1.1%) | 12 (0.2%) | 2 (0.2%) | 100 (1.6%) | 31 (3.3%) |
| White | 8802 (65%) | 3058 (57%) | 753 (70%) | 4169 (69%) | 822 (87%) |
| Unknown | 3220 (24%) | 1370 (26%) | 166 (15%) | 1613 (27%) | 71 (7.5%) |
| **Medical histories** | | | | | |
| Hypertension | 10457 (78%) | 3702 (69%) | 907 (84%) | 4948 (81%) | 900 (95%) |
| Diabetes mellitus | 4531 (34%) | 1783 (33%) | 476 (44%) | 1843 (30%) | 429 (45%) |
| Dyslipidemia | 9426 (70%) | 3193 (60%) | 771 (71%) | 4643 (76%) | 819 (86%) |
| Family history | 5012 (37%) | 1416 (26%) | 208 (19%) | 2921 (48%) | 467 (49%) |
| Smoking | 3308 (25%) | 740 (14%) | 126 (12%) | 2016 (33%) | 426 (45%) |
| **Myocardial perfusion imaging and coronary artery calcium score variables** | | | | | |
| Stress total perfusion deficit [%] | 4 (2, 10) | 3 (1, 8) | 8 (3, 18) | 5 (2, 11) | 6 (2, 13) |
| Myocardial flow reserve | 2.38 (1.88, 2.94) | 2.44 (1.92, 3.08) | 1.87 (1.44, 2.41) | 2.51 (2.06, 3.02) | 1.95 (1.53, 2.38) |
| Left ventricle ejection fraction [%] | 68 (58, 76) | 66 (56, 73) | 59 (42, 69) | 72 (62, 80) | 65 (48, 75) |
| log10(CAC score+1) [AU] | 2.01 (0.00, 2.92) | 1.84 (0.00, 2.84) | 2.62 (1.40, 3.18) | 1.81 (0.00, 2.84) | 2.76 (2.04, 3.27) |
| **Body composition metrics** | | | | | |
| VAT volume index [cm^3^/m^2^] | 401 (269, 560) | 351 (234, 500) | 328 (221, 472) | 449 (316, 611) | 458 (320, 620) |
| VAT attenuation [HU] | -83 (-88, -78) | -86 (-91, -81) | -83 (-88, -78) | -82 (-86, -77) | -77 (-83, -70) |
| VAT standard deviation [HU] | 62 (55, 71) | 61 (53, 76) | 61 (51, 88) | 62 (56, 69) | 64 (59, 70) |
| EAT volume index [cm^3^/m^2^] | 40 (27, 56) | 43 (29, 61) | 41 (27, 60) | 37 (25, 50) | 39 (27, 54) |
| EAT attenuation [HU] | -67 (-72, -63) | -66 (-72, -58) | -68 (-73, -63) | -68 (-71, -64) | -67 (-71, -63) |
| EAT standard deviation [HU] | 44 (39, 50) | 46 (36, 54) | 43 (33, 54) | 43 (40, 47) | 45 (40, 50) |
| Bone volume index [cm^3^/m^2^] | 302 (265, 345) | 302 (263, 347) | 302 (263, 342) | 302 (266, 344) | 307 (267, 347) |
| Bone attenuation [HU] | 255 (226, 285) | 256 (227, 287) | 234 (208, 264) | 258 (231, 286) | 245 (215, 276) |
| Bone standard deviation [HU] | 202 (179, 226) | 198 (174, 222) | 189 (160, 221) | 206 (186, 229) | 208 (183, 231) |
| SM volume index [cm^3^/m^2^] | 731 (592, 903) | 742 (603, 915) | 665 (536, 810) | 748 (606, 919) | 651 (518, 812) |
| SM attenuation [HU] | 24 (18, 30) | 27 (21, 33) | 23 (16, 29) | 23 (18, 28) | 19 (14, 23) |
| SM standard deviation [HU] | 47 (40, 55) | 48 (38, 62) | 44 (38, 59) | 46 (41, 52) | 50 (45, 55) |
| IMAT volume index [cm^3^/m^2^] | 82 (57, 120) | 90 (59, 139) | 99 (67, 146) | 76 (54, 105) | 81 (60, 111) |
| IMAT attenuation [HU] | -69.2 (-73.1, -65.6) | -69.7 (-73.8, -66.0) | -67.1 (-71.3, -63.3) | -69.7 (-73.3, -66.3) | -65.7 (-69.3, -62.5) |
| IMAT standard deviation [HU] | 43 (35, 53) | 43 (31, 60) | 36 (29, 54) | 44 (38, 51) | 45 (40, 52) |
| SAT volume index [cm^3^/m^2^] | 1223 (741, 1896) | 1224 (755, 1956) | 1039 (682, 1664) | 1275 (768, 1912) | 1052 (602, 1747) |
| SAT attenuation [HU] | -100 (-104, -94) | -101 (-105, -94) | -93 (-102, -85) | -100 (-104, -96) | -97 (-102, -91) |
| SAT standard deviation [HU] | 34 (30, 42) | 39 (31, 54) | 37 (31, 54) | 33 (29, 36) | 34 (30, 38) |
| Ratio SM2AT | 0.40 (0.27, 0.62) | 0.41 (0.28, 0.64) | 0.42 (0.28, 0.65) | 0.40 (0.27, 0.61) | 0.38 (0.25, 0.62) |
| Ratio ECTOPIC2AT | 0.31 (0.22, 0.42) | 0.29 (0.20, 0.40) | 0.32 (0.23, 0.42) | 0.32 (0.22, 0.42) | 0.37 (0.26, 0.48) |
| **Outcome** | | | | | |
| Death follow up [year] | 3.83 (2.02, 4.99) | 3.39 (1.97, 5.52) | 2.67 (1.10, 5.17) | 4.49 (2.60, 4.95) | 2.39 (1.13, 3.48) |
| **Imaging acquisition protocols** | | | | | |
| Site |  |  |  |  |  |
| Brigham and Women’s Hospital | 1163 (8.6%) | 722 (13%) | 441 (41%) | 0 (0%) | 0 (0%) |
| University of Calgary | 877 (6.5%) | 791 (15%) | 86 (7.9%) | 0 (0%) | 0 (0%) |
| Columbia University | 574 (4.3%) | 537 (10%) | 37 (3.4%) | 0 (0%) | 0 (0%) |
| Cedars-Sina Medical Center | 1375 (10%) | 1053 (20%) | 322 (30%) | 0 (0%) | 0 (0%) |
| Intermountain Healthcare | 5611 (42%) | 0 (0%) | 0 (0%) | 4690 (77%) | 921 (97%) |
| University of Kansas Medical Center | 164 (1.2%) | 141 (2.6%) | 23 (2.1%) | 0 (0%) | 0 (0%) |
| Mayo Clinic | 473 (3.5%) | 411 (7.7%) | 62 (5.7%) | 0 (0%) | 0 (0%) |
| Montefiore Medical Center | 281 (2.1%) | 265 (4.9%) | 16 (1.5%) | 0 (0%) | 0 (0%) |
| UNAM | 101 (0.7%) | 75 (1.4%) | 26 (2.4%) | 0 (0%) | 0 (0%) |
| University of Naples Federico II | 126 (0.9%) | 121 (2.3%) | 5 (0.5%) | 0 (0%) | 0 (0%) |
| University of Ottawa | 1417 (11%) | 0 (0%) | 0 (0%) | 1389 (23%) | 28 (3.0%) |
| Yale University | 1310 (9.7%) | 1246 (23%) | 64 (5.9%) | 0 (0%) | 0 (0%) |
| KVP |  |  |  |  |  |
| 80 | 1 (<0.1%) | 1 (<0.1%) | 0 (0%) | 0 (0%) | 0 (0%) |
| 90 | 1 (<0.1%) | 1 (<0.1%) | 0 (0%) | 0 (0%) | 0 (0%) |
| 100 | 1375 (10%) | 1053 (20%) | 322 (30%) | 0 (0%) | 0 (0%) |
| 120 | 7299 (54%) | 3932 (73%) | 436 (40%) | 2432 (40%) | 499 (53%) |
| 130 | 4097 (30%) | 0 (0%) | 0 (0%) | 3647 (60%) | 450 (47%) |
| 140 | 699 (5.2%) | 375 (7.0%) | 324 (30%) | 0 (0%) | 0 (0%) |
| Reconstruction diameter [mm] |  |  |  |  |  |
| 357 | 160 (1.2%) | 138 (2.6%) | 22 (2.0%) | 0 (0%) | 0 (0%) |
| 358 | 6 (<0.1%) | 6 (0.1%) | 0 (0%) | 0 (0%) | 0 (0%) |
| 500 | 5667 (42%) | 3453 (64%) | 797 (74%) | 1389 (23%) | 28 (3.0%) |
| 600 | 975 (7.2%) | 917 (17%) | 58 (5.4%) | 0 (0%) | 0 (0%) |
| 700 | 3733 (28%) | 848 (16%) | 205 (19%) | 2258 (37%) | 422 (44%) |
| 780 | 2931 (22%) | 0 (0%) | 0 (0%) | 2432 (40%) | 499 (53%) |
| Tube current [mAs] | 32 (22, 42) | 31 (13, 60) | 13 (10, 30) | 35 (27, 38) | 37 (26, 38) |
| Convolution kernel |  |  |  |  |  |
| I26f | 2931 (22%) | 0 (0%) | 0 (0%) | 2432 (40%) | 499 (53%) |
| B | 975 (7.2%) | 917 (17%) | 58 (5.4%) | 0 (0%) | 0 (0%) |
| B18f | 59 (0.4%) | 43 (0.8%) | 16 (1.5%) | 0 (0%) | 0 (0%) |
| B19s | 2680 (20%) | 0 (0%) | 0 (0%) | 2258 (37%) | 422 (44%) |
| B25f | 39 (0.3%) | 30 (0.6%) | 9 (0.8%) | 0 (0%) | 0 (0%) |
| B30f | 3 (<0.1%) | 2 (<0.1%) | 1 (<0.1%) | 0 (0%) | 0 (0%) |
| B30s | 1417 (11%) | 0 (0%) | 0 (0%) | 1389 (23%) | 28 (3.0%) |
| PET AC | 2 (<0.1%) | 1 (<0.1%) | 1 (<0.1%) | 0 (0%) | 0 (0%) |
| SOFT | 1163 (8.6%) | 722 (13%) | 441 (41%) | 0 (0%) | 0 (0%) |
| STANDARD | 4203 (31%) | 3647 (68%) | 556 (51%) | 0 (0%) | 0 (0%) |
| Manufacturer |  |  |  |  |  |
| GE | 3993 (30%) | 3317 (62%) | 676 (62%) | 0 (0%) | 0 (0%) |
| Philips | 975 (7.2%) | 917 (17%) | 58 (5.4%) | 0 (0%) | 0 (0%) |
| Siemens | 8504 (63%) | 1128 (21%) | 348 (32%) | 6079 (100%) | 949 (100%) |
| Slice thickness [mm] |  |  |  |  |  |
| 2 | 5611 (42%) | 0 (0%) | 0 (0%) | 4690 (77%) | 921 (97%) |
| 2.5 | 1235 (9.2%) | 1172 (22%) | 63 (5.8%) | 0 (0%) | 0 (0%) |
| 3 | 2451 (18%) | 2045 (38%) | 406 (38%) | 0 (0%) | 0 (0%) |
| 3.27 | 2 (<0.1%) | 0 (0%) | 2 (0.2%) | 0 (0%) | 0 (0%) |
| 3.75 | 643 (4.8%) | 558 (10%) | 85 (7.9%) | 0 (0%) | 0 (0%) |
| 5 | 3530 (26%) | 1587 (30%) | 526 (49%) | 1389 (23%) | 28 (3.0%) |
| Pixel spacing x/y [mm] |  |  |  |  |  |
| 0.7 | 166 (1.2%) | 144 (2.7%) | 22 (2.0%) | 0 (0%) | 0 (0%) |
| 0.98 | 4291 (32%) | 2399 (45%) | 475 (44%) | 1389 (23%) | 28 (3.0%) |
| 1.17 | 975 (7.2%) | 917 (17%) | 58 (5.4%) | 0 (0%) | 0 (0%) |
| 1.37 | 5109 (38%) | 1902 (35%) | 527 (49%) | 2258 (37%) | 422 (44%) |
| 1.52 | 2931 (22%) | 0 (0%) | 0 (0%) | 2432 (40%) | 499 (53%) |

Bold indicates statistical significance. AU – Agatston unit, HU – Hounsfield unit, VAT – visceral adipose tissue, EAT – epicardial adipose tissue, SM – skeletal muscle, IMAT – intramuscular adipose tissue, SAT, subcutaneous adipose tissue, Ratio SM2AT – ratio of SM volume to total adipose tissue (including VAT, EAT, IMAT, and SAT) volume, Ratio ECTOPIC2AT – ratio of ectopic adipose tissue (including VAT, EAT, and IMAT) volume to total adipose tissue volume.

**Supplementary Table 4. Baseline characteristics of the development cohort patients stratified by all-cause mortality.**

| Characteristics | Overall N = 15037 | No death  N = 12371 | With death  N=2666 | p-value |
| --- | --- | --- | --- | --- |
| **Demographics** | | | | |
| Age [year] | 66 (58, 74) | 65 (57, 73) | 72 (64, 80) | **<0.0001** |
| Sex |  |  |  | **<0.0001** |
| Female | 6562 (44%) | 5504 (44%) | 1058 (40%) |  |
| Male | 8475 (56%) | 6867 (56%) | 1608 (60%) |  |
| BMI [kg/m^2^] | 29.6 (25.7, 34.9) | 29.9 (26.0, 35.3) | 28.2 (24.7, 33.3) | **<0.0001** |
| Race |  |  |  | **<0.0001** |
| American Indian or Alaska Native | 39 (0.3%) | 30 (0.2%) | 9 (0.3%) |  |
| Asian | 345 (2.3%) | 289 (2.3%) | 56 (2.1%) |  |
| Black or African American | 2162 (14%) | 1738 (14%) | 424 (16%) |  |
| Native Hawaiian or Other Pacific Islander | 22 (0.1%) | 19 (0.2%) | 3 (0.1%) |  |
| White | 8982 (60%) | 7168 (58%) | 1814 (68%) |  |
| Unknown | 3487 (23%) | 3127 (25%) | 360 (14%) |  |
| **Medical history** | | | | |
| Hypertension | 10789 (72%) | 8593 (69%) | 2196 (82%) | **<0.0001** |
| Diabetes mellitus | 5159 (34%) | 3991 (32%) | 1168 (44%) | **<0.0001** |
| Dyslipidemia | 9078 (60%) | 7329 (59%) | 1749 (66%) | **<0.0001** |
| Family history of CAD | 3757 (25%) | 3287 (27%) | 470 (18%) | **<0.0001** |
| Smoking | 2083 (14%) | 1759 (14%) | 324 (12%) | **0.0051** |
| **Myocardial perfusion imaging findings and coronary artery calcium score** | | | | |
| Stress total perfusion deficit [%] | 4 (2, 10) | 4 (1, 8) | 8 (3, 18) | **<0.0001** |
| Left ventricle ejection fraction [%] | 65 (54, 73) | 66 (56, 73) | 58 (42, 69) | **<0.0001** |
| log10(CAC score+1) [AU] | 2.00 (0.00, 2.92) | 1.83 (0.00, 2.84) | 2.56 (1.51, 3.18) | **<0.0001** |
| **Body composition measures** | | | | |
| VAT volume index [cm^3^/m^2^] | 355 (238, 502) | 361 (242, 507) | 328 (218, 471) | **<0.0001** |
| VAT attenuation [HU] | -86 (-91, -81) | -86 (-91, -82) | -83 (-88, -78) | **<0.0001** |
| VAT standard deviation [HU] | 61 (53, 78) | 61 (53, 76) | 63 (52, 90) | **<0.0001** |
| EAT volume index [cm^3^/m^2^] | 44 (29, 62) | 44 (30, 62) | 42 (26, 60) | **<0.0001** |
| EAT attenuation [HU] | -66 (-72, -59) | -66 (-72, -58) | -68 (-73, -63) | **<0.0001** |
| EAT standard deviation [HU] | 46 (35, 54) | 46 (36, 54) | 42 (34, 54) | **<0.0001** |
| Bone volume index [cm^3^/m^2^] | 302 (264, 347) | 303 (265, 348) | 297 (260, 341) | **<0.0001** |
| Bone attenuation [HU] | 253 (223, 285) | 256 (227, 288) | 236 (207, 268) | **<0.0001** |
| Bone standard deviation [HU] | 197 (173, 223) | 198 (175, 223) | 188 (161, 222) | **<0.0001** |
| SM volume index [cm^3^/m^2^] | 732 (595, 895) | 747 (610, 912) | 663 (540, 809) | **<0.0001** |
| SM attenuation [HU] | 26 (20, 33) | 27 (20, 33) | 23 (16, 29) | **<0.0001** |
| SM standard deviation [HU] | 47 (38, 62) | 48 (37, 62) | 44 (38, 59) | **0.0037** |
| IMAT volume index [cm^3^/m^2^] | 92 (60, 140) | 90 (59, 138) | 97 (65, 150) | **<0.0001** |
| IMAT attenuation [HU] | -69 (-73, -66) | -70 (-74, -66) | -67 (-72, -63) | **<0.0001** |
| IMAT standard deviation [HU] | 42 (31, 60) | 43 (31, 61) | 36 (29, 54) | **<0.0001** |
| SAT volume index [cm^3^/m^2^] | 1190 (753, 1854) | 1207 (765, 1884) | 1117 (696, 1713) | **<0.0001** |
| SAT attenuation [HU] | -100 (-105, -93) | -101 (-105, -95) | -94 (-102, -85) | **<0.0001** |
| SAT standard deviation [HU] | 39 (31, 54) | 39 (31, 54) | 37 (32, 54) | 0.7534 |
| Ratio SM2AT | 0.42 (0.28, 0.63) | 0.42 (0.28, 0.64) | 0.41 (0.27, 0.62) | **0.0098** |
| Ratio ECTOPIC2AT | 0.30 (0.21, 0.41) | 0.30 (0.20, 0.41) | 0.31 (0.22, 0.41) | **0.0001** |
| **Outcomes** | | | | |
| Death | 2666 (18%) | 0 (0%) | 2666 (100%) | **<0.0001** |
| Death follow up [year] | 3.35 (1.85, 5.45) | 3.44 (1.99, 5.50) | 2.73 (1.10, 5.18) | **<0.0001** |
| **Imaging acquisition** | | | | |
| KVP |  |  |  | **<0.0001** |
| 80 | 1 (<0.1%) | 1 (<0.1%) | 0 (0%) |  |
| 100 | 3226 (21%) | 2403 (19%) | 823 (31%) |  |
| 120 | 10280 (68%) | 9190 (74%) | 1090 (41%) |  |
| 140 | 1530 (10%) | 777 (6.3%) | 753 (28%) |  |
| Reconstruction diameter [mm] |  |  |  | **<0.0001** |
| 250 | 1 (<0.1%) | 1 (<0.1%) | 0 (0%) |  |
| 287 | 1 (<0.1%) | 1 (<0.1%) | 0 (0%) |  |
| 357 | 409 (2.7%) | 355 (2.9%) | 54 (2.0%) |  |
| 358 | 11 (<0.1%) | 11 (<0.1%) | 0 (0%) |  |
| 500 | 9845 (65%) | 7916 (64%) | 1929 (72%) |  |
| 576 | 1 (<0.1%) | 1 (<0.1%) | 0 (0%) |  |
| 600 | 2214 (15%) | 2090 (17%) | 124 (4.7%) |  |
| 608 | 1 (<0.1%) | 1 (<0.1%) | 0 (0%) |  |
| 700 | 2554 (17%) | 1995 (16%) | 559 (21%) |  |
| Tube current [mAs] | 20 (13, 60) | 31 (13, 60) | 13 (10, 29) | **<0.0001** |
| Convolution kernel |  |  |  | **<0.0001** |
| B | 2214 (15%) | 2091 (17%) | 123 (4.6%) |  |
| B18f | 146 (1.0%) | 92 (0.7%) | 54 (2.0%) |  |
| B25f | 81 (0.5%) | 54 (0.4%) | 27 (1.0%) |  |
| B30f | 4 (<0.1%) | 4 (<0.1%) | 0 (0%) |  |
| PET AC | 1 (<0.1%) | 1 (<0.1%) | 0 (0%) |  |
| SOFT | 2653 (18%) | 1569 (13%) | 1084 (41%) |  |
| STANDARD | 9938 (66%) | 8560 (69%) | 1378 (52%) |  |
| Manufacturer |  |  |  | **<0.0001** |
| GE | 9366 (62%) | 7727 (62%) | 1639 (61%) |  |
| Philips | 2214 (15%) | 2091 (17%) | 123 (4.6%) |  |
| Siemens | 3457 (23%) | 2553 (21%) | 904 (34%) |  |
| Slice thickness [mm] |  |  |  | **<0.0001** |
| 2.5 | 2942 (20%) | 2782 (22%) | 160 (6.0%) |  |
| 3 | 5671 (38%) | 4644 (38%) | 1027 (39%) |  |
| 3.27 | 3 (<0.1%) | 0 (0%) | 3 (0.1%) |  |
| 3.75 | 1594 (11%) | 1381 (11%) | 213 (8.0%) |  |
| 5 | 4827 (32%) | 3564 (29%) | 1263 (47%) |  |
| Pixel spacing x/y [mm] |  |  |  | **<0.0001** |
| 0.49 | 1 (<0.1%) | 1 (<0.1%) | 0 (0%) |  |
| 0.56 | 1 (<0.1%) | 1 (<0.1%) | 0 (0%) |  |
| 0.7 | 420 (2.8%) | 366 (3.0%) | 54 (2.0%) |  |
| 0.98 | 6619 (44%) | 5513 (45%) | 1106 (41%) |  |
| 1.12 | 1 (<0.1%) | 1 (<0.1%) | 0 (0%) |  |
| 1.17 | 2214 (15%) | 2090 (17%) | 124 (4.7%) |  |
| 1.19 | 1 (<0.1%) | 1 (<0.1%) | 0 (0%) |  |
| 1.37 | 5780 (38%) | 4398 (36%) | 1382 (52%) |  |
| **Others** | | | | |
| Site |  |  |  | **<0.0001** |
| Brigham and Women’s Hospital | 2654 (18%) | 1569 (13%) | 1085 (41%) |  |
| University of Calgary | 2013 (13%) | 1841 (15%) | 172 (6.5%) |  |
| Columbia University | 1292 (8.6%) | 1204 (9.7%) | 88 (3.3%) |  |
| Cedars-Sina Medical Center | 3226 (21%) | 2403 (19%) | 823 (31%) |  |
| University of Kansas Medical Center | 416 (2.8%) | 361 (2.9%) | 55 (2.1%) |  |
| Mayo Clinic | 1166 (7.8%) | 1008 (8.1%) | 158 (5.9%) |  |
| Montefiore Einstein Medical Center | 618 (4.1%) | 586 (4.7%) | 32 (1.2%) |  |
| UNAM | 231 (1.5%) | 150 (1.2%) | 81 (3.0%) |  |
| University of Naples Federico II | 315 (2.1%) | 312 (2.5%) | 3 (0.1%) |  |
| Yale University | 3106 (21%) | 2937 (24%) | 169 (6.3%) |  |
| Modality |  |  |  | **<0.0001** |
| Positron emission tomography | 8626 (57%) | 6389 (52%) | 2237 (84%) |  |
| Single photon emission computed tomography | 6411 (43%) | 5982 (48%) | 429 (16%) |  |

Bold indicates statistical significance. Frequency (%) or median (IQR). VAT – visceral adipose tissue, EAT – epicardial adipose tissue, SM – skeletal muscle, IMAT – intramuscular adipose tissue, SAT – subcutaneous adipose tissue, ratio SM2AT – ratio of skeletal muscle volume to all adipose tissue volume, ratio ECTOPIC2AT – ratio of ectopic adipose tissue volume (including EAT, VAT, and IMAT) to all adipose tissue volume, CAC – coronary artery calcium, IQR – interquartile range, AU – Agatston unit, HU – Hounsfield unit, CAD – coronary artery disease, UNAM – National Autonomous University of Mexico.

**Supplementary Table 5: Prediction performance comparison between body composition index (BCI) and other baselines in external testing cohort.**

| Predictor | AUC | p-value  (AUC) | C-index | p-value  (C-index) |
| --- | --- | --- | --- | --- |
| Age | 0.68 [0.66, 0.70] | **<0.0001** | 0.65 [0.63, 0.67] | **<0.0001** |
| Body mass index (BMI) | 0.57 [0.55, 0.59] | **<0.0001** | 0.44 [0.42, 0.46] | **<0.0001** |
| Stress total perfusion deficit | 0.55 [0.53, 0.57] | **<0.0001** | 0.58 [0.56, 0.60] | **<0.0001** |
| Coronary artery calcium score | 0.68 [0.66, 0.70] | **<0.0001** | 0.66 [0.65, 0.68] | **<0.0001** |
| Left ventricular ejection fraction | 0.63 [0.61, 0.65] | **<0.0001** | 0.36 [0.34, 0.38] | **<0.0001** |
| Ratio SM2AT | 0.51 [0.49, 0.53] | **<0.0001** | 0.49 [0.47, 0.51] | **<0.0001** |
| Ratio ECTOPIC2AT | 0.60 [0.58, 0.62] | **<0.0001** | 0.59 [0.57, 0.61] | **<0.0001** |
| Bone attenuation | 0.58 [0.56, 0.60] | **<0.0001** | 0.42 [0.40, 0.44] | **<0.0001** |
| Bone SD | 0.51 [0.49, 0.53] | **<0.0001** | 0.49 [0.47, 0.51] | **<0.0001** |
| Bone volume index | 0.52 [0.50, 0.54] | **<0.0001** | 0.53 [0.51, 0.55] | **<0.0001** |
| SM attenuation | 0.68 [0.67, 0.70] | **<0.0001** | 0.34 [0.33, 0.36] | **<0.0001** |
| SM SD | 0.63 [0.61, 0.65] | **<0.0001** | 0.58 [0.57, 0.60] | **<0.0001** |
| SM volume index | 0.62 [0.60, 0.64] | **<0.0001** | 0.41 [0.39, 0.43] | **<0.0001** |
| EAT attenuation | 0.53 [0.51, 0.55] | **<0.0001** | 0.52 [0.50, 0.54] | **<0.0001** |
| EAT SD | 0.57 [0.55, 0.59] | **<0.0001** | 0.57 [0.55, 0.59] | **<0.0001** |
| EAT volume index | 0.54 [0.52, 0.56] | **<0.0001** | 0.54 [0.52, 0.56] | **<0.0001** |
| IMAT attenuation | 0.71 [0.69, 0.72] | **<0.0001** | 0.68 [0.66, 0.70] | **<0.0001** |
| IMAT SD | 0.54 [0.53, 0.56] | **<0.0001** | 0.51 [0.49, 0.53] | **<0.0001** |
| IMAT volume index | 0.55 [0.53, 0.57] | **<0.0001** | 0.55 [0.53, 0.57] | **<0.0001** |
| SAT attenuation | 0.64 [0.62, 0.66] | **<0.0001** | 0.63 [0.62, 0.65] | **<0.0001** |
| SAT SD | 0.56 [0.54, 0.58] | **<0.0001** | 0.54 [0.52, 0.56] | **<0.0001** |
| SAT volume index | 0.57 [0.55, 0.59] | **<0.0001** | 0.44 [0.42, 0.46] | **<0.0001** |
| VAT attenuation | 0.67 [0.65, 0.69] | **<0.0001** | 0.64 [0.62, 0.66] | **<0.0001** |
| VAT SD | 0.57 [0.55, 0.58] | **<0.0001** | 0.51 [0.50, 0.53] | **<0.0001** |
| VAT volume index | 0.51 [0.49, 0.53] | **<0.0001** | 0.51 [0.49, 0.53] | **<0.0001** |
| BCI: body composition metrics | 0.75 [0.73, 0.77] | **<0.0001** | 0.72 [0.71, 0.74] | **<0.0001** |
| BCI: body composition metrics + demographics | 0.77 [0.75, 0.78] | **<0.0001** | 0.74 [0.72, 0.75] | **<0.0001** |
| BCI: body composition metrics + CAC | 0.77 [0.75, 0.78] | **<0.0001** | 0.74 [0.72, 0.75] | **<0.0001** |
| BCI: body composition metric + demographics + CAC | 0.78 [0.76, 0.79] | **<0.0001** | 0.75 [0.73, 0.76] | **<0.0001** |
| Risk index: demographics + CAC | 0.72 [0.71, 0.74] | **<0.0001** | 0.69 [0.68, 0.71] | **<0.0001** |

Bold indicates statistical significance. AUC – area under curve, C-index – concordance index, SM2AT – ratio of skeletal muscle (SM) volume to adipose tissue volume, ECTOPIC2AT – ratio of ectopic adipose tissue volume to adipose tissue volume, SD – standard deviation, EAT – epicardial adipose tissue, IMAT – intramuscular adipose tissue, SAT – subcutaneous adipose tissue, VAT – visceral adipose tissue, MPI – myocardial perfusion imaging, CAC – coronary artery calcium.

**Supplementary Table 6: Reliable discriminative prediction performance of the body composition index (BCI), integrating CT-derived body composition metrics, coronary artery calcium score, and demographic variables, in the external testing cohort overall and across subgroups defined by patient characteristics and imaging acquisition protocol.**

| Population | N | AUC | C-index |
| --- | --- | --- | --- |
| Entire | 7028 | 0.78 [0.76, 0.79] | 0.75 [0.73, 0.76] |
| **By sex** |  |  |  |
| Female | 3069 | 0.78 [0.75, 0.80] | 0.75 [0.72, 0.78] |
| Male | 3959 | 0.78 [0.76, 0.80] | 0.75 [0.73, 0.77] |
| **By age [year]** |  |  |  |
| Age < 60 | 2145 | 0.73 [0.68, 0.77] | 0.71 [0.67, 0.76] |
| 60 ≤ Age ≤ 69 | 2258 | 0.76 [0.73, 0.79] | 0.74 [0.71, 0.77] |
| 70 ≤ Age | 2625 | 0.73 [0.71, 0.76] | 0.71 [0.69, 0.73] |
| **By body mass index (BMI) [kg/m^2^]** |  |  |  |
| BMI < 30 | 3181 | 0.78 [0.76, 0.80] | 0.75 [0.73, 0.77] |
| 30 ≤ BMI < 35 | 1940 | 0.77 [0.74, 0.80] | 0.74 [0.70, 0.77] |
| 35 ≤ BMI < 40 | 1126 | 0.78 [0.73, 0.82] | 0.75 [0.71, 0.80] |
| 40 ≤ BMI | 781 | 0.76 [0.70, 0.81] | 0.74 [0.68, 0.79] |
| **By hypertension** |  |  |  |
| No hypertension | 1180 | 0.74 [0.66, 0.82] | 0.73 [0.65, 0.81] |
| With hypertension | 5848 | 0.77 [0.75, 0.78] | 0.74 [0.72, 0.75] |
| **By diabetes** |  |  |  |
| No diabetes | 4757 | 0.78 [0.75, 0.80] | 0.75 [0.73, 0.77] |
| With diabetes | 2271 | 0.77 [0.74, 0.79] | 0.73 [0.71, 0.75] |
| **By dyslipidemia** |  |  |  |
| No dyslipidemia | 1566 | 0.77 [0.73, 0.82] | 0.75 [0.71, 0.80] |
| With dyslipidemia | 5462 | 0.77 [0.76, 0.79] | 0.74 [0.73, 0.76] |
| **By family history of coronary artery disease (CAD)** |  |  |  |
| No family history of CAD | 3639 | 0.78 [0.76, 0.80] | 0.75 [0.73, 0.77] |
| With family history of CAD | 3389 | 0.78 [0.76, 0.80] | 0.75 [0.73, 0.77] |
| **By smoking** |  |  |  |
| No smoking | 4586 | 0.80 [0.78, 0.81] | 0.76 [0.74, 0.78] |
| With smoking | 2442 | 0.75 [0.72, 0.77] | 0.72 [0.70, 0.75] |
| **By stress total perfusion deficit (TPD)** |  |  |  |
| Stress TPD < 1% | 996 | 0.76 [0.71, 0.81] | 0.73 [0.67, 0.78] |
| 1% ≤ Stress TPD < 5% | 2612 | 0.76 [0.73, 0.79] | 0.74 [0.71, 0.77] |
| 5% ≤ Stress TPD < 10% | 1500 | 0.80 [0.77, 0.83] | 0.76 [0.73, 0.79] |
| 10% ≤ Stress TPD | 1920 | 0.79 [0.76, 0.81] | 0.74 [0.72, 0.77] |
| **By left ventricle ejection fraction (LVEF)** |  |  |  |
| LVEF < 50% | 863 | 0.71 [0.67, 0.74] | 0.67 [0.64, 0.70] |
| 50% ≤ LVEF ≤ 70% | 2351 | 0.78 [0.76, 0.81] | 0.74 [0.71, 0.77] |
| 70% < LVEF | 3814 | 0.77 [0.74, 0.79] | 0.75 [0.72, 0.77] |
| **By coronary artery calcium (CAC) score [AU]** |  |  |  |
| CAC = 0 | 2244 | 0.73 [0.68, 0.78] | 0.70 [0.65, 0.76] |
| 1 ≤ CAC ≤ 99 | 1245 | 0.71 [0.66, 0.76] | 0.68 [0.63, 0.74] |
| 100 ≤ CAC ≤ 299 | 846 | 0.74 [0.70, 0.79] | 0.72 [0.68, 0.77] |
| 300 ≤ CAC | 2693 | 0.74 [0.71, 0.76] | 0.70 [0.68, 0.72] |
| **By imaging modality** |  |  |  |
| PET (Intermountain Healthcare) | 5611 | 0.76 [0.75, 0.78] | 0.74 [0.73, 0.76] |
| SPECT (University of Ottawa) | 1417 | 0.67 [0.55, 0.79] | 0.67 [0.55, 0.79] |
| **By tube voltage [kV]** |  |  |  |
| 120 | 2931 | 0.78 [0.76, 0.80] | 0.76 [0.73, 0.78] |
| 130 | 4097 | 0.79 [0.77, 0.81] | 0.75 [0.73, 0.77] |
| **By reconstruction diameter [mm]** |  |  |  |
| 500 | 1417 | 0.67 [0.55, 0.79] | 0.67 [0.55, 0.79] |
| 700 | 2680 | 0.76 [0.73, 0.78] | 0.74 [0.71, 0.76] |
| 780 | 2931 | 0.78 [0.76, 0.80] | 0.76 [0.73, 0.78] |
| **By convolutional kernel** |  |  |  |
| I26f | 2931 | 0.78 [0.76, 0.80] | 0.76 [0.73, 0.78] |
| B19s | 2680 | 0.76 [0.73, 0.78] | 0.74 [0.71, 0.76] |
| B30s | 1417 | 0.67 [0.55, 0.79] | 0.67 [0.55, 0.79] |
| **By slice thickness [mm]** |  |  |  |
| 2.0 | 5611 | 0.76 [0.75, 0.78] | 0.74 [0.73, 0.76] |
| 5.0 | 1417 | 0.67 [0.55, 0.79] | 0.67 [0.55, 0.79] |
| **By pixel spacing [mm]** |  |  |  |
| 0.98 | 1417 | 0.67 [0.55, 0.79] | 0.67 [0.55, 0.79] |
| 1.37 | 2680 | 0.76 [0.73, 0.78] | 0.74 [0.71, 0.76] |
| 1.52 | 2931 | 0.78 [0.76, 0.80] | 0.76 [0.73, 0.78] |

Bold indicates statistical significance. Subpopulations stratification by body mass index (BMI), age, stress total perfusion deficit (TPD), left ventricular ejection fraction (LVEF), and coronary artery calcium (CAC) score follows existing literature.^11-16^ The hazard ratio was with respect to per standard deviation (SD) increase in the body composition index (BCI). N – number of patients in each subpopulation, AUC – area under receiver operating characteristic curve, C-index – Harrell concordance index.

**Supplementary Table 7. Unadjusted and adjusted hazard ratio (HR) of body composition index (BCI) as continuous variable for mortality risk stratification in external testing data subpopulations stratified by patient characteristics and imaging acquisition protocols.**

|  | |  | HR [95% CI], p-value | | |
| --- | --- | --- | --- | --- | --- |
| Population | | Metric change | BCI^a^ | BCI^b^ | |
|  |  |  | Unadjusted | Unadjusted | Adjusted |
| Entire | | + SD | **1.79 [1.71, 1.86], <0.0001** | **1.9 [1.82, 1.98], <0.0001** | **1.73 [1.65, 1.81], <0.0001** |
| Subpopulations by Demographics | | | |  |  |
| Sex | Female | + SD | **1.85 [1.72, 1.99], <0.0001** | **1.96 [1.82, 2.1], <0.0001** | **1.76 [1.63, 1.89], <0.0001** |
|  | Male | + SD | **1.77 [1.68, 1.86], <0.0001** | **1.86 [1.77, 1.96], <0.0001** | **1.71 [1.62, 1.81], <0.0001** |
| Age | Age < 60 [year] | + SD | **1.48 [1.35, 1.62], <0.0001** | **1.53 [1.41, 1.65], <0.0001** | **1.38 [1.26, 1.51], <0.0001** |
|  | 60 ≤ Age ≤ 69 [year] | + SD | **1.71 [1.59, 1.85], <0.0001** | **1.74 [1.62, 1.86], <0.0001** | **1.56 [1.44, 1.69], <0.0001** |
|  | 70 ≤ Age [year] | + SD | **1.7 [1.59, 1.81], <0.0001** | **1.84 [1.72, 1.97], <0.0001** | **1.66 [1.54, 1.78], <0.0001** |
| Body mass index | BMI < 30 [kg/m^2^] | + SD | **1.85 [1.74, 1.97], <0.0001** | **1.98 [1.86, 2.11], <0.0001** | **1.76 [1.65, 1.88], <0.0001** |
|  | 30 ≤ BMI < 35 [kg/m^2^] | + SD | **1.7 [1.56, 1.85], <0.0001** | **1.83 [1.68, 1.98], <0.0001** | **1.69 [1.54, 1.85], <0.0001** |
|  | 35 ≤ BMI < 40 [kg/m^2^] | + SD | **1.77 [1.56, 2.01], <0.0001** | **1.79 [1.61, 2], <0.0001** | **1.72 [1.52, 1.94], <0.0001** |
|  | 40 ≤ BMI [kg/m^2^] | + SD | **1.66 [1.46, 1.9], <0.0001** | **1.72 [1.52, 1.94], <0.0001** | **1.57 [1.38, 1.79], <0.0001** |
| Subpopulations by Medical History | | | |  |  |
| Hypertension | With hypertension | + SD | **1.78 [1.7, 1.86], <0.0001** | **1.89 [1.81, 1.98], <0.0001** | **1.75 [1.67, 1.84], <0.0001** |
|  | No hypertension | + SD | **1.65 [1.4, 1.93], <0.0001** | **1.67 [1.44, 1.94], <0.0001** | **1.59 [1.36, 1.87], <0.0001** |
| Diabetes | With diabetes | + SD | **1.66 [1.55, 1.77], <0.0001** | **1.8 [1.69, 1.93], <0.0001** | **1.66 [1.55, 1.78], <0.0001** |
|  | No diabetes | + SD | **1.86 [1.76, 1.97], <0.0001** | **1.94 [1.83, 2.05], <0.0001** | **1.79 [1.69, 1.9], <0.0001** |
| Dyslipidemia | With dyslipidemia | + SD | **1.77 [1.69, 1.85], <0.0001** | **1.89 [1.81, 1.98], <0.0001** | **1.74 [1.66, 1.83], <0.0001** |
|  | No dyslipidemia | + SD | **1.86 [1.67, 2.08], <0.0001** | **1.88 [1.69, 2.08], <0.0001** | **1.71 [1.52, 1.91], <0.0001** |
| Family history of coronary artery disease | With FH of CAD | + SD | **1.73 [1.63, 1.84], <0.0001** | **1.85 [1.75, 1.96], <0.0001** | **1.7 [1.6, 1.81], <0.0001** |
|  | No FH of CAD | + SD | **1.83 [1.73, 1.95], <0.0001** | **1.94 [1.82, 2.06], <0.0001** | **1.76 [1.65, 1.88], <0.0001** |
| Smoking | Smoking | + SD | **1.75 [1.63, 1.88], <0.0001** | **1.85 [1.73, 1.98], <0.0001** | **1.75 [1.64, 1.88], <0.0001** |
|  | No smoking | + SD | **1.79 [1.7, 1.89], <0.0001** | **1.91 [1.81, 2.02], <0.0001** | **1.71 [1.61, 1.81], <0.0001** |
| Subpopulations by Myocardial Perfusion Imaging Findings | | | |  |  |
| Stress total perfusion deficit | Stress TPD < 1%b | + SD | **1.7 [1.5, 1.92], <0.0001** | **1.8 [1.61, 2.03], <0.0001** | **1.7 [1.5, 1.92], <0.0001** |
|  | 1% ≤ Stress TPD < 5% | + SD | **1.82 [1.69, 1.95], <0.0001** | **1.94 [1.8, 2.09], <0.0001** | **1.84 [1.71, 1.99], <0.0001** |
|  | 5% ≤ Stress TPD < 10% | + SD | **1.83 [1.67, 2.01], <0.0001** | **1.88 [1.72, 2.06], <0.0001** | **1.73 [1.57, 1.9], <0.0001** |
|  | Stress TPD ≥ 10% | + SD | **1.75 [1.62, 1.88], <0.0001** | **1.87 [1.73, 2.02], <0.0001** | **1.65 [1.52, 1.79], <0.0001** |
| Left ventricle ejection fraction | LVEF < 50% | + SD | **1.57 [1.42, 1.73], <0.0001** | **1.65 [1.5, 1.83], <0.0001** | **1.6 [1.44, 1.77], <0.0001** |
|  | 50% ≤ LVEF ≤ 70% | + SD | **1.75 [1.63, 1.88], <0.0001** | **1.86 [1.73, 1.99], <0.0001** | **1.76 [1.64, 1.9], <0.0001** |
|  | LVEF > 70% | + SD | **1.8 [1.69, 1.93], <0.0001** | **1.88 [1.76, 2], <0.0001** | **1.8 [1.69, 1.93], <0.0001** |
| Subpopulations by Coronary Artery Calcium Score | | | |  |  |
| Coronary artery calcium score | CAC 0 [AU] | + SD | **1.65 [1.49, 1.83], <0.0001** | **1.55 [1.42, 1.69], <0.0001** | **1.46 [1.33, 1.61], <0.0001** |
|  | 1 ≤ CAC ≤ 99 [AU] | + SD | **1.57 [1.38, 1.8], <0.0001** | **1.63 [1.43, 1.84], <0.0001** | **1.53 [1.35, 1.74], <0.0001** |
|  | 100 ≤ CAC ≤ 299 [AU] | + SD | **1.79 [1.6, 2.01], <0.0001** | **1.97 [1.74, 2.24], <0.0001** | **1.88 [1.65, 2.13], <0.0001** |
|  | 300 ≤ CAC [AU] | + SD | **1.73 [1.63, 1.84], <0.0001** | **1.82 [1.7, 1.94], <0.0001** | **1.7 [1.59, 1.81], <0.0001** |
| Subpopulations by Imaging Modality and Reconstruction Parameters | | | |  |  |
| Modality (Site, breathing mode) | PET (Intermountain Healthcare, normal breath) | + SD | **1.81 [1.73, 1.89], <0.0001** | **1.92 [1.84, 2.01], <0.0001** | **1.78 [1.69, 1.86], <0.0001** |
|  | SPECT (University of Ottawa, breath hold) | + SD | **1.43 [1.12, 1.82], 0.0038** | **1.61 [1.3, 2], <0.0001** | **1.46 [1.15, 1.84], 0.0017** |
| Reconstruction diameter | Reconstruction diameter 500 [mm] | + SD | **1.43 [1.12, 1.82], 0.0038** | **1.61 [1.3, 2], <0.0001** | **1.46 [1.15, 1.84], 0.0017** |
|  | Reconstruction diameter 700 [mm] | + SD | **1.85 [1.73, 1.98], <0.0001** | **1.97 [1.84, 2.12], <0.0001** | **1.81 [1.68, 1.95], <0.0001** |
|  | Reconstruction diameter 780 [mm] | + SD | **1.86 [1.75, 1.98], <0.0001** | **1.97 [1.86, 2.08], <0.0001** | **1.84 [1.73, 1.96], <0.0001** |
| Tube voltage | Tube voltage 120 [kV] | + SD | **1.86 [1.75, 1.98], <0.0001** | **1.97 [1.86, 2.08], <0.0001** | **1.84 [1.73, 1.96], <0.0001** |
|  | Tube voltage 130 [kV] | + SD | **1.81 [1.71, 1.93], <0.0001** | **1.93 [1.81, 2.05], <0.0001** | **1.72 [1.61, 1.84], <0.0001** |
| Kernel | I26f kernel | + SD | **1.86 [1.75, 1.98], <0.0001** | **1.97 [1.86, 2.08], <0.0001** | **1.84 [1.73, 1.96], <0.0001** |
|  | B19s kernel | + SD | **1.85 [1.73, 1.98], <0.0001** | **1.97 [1.84, 2.12], <0.0001** | **1.81 [1.68, 1.95], <0.0001** |
|  | B30s kernel | + SD | **1.43 [1.12, 1.82], 0.0038** | **1.61 [1.3, 2], <0.0001** | **1.46 [1.15, 1.84], 0.0017** |
| Subpopulations by Imaging Resolution Parameters | | | |  |  |
| Pixel spacing | Pixel spacing 0.98 [mm] | + SD | **1.43 [1.12, 1.82], 0.0038** | **1.61 [1.3, 2], <0.0001** | **1.46 [1.15, 1.84], 0.0017** |
|  | Pixel spacing 1.37 [mm] | + SD | **1.85 [1.73, 1.98], <0.0001** | **1.97 [1.84, 2.12], <0.0001** | **1.81 [1.68, 1.95], <0.0001** |
|  | Pixel spacing 1.52 [mm] | + SD | **1.86 [1.75, 1.98], <0.0001** | **1.97 [1.86, 2.08], <0.0001** | **1.84 [1.73, 1.96], <0.0001** |
| Slice thickness | Slice thickness 2 [mm] | + SD | **1.81 [1.73, 1.89], <0.0001** | **1.92 [1.84, 2.01], <0.0001** | **1.78 [1.69, 1.86], <0.0001** |
|  | Slice thickness 5 [mm] | + SD | **1.43 [1.12, 1.82], 0.0038** | **1.61 [1.3, 2], <0.0001** | **1.46 [1.15, 1.84], 0.0017** |

Bold indicates statistical significance. ^a^ indicates that the BCI was developed using only body composition metrics. ^b^ indicates that the BCI was developed using body composition metrics, demographics, and CT-derived coronary artery calcium (CAC) score. Subpopulations stratification by body mass index (BMI), age, stress total perfusion deficit (TPD), left ventricular ejection fraction (LVEF), and CAC score follows existing literature.^11-16^ Medical histories and myocardial perfusion imaging variables were used for adjustment. The hazard ratio was with respect to per standard deviation (SD) increase in the body composition index (BCI). FH – family history, CAD – coronary artery disease, AU – Agatston unit

**Supplementary Table 8. Characteristics of external testing cohort patients stratified by body composition index (BCI) with cutoffs corresponding to negative predictive value 98% and positive predictive value 40% derived from development cohort.**

| Characteristics | Overall  N = 7028 | High-risk  (BCI > 0.29)  N=921 | Intermediate-risk  0.29 ≥ BCI ≥ 0.04  N=4172 | Low-risk  (BCI < 0.04)  N=1935 | p-value |
| --- | --- | --- | --- | --- | --- |
| **Coronary Artery Calcium Score and Demographics** | | | | | |
| Age | 66 (57, 74) | 77 (69, 83) | 68 (60, 74) | 57 (50, 64) | **<0.0001** |
| Sex |  |  |  |  | 0.0658 |
| Female | 3069 (44%) | 414 (45%) | 1853 (44%) | 802 (41%) |  |
| Male | 3959 (56%) | 507 (55%) | 2319 (56%) | 1133 (59%) |  |
| BMI | 30.8 (26.6, 35.6) | 28.2 (24.8, 32.1) | 30.2 (26.2, 34.9) | 33.0 (29.2, 37.8) | **<0.0001** |
| log10(CAC score+1) | 2.02 (0.00, 2.93) | 2.89 (2.31, 3.31) | 2.20 (0.00, 2.99) | 0.00 (0.00, 1.99) | **<0.0001** |
| **Body Composition Metrics** | | | | | |
| VAT volume index | 450 (317, 612) | 403 (285, 580) | 440 (308, 602) | 489 (364, 646) | **<0.0001** |
| VAT attenuation | -82 (-86, -76) | -74 (-80, -68) | -81 (-85, -76) | -86 (-89, -82) | **<0.0001** |
| VAT standard deviation | 63 (57, 69) | 61 (57, 67) | 63 (57, 70) | 63 (55, 68) | **<0.0001** |
| EAT volume index | 37 (25, 51) | 40 (29, 56) | 38 (25, 52) | 34 (24, 46) | **<0.0001** |
| EAT attenuation | -67.7 (-71.2, -64.3) | -67.6 (-71.4, -63.3) | -67.9 (-71.5, -64.3) | -67.4 (-70.6, -64.4) | **0.0004** |
| EAT standard deviation | 43 (40, 48) | 43 (39, 48) | 43 (40, 48) | 44 (41, 48) | **<0.0001** |
| Bone volume index | 302 (266, 345) | 285 (249, 323) | 300 (265, 343) | 316 (279, 357) | **<0.0001** |
| Bone attenuation | 257 (229, 285) | 223 (201, 252) | 252 (227, 279) | 279 (255, 301) | **<0.0001** |
| Bone standard deviation | 207 (186, 229) | 191 (170, 216) | 205 (184, 227) | 217 (198, 235) | **<0.0001** |
| SM volume index | 736 (593, 908) | 542 (450, 652) | 710 (589, 851) | 918 (758, 1079) | **<0.0001** |
| SM attenuation | 22 (18, 27) | 16 (13, 19) | 22 (18, 26) | 27 (22, 31) | **<0.0001** |
| SM standard deviation | 47 (42, 52) | 47 (44, 53) | 47 (42, 52) | 47 (41, 52) | **<0.0001** |
| IMAT volume index | 77 (55, 105) | 78 (55, 109) | 76 (54, 106) | 77 (56, 102) | 0.1332 |
| IMAT attenuation | -69.2 (-72.9, -65.7) | -64.4 (-67.6, -61.8) | -68.6 (-71.8, -65.6) | -72.8 (-75.7, -69.7) | **<0.0001** |
| IMAT standard deviation | 44 (38, 51) | 42 (37, 48) | 44 (38, 51) | 46 (39, 53) | **<0.0001** |
| SAT volume index | 1248 (742, 1893) | 1008 (597, 1617) | 1225 (714, 1880) | 1408 (894, 2035) | **<0.0001** |
| SAT attenuation | -100 (-104, -95) | -96 (-101, -89) | -100 (-104, -95) | -101 (-105, -98) | **<0.0001** |
| SAT standard deviation | 32.8 (29.4, 36.7) | 32.4 (29.5, 36.0) | 32.5 (29.1, 36.4) | 33.6 (30.0, 37.4) | **<0.0001** |
| Ratio SM2AT | 0.39 (0.27, 0.61) | 0.34 (0.23, 0.51) | 0.38 (0.26, 0.60) | 0.45 (0.31, 0.65) | **<0.0001** |
| Ratio ECTOPIC2AT | 0.33 (0.23, 0.43) | 0.35 (0.26, 0.47) | 0.33 (0.23, 0.44) | 0.31 (0.21, 0.41) | **<0.0001** |
| **Prediction** | | | | | |
| Body composition index | 0.08 (0.04, 0.18) | 0.40 (0.34, 0.49) | 0.10 (0.07, 0.17) | 0.03 (0.02, 0.03) | **<0.0001** |
| **Clinical Outcome** | | | | | |
| Death | 949 (14%) | 364 (40%) | 537 (13%) | 48 (2.5%) | **<0.0001** |
| Death follow up | 4.36 (2.30, 4.89) | 3.77 (1.71, 4.78) | 4.45 (2.49, 4.93) | 4.19 (2.16, 4.85) | **<0.0001** |

Bold indicates statistical significance. BCI integrated all body composition metrics, coronary artery calcium score, and demographics variable modifiers. Cutoffs 0.0502 and 0.3254 corresponding to 98% negative predictive value and 40% positive predictive values, respectively were used to categorize patients. CAD – coronary artery calcium score, PET – positron emission tomography, SPECT – single photon emission computed tomography, VAT – visceral adipose tissue, EAT – epicardial adipose tissue, SM – skeletal muscle, IMAT – intramuscular adipose tissue, SAT – subcutaneous adipose tissue, ratio SM2AT – ratio of skeletal muscle volume to adipose tissue (including SAT, IMAT, EAT, and VAT) volume, ratio ECTOPIC2AT – ratio of ectopic adipose tissue (including IMAT, EAT, and VAT) volume to adipose tissue.

**Supplementary Table 9. Patient characteristics in different prediction pattern groups when the Youden index cutoff 0.1368 and the observed death event were used for the body composition index (BCI) to categorize patients into false negative, false positive, true negative, and false negative.**

| Characteristics | Overall N = 7028 | False negative  N = 273 | False positive  N = 1679 | True negative  N = 4400 | True positive  N=676 |
| --- | --- | --- | --- | --- | --- |
| **Demographics and Coronary Artery Calcium Score** | | | | | |
| Age | 66 (57, 74) | 65 (57, 71) | 73 (66, 79) | 62 (54, 69) | 76 (68, 83) |
| Sex |  |  |  |  |  |
| Female | 3069 (44%) | 99 (36%) | 789 (47%) | 1908 (43%) | 273 (40%) |
| Male | 3959 (56%) | 174 (64%) | 890 (53%) | 2492 (57%) | 403 (60%) |
| BMI | 30.8 (26.6, 35.6) | 32.3 (27.1, 37.9) | 28.7 (25.1, 33.0) | 32.0 (27.8, 36.6) | 28.5 (24.6, 32.9) |
| log10(CAC score+1) | 2.02 (0.00, 2.93) | 2.26 (0.00, 2.99) | 2.67 (1.90, 3.17) | 1.22 (0.00, 2.54) | 2.90 (2.27, 3.34) |
| **Body Composition Features** | | | | | |
| VAT volume index | 450 (317, 612) | 520 (407, 681) | 406 (284, 567) | 466 (333, 627) | 428 (304, 593) |
| VAT attenuation | -82 (-86, -76) | -82 (-86, -78) | -78 (-82, -71) | -84 (-87, -79) | -75 (-81, -68) |
| VAT standard deviation | 63 (57, 69) | 65 (61, 70) | 62 (57, 68) | 63 (56, 69) | 63 (58, 70) |
| EAT volume index | 37 (25, 51) | 39 (26, 53) | 39 (27, 55) | 36 (24, 49) | 40 (27, 54) |
| EAT attenuation | -67.7 (-71.2, -64.3) | -68.2 (-71.6, -64.8) | -68.2 (-72.1, -64.2) | -67.6 (-71.0, -64.4) | -67.1 (-70.9, -62.8) |
| EAT standard deviation | 43 (40, 48) | 46 (42, 50) | 42 (39, 47) | 43 (40, 47) | 44 (40, 50) |
| Bone volume index | 302 (266, 345) | 326 (284, 363) | 287 (252, 329) | 307 (272, 349) | 298 (263, 343) |
| Bone attenuation | 257 (229, 285) | 266 (241, 294) | 234 (211, 262) | 266 (241, 292) | 235 (209, 264) |
| Bone standard deviation | 207 (186, 229) | 219 (197, 241) | 194 (174, 218) | 211 (191, 232) | 203 (179, 228) |
| SM volume index | 736 (593, 908) | 807 (665, 967) | 595 (494, 717) | 808 (669, 974) | 598 (486, 738) |
| SM attenuation | 22 (18, 27) | 22 (18, 27) | 19 (15, 22) | 25 (20, 29) | 17 (13, 21) |
| SM standard deviation | 47 (42, 52) | 51 (46, 55) | 46 (42, 51) | 47 (41, 52) | 49 (45, 54) |
| IMAT volume index | 77 (55, 105) | 83 (64, 111) | 76 (53, 104) | 76 (54, 105) | 80 (59, 111) |
| IMAT attenuation | -69.2 (-72.9, -65.7) | -68.8 (-72.0, -65.6) | -66.5 (-69.4, -63.5) | -70.9 (-74.3, -67.8) | -64.6 (-67.8, -62.0) |
| IMAT standard deviation | 44 (38, 51) | 49 (43, 55) | 42 (36, 48) | 45 (38, 52) | 44 (39, 50) |
| SAT volume index | 1248 (742, 1893) | 1305 (809, 2175) | 1099 (642, 1718) | 1342 (824, 1970) | 969 (551, 1582) |
| SAT attenuation | -100 (-104, -95) | -100 (-104, -95) | -98 (-102, -93) | -101 (-104, -97) | -95 (-101, -89) |
| SAT standard deviation | 32.8 (29.4, 36.7) | 36.0 (31.8, 39.4) | 31.7 (28.6, 35.3) | 32.9 (29.5, 36.9) | 33.1 (30.0, 37.2) |
| Ratio SM2AT | 0.39 (0.27, 0.61) | 0.41 (0.26, 0.65) | 0.35 (0.24, 0.55) | 0.42 (0.28, 0.62) | 0.37 (0.25, 0.59) |
| Ratio ECTOPIC2AT | 0.33 (0.23, 0.43) | 0.36 (0.24, 0.47) | 0.32 (0.24, 0.44) | 0.32 (0.22, 0.42) | 0.37 (0.27, 0.49) |
| **Predictions** | | | | | |
| BCI | 0.08 (0.04, 0.18) | 0.07 (0.05, 0.10) | 0.23 (0.18, 0.33) | 0.05 (0.03, 0.08) | 0.31 (0.21, 0.44) |
| **Truth Outcomes** | | | | | |
| Death | 949 (14%) | 273 (100%) | 0 (0%) | 0 (0%) | 676 (100%) |
| Death follow-up | 4.36 (2.30, 4.89) | 2.40 (1.23, 3.54) | 4.61 (3.36, 4.99) | 4.44 (2.43, 4.93) | 2.39 (1.05, 3.45) |

Bold indicates statistical significance. BCI integrated all body composition metrics, coronary artery calcium score, and demographical variable modifiers. Patients with death event during follow up had positive labels while others had negative labels. Patients with BCI lower than 0.1368 were predicted as positive while others were predicted as negative. Youden index cutoff achieved a sensitivity of 0.71 and specificity of 0.72. BMI – body mass index, CAC – coronary artery calcium, SM2AT – ratio of skeletal muscle (SM) volume to adipose tissue volume, ECTOPIC2AT – ratio of ectopic adipose tissue volume to adipose tissue volume, SD – standard deviation, EAT – epicardial adipose tissue, IMAT – intramuscular adipose tissue, SAT – subcutaneous adipose tissue, VAT – visceral adipose tissue.

**Supplementary Table 10: Unadjusted hazard ratio (HR) of body composition index (BCI) using negative predictive value and positive predictive value-based cutoffs for ternary categorization for mortality risk stratification in different subpopulations of external testing cohort.**

| Population | | BCI model 4 risk group | HR [95% CI], p-value |
| --- | --- | --- | --- |
| Entire  N=7028 | | High | **17.11 [12.66, 23.12], <0.0001** |
|  |  | Intermediate | **4.89 [3.64, 6.57], <0.0001** |
| Subpopulations by demographics | | | |
| Sex | Female  N=3069 | High | **19.4 [11.59, 32.48], <0.0001** |
|  |  | Intermediate | **5.25 [3.16, 8.73], <0.0001** |
|  | Male  N=3959 | High | **16.11 [11.11, 23.36], <0.0001** |
|  |  | Intermediate | **4.76 [3.31, 6.84], <0.0001** |
| Age | Age < 60 [year]  N=2145 | High | **9.19 [5.34, 15.84], <0.0001** |
|  |  | Intermediate | **3.13 [2.11, 4.63], <0.0001** |
|  | 60 ≤ Age ≤ 69 [year], N=2258 | High | **23.86 [12.89, 44.17], <0.0001** |
|  |  | Intermediate | **5.54 [3.08, 9.96], <0.0001** |
|  | 70 ≤ Age [year]  N=2625 | High | **36.55 [9.09, 146.87], <0.0001** |
|  |  | Intermediate | **12.57 [3.13, 50.5], 0.0004** |
| Body mass index | BMI < 30 [kg/m^2^]  N=3181 | High | **25.34 [13.46, 47.73], <0.0001** |
|  |  | Intermediate | **7.21 [3.83, 13.55], <0.0001** |
|  | 30 ≤ BMI < 35 [kg/m^2^]  N=1940 | High | **12.54 [7.51, 20.92], <0.0001** |
|  |  | Intermediate | **3.95 [2.41, 6.45], <0.0001** |
|  | 35 ≤ BMI < 40 [kg/m^2^]  N=1126 | High | **20.46 [9.81, 42.68], <0.0001** |
|  |  | Intermediate | **5.09 [2.55, 10.17], <0.0001** |
|  | 40 ≤ BMI [kg/m^2^]  N=781 | High | **14.74 [7.01, 31.01], <0.0001** |
|  |  | Intermediate | **4.09 [2.14, 7.81], <0.0001** |
| Subpopulations by medical history | | | |
| Hypertension | With hypertension  N=5848 | High | **15.57 [11.27, 21.52], <0.0001** |
|  |  | Intermediate | **4.73 [3.44, 6.51], <0.0001** |
|  | No hypertension  N=1180 | High | **18.53 [7.39, 46.46], <0.0001** |
|  |  | Intermediate | **3.38 [1.48, 7.72], 0.0038** |
| Diabetes | With diabetes  N=2271 | High | **21.37 [11.61, 39.36], <0.0001** |
|  |  | Intermediate | **7.28 [3.98, 13.31], <0.0001** |
|  | No diabetes  N=4757 | High | **14.96 [10.54, 21.25], <0.0001** |
|  |  | Intermediate | **3.81 [2.7, 5.36], <0.0001** |
| Dyslipidemia | With dyslipidemia  N=5462 | High | **17.67 [12.41, 25.17], <0.0001** |
|  |  | Intermediate | **5.25 [3.7, 7.43], <0.0001** |
|  | No dyslipidemia  N=1566 | High | **15.3 [8.42, 27.81], <0.0001** |
|  |  | Intermediate | **3.53 [1.98, 6.27], <0.0001** |
| Family history (FH) of coronary artery disease (CAD) | With FH of CAD  N=3389 | High | **16.13 [10.74, 24.21], <0.0001** |
|  |  | Intermediate | **4.59 [3.09, 6.82], <0.0001** |
|  | No FH of CAD  N=3639 | High | **18.19 [11.59, 28.53], <0.0001** |
|  |  | Intermediate | **5.26 [3.37, 8.2], <0.0001** |
| Smoking | Smoking  N=2442 | High | **11.65 [7.64, 17.77], <0.0001** |
|  |  | Intermediate | **3.66 [2.43, 5.53], <0.0001** |
|  | No smoking  N=4586 | High | **22.3 [14.49, 34.31], <0.0001** |
|  |  | Intermediate | **5.99 [3.92, 9.16], <0.0001** |
| Subpopulation by myocardial perfusion imaging findings and coronary artery calcium score | | | |
| Stress total perfusion deficit (TPD) | Stress TPD < 1%  N=996 | High | **12.18 [6.02, 24.67], <0.0001** |
|  |  | Intermediate | **3.05 [1.56, 5.96], 0.0011** |
|  | 1% ≤ Stress TPD < 5%  N=2612 | High | **20.31 [11.66, 35.37], <0.0001** |
|  |  | Intermediate | **6.47 [3.77, 11.11], <0.0001** |
|  | 5% ≤ Stress TPD < 10%  N=1500 | High | **18.38 [9.25, 36.55], <0.0001** |
|  |  | Intermediate | **5.07 [2.57, 10], <0.0001** |
|  | Stress TPD ≥ 10%  N=1920 | High | **14.7 [8.61, 25.09], <0.0001** |
|  |  | Intermediate | **4.37 [2.57, 7.43], <0.0001** |
| Left ventricle ejection fraction (LVEF) | LVEF < 50%  N=863 | High | **6.42 [3.25, 12.67], <0.0001** |
|  |  | Intermediate | **3.14 [1.6, 6.15], 0.0009** |
|  | 50% ≤ LVEF ≤ 70%  N=2351 | High | **15.55 [9.39, 25.77], <0.0001** |
|  |  | Intermediate | **4.48 [2.73, 7.36], <0.0001** |
|  | LVEF > 70%  N=3814 | High | **17.53 [11.23, 27.37], <0.0001** |
|  |  | Intermediate | **4.68 [3.04, 7.19], <0.0001** |
| Subpopulation by coronary artery calcium score | | | |
| Coronary artery calcium (CAC) score | CAC 0 [AU]  N=2244 | High | **11.31 [5.73, 22.33], <0.0001** |
|  |  | Intermediate | **3.05 [1.92, 4.86], <0.0001** |
|  | 1 ≤ CAC ≤ 99 [AU]  N=1245 | High | **7.61 [3.69, 15.69], <0.0001** |
|  |  | Intermediate | **3.27 [1.74, 6.14], 0.0002** |
|  | 100 ≤ CAC ≤ 299 [AU]  N=846 | High | **15.81 [6.33, 39.44], <0.0001** |
|  |  | Intermediate | **3.89 [1.57, 9.6], 0.0033** |
|  | 300 ≤ CAC [AU]  N=2693 | High | **18.27 [9.4, 35.49], <0.0001** |
|  |  | Intermediate | **6.63 [3.42, 12.87], <0.0001** |
| Subpopulation by imaging protocols | | | |
| Myocardial perfusion imaging (MPI) modality, Site, Breathing mode | PET/CT, Intermountain Healthcare, normal breath  N=5611 | High | **17.41 [12.6, 24.06], <0.0001** |
|  |  | Intermediate | **5.14 [3.74, 7.07], <0.0001** |
|  | SPECT/CT, University of Ottawa, breath hold  N=1417 | High | **7.2 [2.53, 20.55], 0.0002** |
|  |  | Intermediate | 1.6 [0.65, 3.97], 0.3086 |
| KVP | KVP 120  N=2931 | High | **20.18 [13.59, 29.96], <0.0001** |
|  |  | Intermediate | **5.18 [3.54, 7.59], <0.0001** |
|  | KVP 130  N=4097 | High | **16.56 [10.34, 26.52], <0.0001** |
|  |  | Intermediate | **4.68 [2.93, 7.47], <0.0001** |
| Reconstruction diameter | Diameter 500 [mm]  N=1417 | High | **7.2 [2.53, 20.55], 0.0002** |
|  |  | Intermediate | 1.6 [0.65, 3.97], 0.3086 |
|  | Diameter 700 [mm]  N=2680 | High | **18.46 [10.3, 33.08], <0.0001** |
|  |  | Intermediate | **5.59 [3.13, 10], <0.0001** |
|  | Diameter 780 [mm]  N=2931 | High | **20.18 [13.59, 29.96], <0.0001** |
|  |  | Intermediate | **5.18 [3.54, 7.59], <0.0001** |
| Kernel | I26f kernel  N=2931 | High | **20.18 [13.59, 29.96], <0.0001** |
|  |  | Intermediate | **5.18 [3.54, 7.59], <0.0001** |
|  | B19s kernel  N=2680 | High | **18.46 [10.3, 33.08], <0.0001** |
|  |  | Intermediate | **5.59 [3.13, 10], <0.0001** |
|  | B30s kernel  N=2417 | High | **7.2 [2.53, 20.55], 0.0002** |
|  |  | Intermediate | 1.6 [0.65, 3.97], 0.3086 |
| Slice thickness | Slice thickness 2 [mm]  N=5611 | High | **17.41 [12.6, 24.06], <0.0001** |
|  |  | Intermediate | **5.14 [3.74, 7.07], <0.0001** |
|  | Slice thickness 5 [mm]  N=1417 | High | **7.2 [2.53, 20.55], 0.0002** |
|  |  | Intermediate | 1.6 [0.65, 3.97], 0.3086 |
| Pixel spacing | Pixel spacing 0.98 [mm]  N=1417 | High | **7.2 [2.53, 20.55], 0.0002** |
|  |  | Intermediate | 1.6 [0.65, 3.97], 0.3086 |
|  | Pixel spacing 1.37 [mm]  N=2680 | High | **18.46 [10.3, 33.08], <0.0001** |
|  |  | Intermediate | **5.59 [3.13, 10], <0.0001** |
|  | Pixel spacing 1.52 [mm]  N=2931 | High | **20.18 [13.59, 29.96], <0.0001** |
|  |  | Intermediate | **5.18 [3.54, 7.59], <0.0001** |

Bold indicates statistical significance. Cutoffs corresponding to negative predictive value 98% and positive predictive value 40% were used. BCI integrated body composition metric, coronary artery calcium score, and demographics) Subpopulations stratification by body mass index (BMI), age, stress total perfusion deficit (TPD), left ventricular ejection fraction (LVEF), coronary artery calcium (CAC) score follows existing literature.^11-16^ The patients with low body composition index value was reference group. FH – family history, CAD – coronary artery disease, AU – Agatston unit

**Supplementary Table 11. Predictive value of body composition index (BCI) in external testing cohort.**

| Population | BCI cutoff 0.04 corresponding to NPV 0.98 | | BCI cutoff 0.29 corresponding to PPV 0.40 | |
| --- | --- | --- | --- | --- |
|  | Negative predictive value | Sensitivity | Positive predictive value | Specificity |
| Entire | 0.98 [0.97, 0.98] | 0.95 [0.93, 0.96] | 0.40 [0.36, 0.43] | 0.91 [0.90, 0.92] |
| **By demographics** | | | | |
| Female | 0.98 [0.97, 0.99] | 0.96 [0.93, 0.98] | 0.36 [0.32, 0.41] | 0.90 [0.89, 0.91] |
| Male | 0.97 [0.96, 0.98] | 0.94 [0.92, 0.96] | 0.42 [0.38, 0.46] | 0.91 [0.90, 0.92] |
| Age < 60 [year] | 0.97 [0.96, 0.98] | 0.77 [0.69, 0.83] | 0.28 [0.19, 0.40] | 0.97 [0.97, 0.98] |
| 60 ≤ Age ≤ 69 [year] | 0.98 [0.97, 0.99] | 0.95 [0.91, 0.97] | 0.41 [0.33, 0.49] | 0.95 [0.94, 0.96] |
| 70 ≤ Age [year] | 0.99 [0.96, 1.00] | 1.00 [0.99, 1.00] | 0.40 [0.37, 0.44] | 0.80 [0.78, 0.82] |
| BMI < 30 [kg/m^2^] | 0.98 [0.97, 0.99] | 0.98 [0.96, 0.99] | 0.40 [0.36, 0.45] | 0.87 [0.86, 0.88] |
| 30 ≤ BMI < 35 [kg/m^2^] | 0.97 [0.95, 0.98] | 0.92 [0.88, 0.95] | 0.36 [0.30, 0.43] | 0.92 [0.90, 0.93] |
| 35 ≤ BMI < 40 [kg/m^2^] | 0.98 [0.96, 0.99] | 0.92 [0.86, 0.96] | 0.41 [0.31, 0.53] | 0.95 [0.94, 0.96] |
| 40 ≤ BMI [kg/m^2^] | 0.97 [0.94, 0.98] | 0.87 [0.78, 0.93] | 0.41 [0.27, 0.57] | 0.96 [0.94, 0.97] |
| **By medical history** | | | | |
| No hypertension | 0.99 [0.97, 0.99] | 0.86 [0.73, 0.94] | 0.23 [0.13, 0.36] | 0.96 [0.95, 0.97] |
| With hypertension | 0.97 [0.96, 0.98] | 0.95 [0.94, 0.97] | 0.41 [0.37, 0.44] | 0.90 [0.89, 0.90] |
| No diabetes | 0.97 [0.97, 0.98] | 0.93 [0.90, 0.95] | 0.36 [0.32, 0.40] | 0.92 [0.91, 0.92] |
| With diabetes | 0.98 [0.96, 0.99] | 0.97 [0.95, 0.99] | 0.44 [0.39, 0.50] | 0.89 [0.87, 0.90] |
| No dyslipidemia | 0.98 [0.96, 0.99] | 0.89 [0.83, 0.94] | 0.32 [0.24, 0.40] | 0.93 [0.92, 0.94] |
| With dyslipidemia | 0.97 [0.96, 0.98] | 0.96 [0.94, 0.97] | 0.41 [0.38, 0.45] | 0.90 [0.89, 0.91] |
| No FH of CAD | 0.98 [0.97, 0.99] | 0.96 [0.93, 0.97] | 0.38 [0.34, 0.42] | 0.90 [0.89, 0.91] |
| With FH of CAD | 0.97 [0.96, 0.98] | 0.94 [0.92, 0.96] | 0.41 [0.37, 0.46] | 0.92 [0.91, 0.93] |
| No smoking | 0.98 [0.97, 0.99] | 0.96 [0.93, 0.97] | 0.37 [0.33, 0.41] | 0.91 [0.91, 0.92] |
| With smoking | 0.96 [0.94, 0.97] | 0.94 [0.91, 0.96] | 0.43 [0.38, 0.48] | 0.90 [0.88, 0.91] |
| **By myocardial perfusion imaging finding** | | | | |
| Stress TPD < 1% | 0.97 [0.94, 0.98] | 0.90 [0.83, 0.95] | 0.36 [0.27, 0.47] | 0.93 [0.91, 0.95] |
| 1% ≤ Stress TPD < 5% | 0.98 [0.97, 0.99] | 0.96 [0.93, 0.98] | 0.36 [0.31, 0.41] | 0.91 [0.90, 0.92] |
| 5% ≤ Stress TPD < 10% | 0.98 [0.96, 0.99] | 0.96 [0.92, 0.98] | 0.41 [0.34, 0.48] | 0.90 [0.89, 0.92] |
| Stress TPD ≥ 10% | 0.97 [0.95, 0.98] | 0.95 [0.92, 0.97] | 0.44 [0.38, 0.49] | 0.90 [0.88, 0.91] |
| LVEF < 50% | 0.91 [0.84, 0.96] | 0.97 [0.94, 0.98] | 0.47 [0.40, 0.53] | 0.79 [0.75, 0.82] |
| 50% ≤ LVEF ≤ 70% | 0.98 [0.96, 0.99] | 0.95 [0.92, 0.97] | 0.43 [0.37, 0.49] | 0.91 [0.90, 0.93] |
| LVEF > 70% | 0.98 [0.97, 0.99] | 0.94 [0.91, 0.96] | 0.32 [0.28, 0.37] | 0.93 [0.92, 0.93] |
| **By coronary artery calcium score** | | | | |
| CAC 0 [AU] | 0.98 [0.97, 0.99] | 0.80 [0.71, 0.87] | 0.26 [0.15, 0.40] | 0.98 [0.98, 0.99] |
| 1 ≤ CAC ≤ 99 [AU] | 0.97 [0.95, 0.98] | 0.90 [0.83, 0.95] | 0.22 [0.14, 0.31] | 0.93 [0.91, 0.95] |
| 100 ≤ CAC ≤ 299 [AU] | 0.97 [0.92, 0.99] | 0.96 [0.92, 0.99] | 0.44 [0.35, 0.53] | 0.90 [0.87, 0.92] |
| 300 ≤ CAC [AU] | 0.97 [0.95, 0.99] | 0.98 [0.97, 0.99] | 0.42 [0.39, 0.46] | 0.83 [0.81, 0.84] |
| **By imaging acquisition protocol** | | | | |
| PET (Intermountain Healthcare, normal breath) | 0.97 [0.96, 0.98] | 0.96 [0.94, 0.97] | 0.40 [0.36, 0.43] | 0.91 [0.90, 0.92] |
| SPECT (University of Ottawa, breath hold) | 0.99 [0.98, 1.00] | 0.75 [0.55, 0.89] | 0.42 [0.39, 0.46] | 0.90 [0.89, 0.91] |
| KVP 120 | 0.97 [0.95, 0.98] | 0.94 [0.92, 0.96] | 0.09 [0.04, 0.17] | 0.95 [0.94, 0.96] |
| KVP 130 | 0.98 [0.97, 0.99] | 0.96 [0.93, 0.97] | 0.50 [0.44, 0.56] | 0.93 [0.92, 0.94] |
| Reconstruction diameter 500 [mm] | 0.99 [0.98, 1.00] | 0.75 [0.55, 0.89] | 0.34 [0.30, 0.38] | 0.89 [0.88, 0.90] |
| Reconstruction diameter 700 [mm] | 0.98 [0.96, 0.99] | 0.97 [0.95, 0.99] | 0.09 [0.04, 0.17] | 0.95 [0.94, 0.96] |
| Reconstruction diameter 780 [mm] | 0.97 [0.95, 0.98] | 0.94 [0.92, 0.96] | 0.38 [0.33, 0.42] | 0.86 [0.84, 0.87] |
| Kernel_I26f | 0.97 [0.95, 0.98] | 0.94 [0.92, 0.96] | 0.50 [0.44, 0.56] | 0.93 [0.92, 0.94] |
| Kernel_B19s | 0.98 [0.96, 0.99] | 0.97 [0.95, 0.99] | 0.50 [0.44, 0.56] | 0.93 [0.92, 0.94] |
| Kernel_B30s | 0.99 [0.98, 1.00] | 0.75 [0.55, 0.89] | 0.38 [0.33, 0.42] | 0.86 [0.84, 0.87] |
| Slice thickness 2 [mm] | 0.97 [0.96, 0.98] | 0.96 [0.94, 0.97] | 0.09 [0.04, 0.17] | 0.95 [0.94, 0.96] |
| Slice thickness 5 [mm] | 0.99 [0.98, 1.00] | 0.75 [0.55, 0.89] | 0.42 [0.39, 0.46] | 0.90 [0.89, 0.91] |
| Pixel spacing 0.98 [mm] | 0.99 [0.98, 1.00] | 0.75 [0.55, 0.89] | 0.09 [0.04, 0.17] | 0.95 [0.94, 0.96] |
| Pixel spacing 1.37 [mm] | 0.98 [0.96, 0.99] | 0.97 [0.95, 0.99] | 0.09 [0.04, 0.17] | 0.95 [0.94, 0.96] |
| Pixel spacing 1.52 [mm] | 0.97 [0.95, 0.98] | 0.94 [0.92, 0.96] | 0.38 [0.33, 0.42] | 0.86 [0.84, 0.87] |

Bold indicates statistical significance. BCI integrated all body composition metrics, coronary artery calcium, and demographical variable modifiers. Subpopulations stratification by body mass index (BMI), age, stress total perfusion deficit (TPD), left ventricular ejection fraction (LVEF), and coronary artery calcium (CAC) score follows existing literature.^11-16^ The hazard ratio was with respect to per standard deviation (SD) increase in the body composition index (BCI). PET – positron emission tomography, SPECT – single photon emission computed tomography, BMI – body mass index, FH – family history, CAD – coronary artery disease, TPD – total perfusion deficit, LVEF – left ventricular ejection fraction, CAC – coronary artery calcium, AU – Agatston unit, NPV – negative predictive value, PPV – positive predictive value.

**Supplementary Table 12. Discriminative prediction performance of body composition index (BCI) in internal testing subgroups stratified by patient characteristics and imaging acquisition protocols.**

| Population | N | AUC | C-index |
| --- | --- | --- | --- |
| Entire | 6444 | 0.81 [0.80, 0.83] | 0.75 [0.73, 0.76] |
| **By demographics** |  |  |  |
| Female | 2853 | 0.82 [0.80, 0.84] | 0.75 [0.72, 0.78] |
| Male | 3591 | 0.81 [0.79, 0.83] | 0.74 [0.72, 0.76] |
| Age < 60 | 1927 | 0.78 [0.75, 0.82] | 0.73 [0.69, 0.78] |
| 60 ≤ Age ≤ 69 | 1950 | 0.79 [0.76, 0.82] | 0.72 [0.69, 0.76] |
| 70 ≤ Age | 2567 | 0.79 [0.77, 0.81] | 0.71 [0.68, 0.73] |
| BMI < 30 | 3347 | 0.81 [0.80, 0.83] | 0.74 [0.72, 0.76] |
| 30 ≤ BMI < 35 | 1455 | 0.83 [0.80, 0.86] | 0.76 [0.72, 0.80] |
| 35 ≤ BMI < 40 | 814 | 0.81 [0.77, 0.86] | 0.73 [0.67, 0.78] |
| 40 ≤ BMI | 828 | 0.75 [0.69, 0.80] | 0.70 [0.65, 0.76] |
| Black | 916 | 0.80 [0.76, 0.84] | 0.71 [0.67, 0.76] |
| White | 3811 | 0.82 [0.80, 0.83] | 0.75 [0.73, 0.77] |
| **By medical history** |  |  |  |
| No hypertension | 1835 | 0.77 [0.74, 0.81] | 0.73 [0.69, 0.77] |
| With hypertension | 4609 | 0.82 [0.80, 0.83] | 0.74 [0.73, 0.76] |
| No diabetes | 4185 | 0.82 [0.80, 0.84] | 0.74 [0.72, 0.76] |
| With diabetes | 2259 | 0.80 [0.78, 0.82] | 0.74 [0.72, 0.77] |
| No dyslipidemia | 2480 | 0.79 [0.77, 0.82] | 0.74 [0.71, 0.77] |
| With dyslipidemia | 3964 | 0.82 [0.80, 0.83] | 0.74 [0.72, 0.76] |
| No family history of CAD | 4820 | 0.82 [0.80, 0.83] | 0.75 [0.73, 0.77] |
| With family history of CAD | 1624 | 0.80 [0.77, 0.83] | 0.72 [0.69, 0.76] |
| No smoking | 5578 | 0.82 [0.80, 0.83] | 0.75 [0.73, 0.77] |
| With smoking | 866 | 0.79 [0.75, 0.83] | 0.74 [0.69, 0.79] |
| **By myocardial perfusion imaging finding** |  |  |  |
| Stress TPD < 1% | 1252 | 0.81 [0.77, 0.85] | 0.71 [0.66, 0.77] |
| 1% ≤ Stress TPD < 5% | 2458 | 0.81 [0.78, 0.83] | 0.74 [0.71, 0.77] |
| 5% ≤ Stress TPD < 10% | 1232 | 0.80 [0.77, 0.84] | 0.73 [0.70, 0.77] |
| 10% ≤ Stress TPD | 1502 | 0.76 [0.74, 0.79] | 0.70 [0.68, 0.73] |
| LVEF < 50% | 1182 | 0.78 [0.75, 0.80] | 0.69 [0.66, 0.72] |
| 50% ≤ LVEF ≤ 70% | 3117 | 0.82 [0.80, 0.84] | 0.76 [0.73, 0.78] |
| 70% < LVEF | 2145 | 0.81 [0.78, 0.84] | 0.73 [0.69, 0.77] |
| **By coronary artery calcium score** |  |  |  |
| CAC = 0 | 2085 | 0.77 [0.74, 0.81] | 0.68 [0.64, 0.73] |
| 1 ≤ CAC ≤ 99 | 1146 | 0.79 [0.75, 0.83] | 0.68 [0.63, 0.73] |
| 100 ≤ CAC ≤ 299 | 762 | 0.80 [0.76, 0.84] | 0.69 [0.65, 0.74] |
| 300 ≤ CAC | 2451 | 0.81 [0.79, 0.83] | 0.74 [0.72, 0.76] |
| **By imaging acquisition protocol** |  |  |  |
| PET (seven sites) | 3683 | 0.79 [0.77, 0.81] | 0.74 [0.72, 0.76] |
| SPECT (three sites) | 2761 | 0.74 [0.71, 0.78] | 0.74 [0.70, 0.77] |
| KVP 100 | 1375 | 0.77 [0.74, 0.80] | 0.75 [0.72, 0.77] |
| KVP 120 | 4368 | 0.77 [0.75, 0.79] | 0.74 [0.72, 0.77] |
| KVP 140 | 699 | 0.74 [0.71, 0.78] | 0.70 [0.67, 0.73] |
| Reconstruction diameter 500 [mm] | 4250 | 0.84 [0.82, 0.85] | 0.74 [0.72, 0.76] |
| Reconstruction diameter 600 [mm] | 975 | 0.68 [0.61, 0.76] | 0.75 [0.68, 0.81] |
| Reconstruction diameter 700 [mm] | 1053 | 0.72 [0.68, 0.76] | 0.72 [0.68, 0.76] |
| B convolutional kernel | 975 | 0.68 [0.61, 0.76] | 0.75 [0.68, 0.81] |
| SOFT convolutional kernel | 1163 | 0.77 [0.74, 0.80] | 0.71 [0.68, 0.73] |
| STANDARD convolution kernel | 4203 | 0.80 [0.78, 0.82] | 0.75 [0.73, 0.77] |
| Slice thickness 2.5 [mm] | 1235 | 0.81 [0.76, 0.86] | 0.80 [0.75, 0.85] |
| Slice thickness 3.0 [mm] | 2451 | 0.78 [0.75, 0.80] | 0.76 [0.74, 0.79] |
| Slice thickness 3.75 [mm] | 643 | 0.75 [0.69, 0.80] | 0.73 [0.67, 0.78] |
| Slice thickness 5.0 [mm] | 2113 | 0.82 [0.80, 0.84] | 0.72 [0.69, 0.74] |
| Pixel spacing 0.98 [mm] | 2874 | 0.87 [0.85, 0.88] | 0.74 [0.72, 0.76] |
| Pixel spacing 1.17 [mm] | 975 | 0.68 [0.61, 0.76] | 0.75 [0.68, 0.81] |
| Pixel spacing 1.37 [mm] | 2429 | 0.75 [0.73, 0.78] | 0.74 [0.71, 0.76] |

Bold indicates statistical significance. BCI integrated all body composition metrics, coronary artery calcium score, and demographics. Race was not included in BCI. Subpopulations stratification by body mass index (BMI), age, stress total perfusion deficit (TPD), left ventricular ejection fraction (LVEF), and coronary artery calcium (CAC) score follows existing literature.^11-16^ N – number of patients in each subpopulation, AUC – area under receiver operating characteristic curve, C-index – Harrell concordance index.

**Supplementary Table 13. Unadjusted hazard ratio (HR) of body composition index (BCI) as continuous variable for mortality risk stratification in internal testing data subpopulations stratified by patient characteristics and imaging acquisition protocols.**

| Population | | BCI | HR [95% CI], p-value |
| --- | --- | --- | --- |
| Entire | | + SD | **1.97 [1.89, 2.07], <0.0001** |
| Subpopulations by Demographics | | | |
| Sex | Female | + SD | **1.94 [1.8, 2.08], <0.0001** |
|  | Male | + SD | **1.97 [1.85, 2.1], <0.0001** |
| Age | Age < 60 [year] | + SD | **1.71 [1.56, 1.88], <0.0001** |
|  | 60 ≤ Age ≤ 69 [year] | + SD | **1.72 [1.58, 1.87], <0.0001** |
|  | 70 ≤ Age [year] | + SD | **1.9 [1.77, 2.05], <0.0001** |
| Body mass index (BMI) | BMI < 30 [kg/m^2^] | + SD | **2.03 [1.91, 2.16], <0.0001** |
|  | 30 ≤ BMI < 35 [kg/m^2^] | + SD | **1.99 [1.8, 2.19], <0.0001** |
|  | 35 ≤ BMI < 40 [kg/m^2^] | + SD | **1.69 [1.46, 1.95], <0.0001** |
|  | 40 ≤ BMI [kg/m^2^] | + SD | **1.65 [1.44, 1.89], <0.0001** |
| Race | Black | + SD | **1.66 [1.48, 1.88], <0.0001** |
|  | White | + SD | **2.14 [2.01, 2.27], <0.0001** |
| Subpopulations by Medical History | | | |
| Hypertension | With hypertension | + SD | **2 [1.89, 2.11], <0.0001** |
|  | No hypertension | + SD | **1.75 [1.59, 1.92], <0.0001** |
| Diabetes | With diabetes | + SD | **1.95 [1.81, 2.1], <0.0001** |
|  | No diabetes | + SD | **1.96 [1.84, 2.07], <0.0001** |
| Dyslipidemia | With dyslipidemia | + SD | **1.98 [1.87, 2.1], <0.0001** |
|  | No dyslipidemia | + SD | **1.91 [1.76, 2.06], <0.0001** |
| Family history of coronary artery disease | With FH of CAD | + SD | **1.82 [1.66, 2], <0.0001** |
|  | No FH of CAD | + SD | **2 [1.9, 2.11], <0.0001** |
| Smoking | Smoking | + SD | **1.71 [1.51, 1.93], <0.0001** |
|  | No smoking | + SD | **2.02 [1.92, 2.12], <0.0001** |
| Subpopulations by Myocardial Perfusion Imaging Findings | | | |
| Stress total perfusion deficit (TPD) | Stress TPD < 1% | + SD | **1.71 [1.5, 1.95], <0.0001** |
|  | 1% ≤ Stress TPD < 5% | + SD | **1.88 [1.74, 2.03], <0.0001** |
|  | 5% ≤ Stress TPD < 10% | + SD | **1.79 [1.61, 1.99], <0.0001** |
|  | Stress TPD ≥ 10% | + SD | **1.92 [1.77, 2.09], <0.0001** |
| Left ventricle ejection fraction (LVEF) | LVEF < 50% | + SD | **1.85 [1.69, 2.03], <0.0001** |
|  | 50% ≤ LVEF ≤ 70% | + SD | **1.9 [1.77, 2.03], <0.0001** |
|  | LVEF > 70% | + SD | **1.84 [1.68, 2.02], <0.0001** |
| Subpopulations by Coronary Artery Calcium Score | | | |
| Coronary artery calcium (CAC) score | CAC 0 [AU] | + SD | **1.52 [1.39, 1.67], <0.0001** |
|  | 1 ≤ CAC ≤ 99 [AU] | + SD | **1.54 [1.37, 1.75], <0.0001** |
|  | 100 ≤ CAC ≤ 299 [AU] | + SD | **1.7 [1.49, 1.95], <0.0001** |
|  | 300 ≤ CAC [AU] | + SD | **2.1 [1.96, 2.26], <0.0001** |
| Subpopulations by Imaging Modality and Reconstruction Parameters | | | |
| Modality (Site) | PET (seven sites) | + SD | **1.97 [1.89, 2.07], <0.0001** |
|  | SPECT (three sites) | + SD | **2.14 [2.01, 2.27], <0.0001** |
| Reconstruction diameter | Reconstruction diameter 500 [mm] | + SD | **1.57 [1.44, 1.7], <0.0001** |
|  | Reconstruction diameter 600 [mm] | + SD | **2 [1.89, 2.12], <0.0001** |
|  | Reconstruction diameter 700 [mm] | + SD | **1.74 [1.45, 2.07], <0.0001** |
| Tube voltage | Tube voltage 100 [kV] | + SD | **2.05 [1.82, 2.3], <0.0001** |
|  | Tube voltage 120 [kV] | + SD | **2.28 [2.06, 2.52], <0.0001** |
|  | Tube voltage 140 [kV] | + SD | **1.76 [1.66, 1.87], <0.0001** |
| Kernel | B kernel | + SD | **2.07 [1.84, 2.32], <0.0001** |
|  | SOFT kernel | + SD | **1.74 [1.45, 2.07], <0.0001** |
|  | STANDARD kernel | + SD | **2.03 [1.85, 2.24], <0.0001** |
|  | Subpopulations by Imaging Resolution Parameters | | |
| Pixel spacing | Pixel spacing 0.98 [mm] | + SD | **1.8 [1.68, 1.92], <0.0001** |
|  | Pixel spacing 1.17 [mm] | + SD | **1.74 [1.45, 2.07], <0.0001** |
|  | Pixel spacing 1.37 [mm] | + SD | **2.15 [1.99, 2.31], <0.0001** |
| Slice thickness | Slice thickness 2.5 [mm] | + SD | **1.81 [1.62, 2.02], <0.0001** |
|  | Slice thickness 3 [mm] | + SD | **2.19 [2.02, 2.36], <0.0001** |
|  | Slice thickness 3.75 [mm] | + SD | **2.01 [1.7, 2.37], <0.0001** |
|  | Slice thickness 5 [mm] | + SD | **1.93 [1.79, 2.09], <0.0001** |

Bold indicates statistical significance. BCI integrated body composition metrics, coronary artery calcium (CAC) score, and demographical variable modifiers. Race was not integrated in BCI. Subpopulations stratification by body mass index (BMI), age, stress total perfusion deficit (TPD), left ventricular ejection fraction (LVEF), and coronary artery calcium (CAC) score follows existing literature.^11-16^ PET – positron emission tomography, SPECT – single photon emission computed tomography, SD – standard deviation.

**Figures**

**Supplementary Figure 1: Cohort creation flowchart for the internal and external testing scheme.**

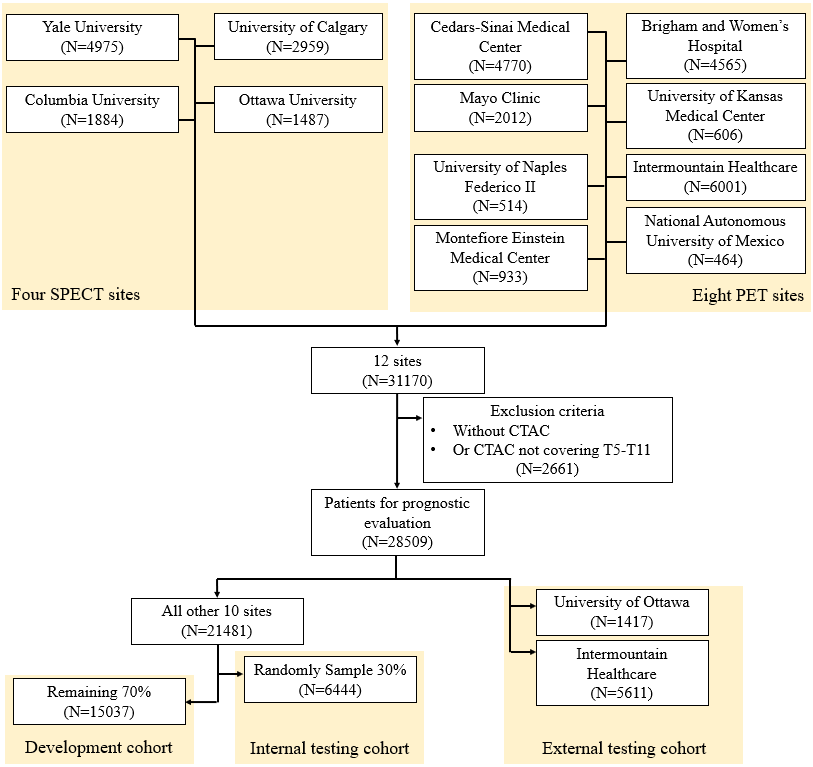

**Supplementary Figure 2. Support vector machine (SVM) hyperparameter tuning, and model training and testing in external testing scheme.** (A): hyperparameter tuning pipeline; (B): model training on development data with the selected hyperparameters; (C): model validation on internal and external testing cohorts with the selected model. NAPLES – University of Naples Federic II, MONTE – Montefiore Einstein Medical Center, BWH – Brigham and Women’s Hospital, CSMC – Cedars-Sinai Medical Center, CALGARY – University of Calgary, COLUMBA – Columbia University, MAYO – Mayo Clinic, KUMC – University of Kansas Medical Center, MXC – National Autonomous University of Mexico, ITMT – Intermountain Healthcare, OTTAWA – University of Ottawa, YALE – Yale University.

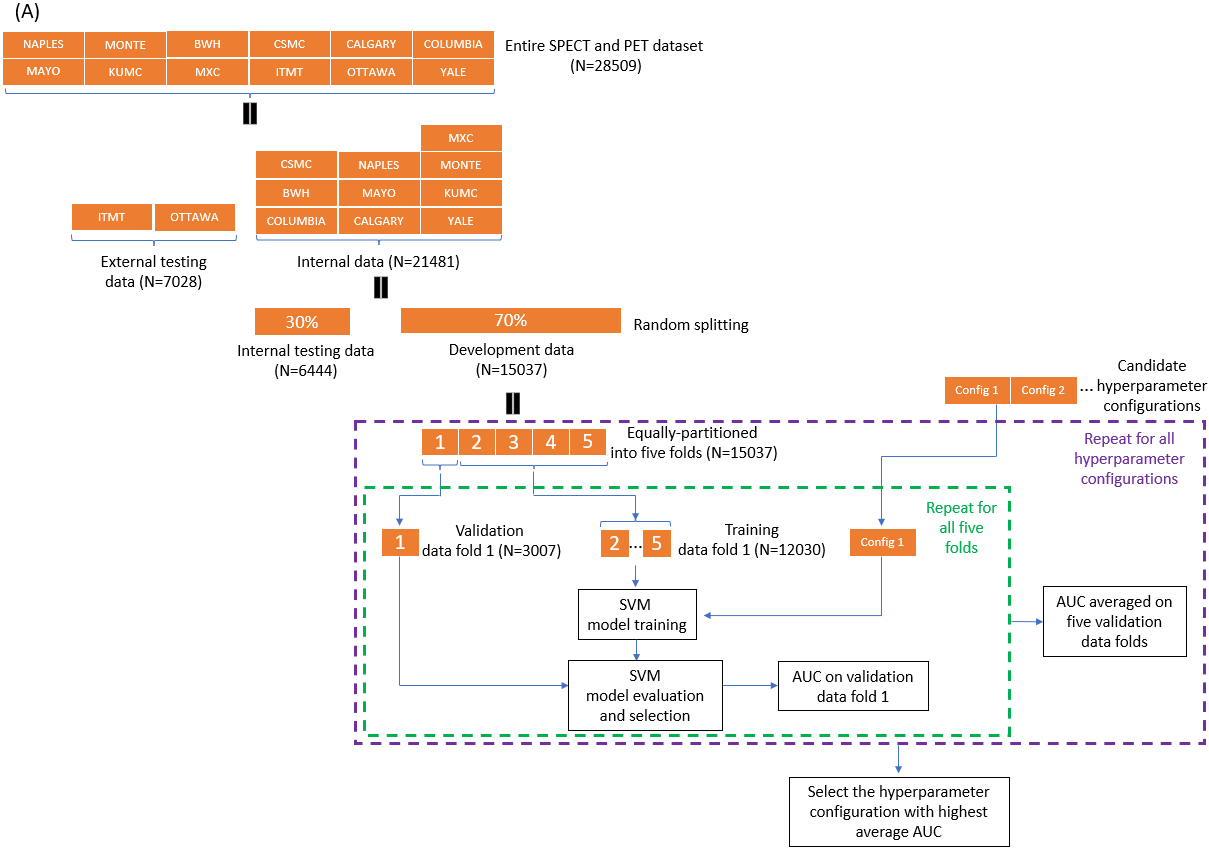

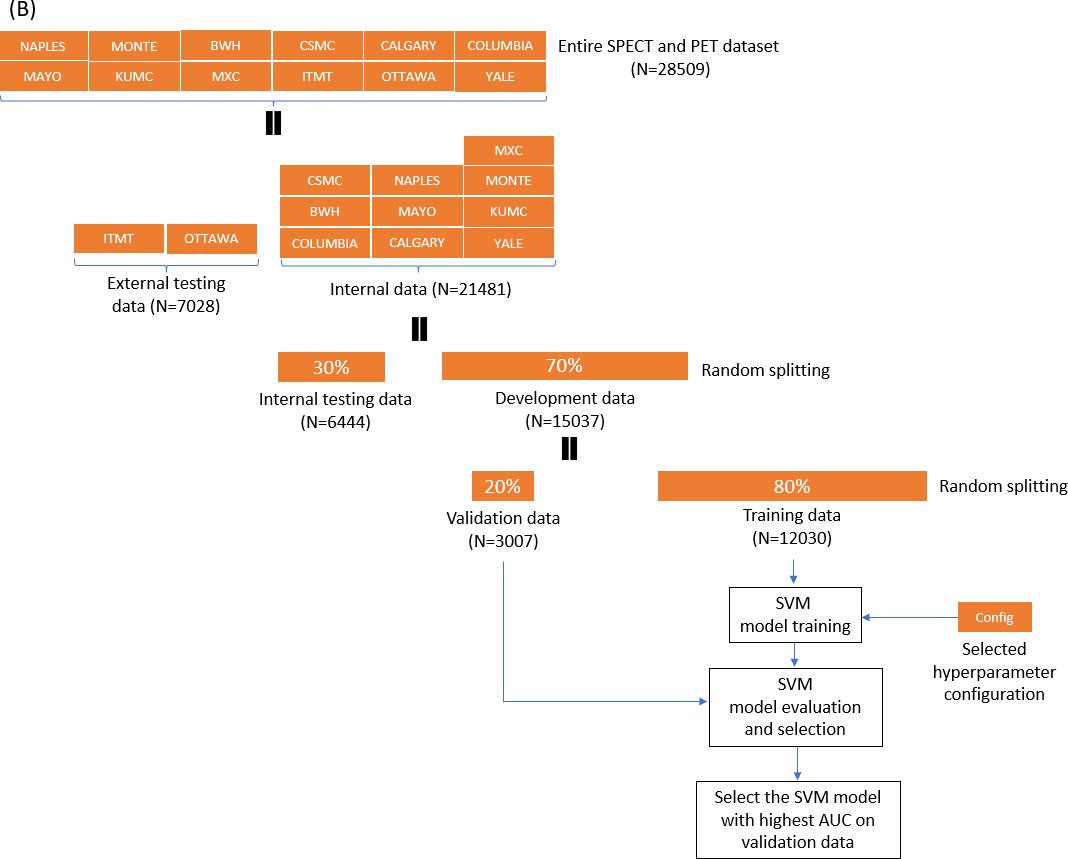

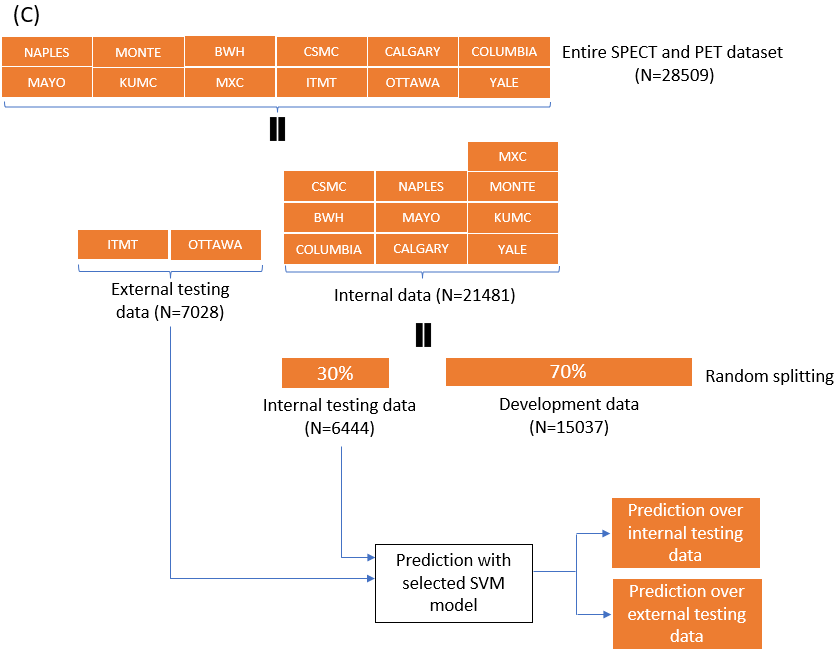

**Supplementary Figure 3. Body composition index (BCI) achieved consistent performance in patient subgroups in external testing cohort.** (A): AUCs for death event prediction at different time points; (B) ROCs for death event prediction during the entire follow-up time window; (C) calibration curves; (D) decision curves.

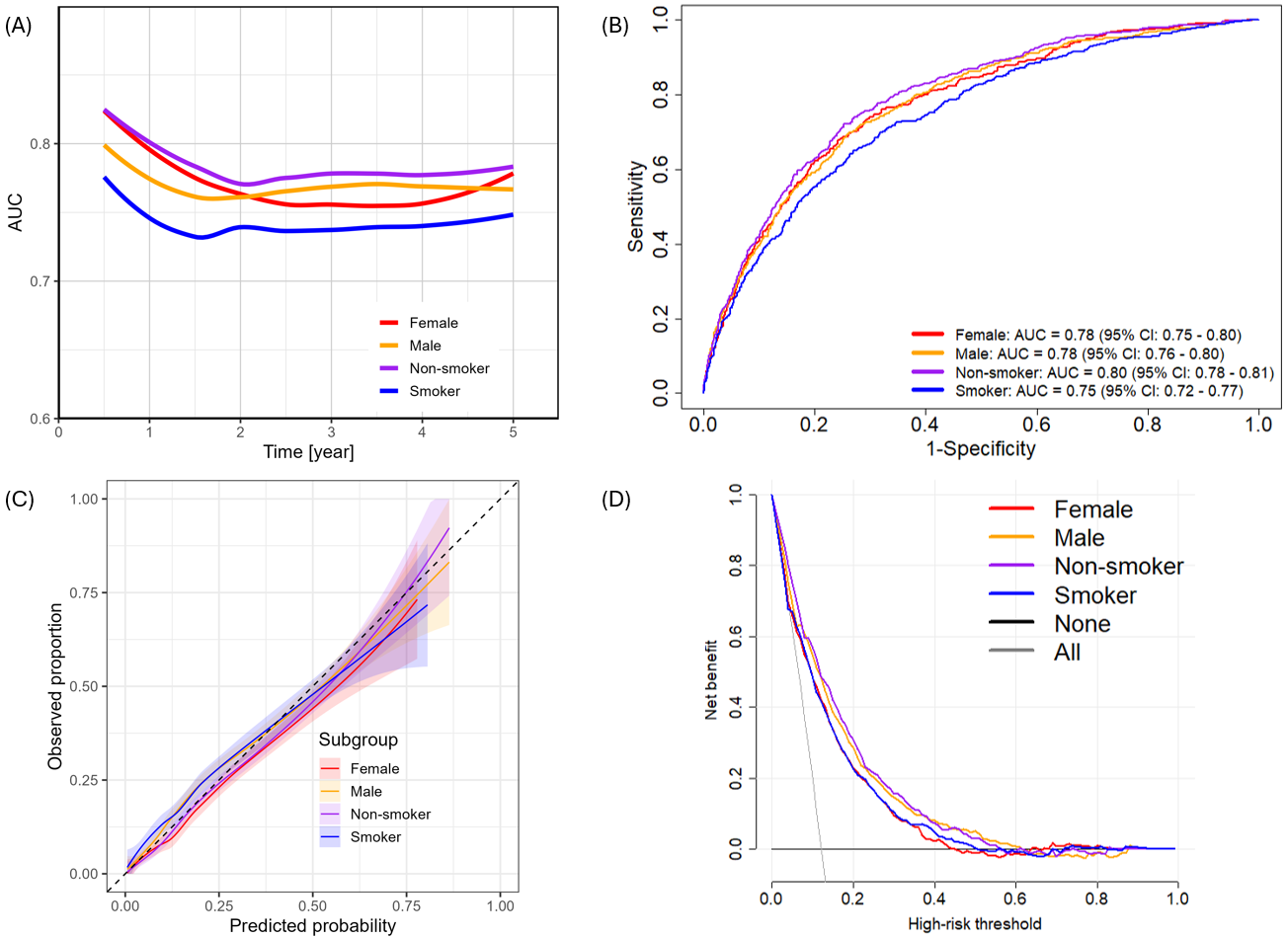

The BCI took body composition metrics, demographical variables, and coronary artery calcium (CAC) score as input. AUC – area under receiver operating characteristic (ROC) curve, Demo – demographical variables.

**Supplementary Figure 4: Feature importance of body composition index (BCI) in different risk groups stratified by body composition index (BCI).** (A) high-risk group. (B): intermediate-risk group; (C): low-risk group.

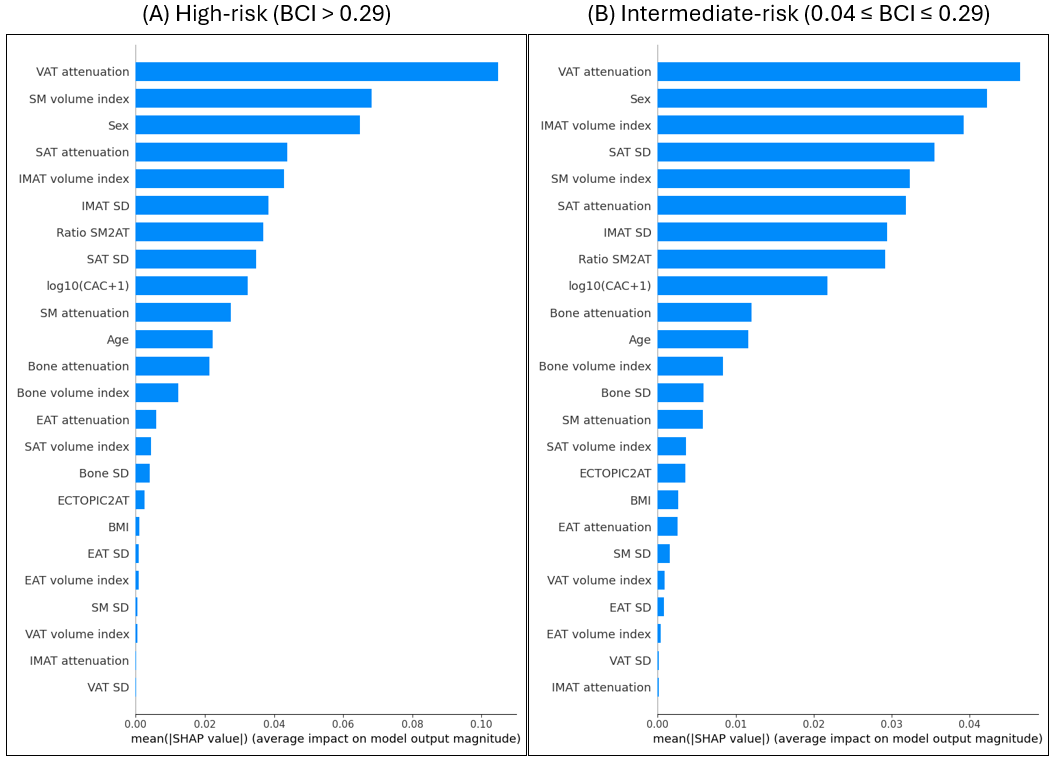

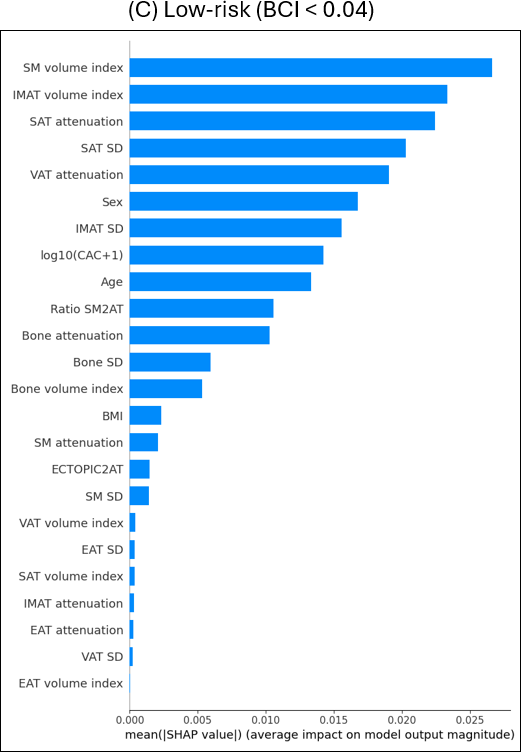

Body composition index (BCI) integrated body composition metrics, coronary artery calcium score, and demographics. VAT  – visceral adipose tissue, EAT – epicardial adipose tissue, SM – skeletal muscle, IMAT – intramuscular adipose tissue, SAT – subcutaneous adipose tissue, ratio SM2AT – ratio of skeletal muscle volume to all adipose tissue volume, ratio ECTOPIC2AT  – ratio of ectopic adipose tissue volume (including EAT, VAT, and IMAT) to all adipose tissue volume, CAC – coronary artery calcium, CAD – coronary artery disease, LVEF – left ventricular ejection fraction, BMI – body mass index, SD – standard deviation.

**Supplementary Figure 5. Body composition index (BCI) achieved better calibration than baseline risk prediction model in external testing cohort.** (A) baseline risk prediction model integrating CT-derived coronary artery calcium (CAC) score and demographical variables; (B) BCI integrating only body composition metrics; (C) BCI integrating body composition metrics, CT-derived CAC score, and demographical variables.

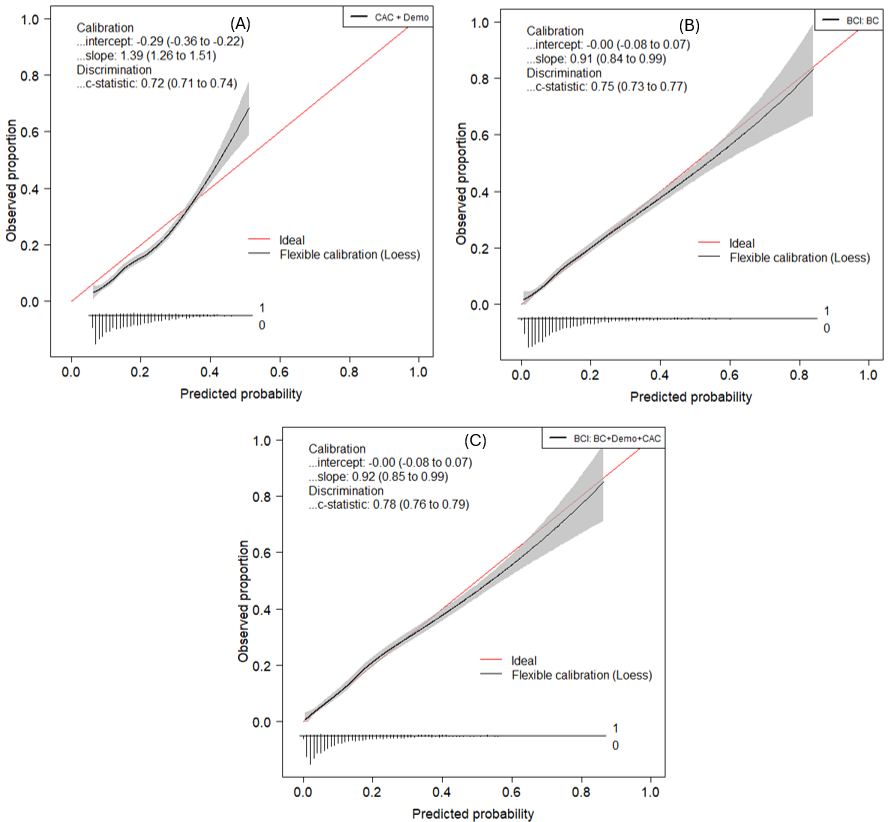
 The baseline model took demographic (Demo) variables (including sex, age, and body mass index) and CT-derived coronary artery calcium (CAC) score as input. Two body composition index models were considered, with one taking only body composition (BC) metrics as input and the other taking BC metrics, Demo variables, and CAC score as input.

**Supplementary Figure 6: Kaplan-Meier curve of body composition index for all-cause mortality risk stratification in the external testing cohort and its subpopulations.** (A): in entire external testing cohort; (B): in female subpopulation; (C): in male subpopulation; (D): in patients with age < 60 years; (E): in patients with 60 ≤ age ≤ 69 years; (F): in patients with 70 years ≤ age.

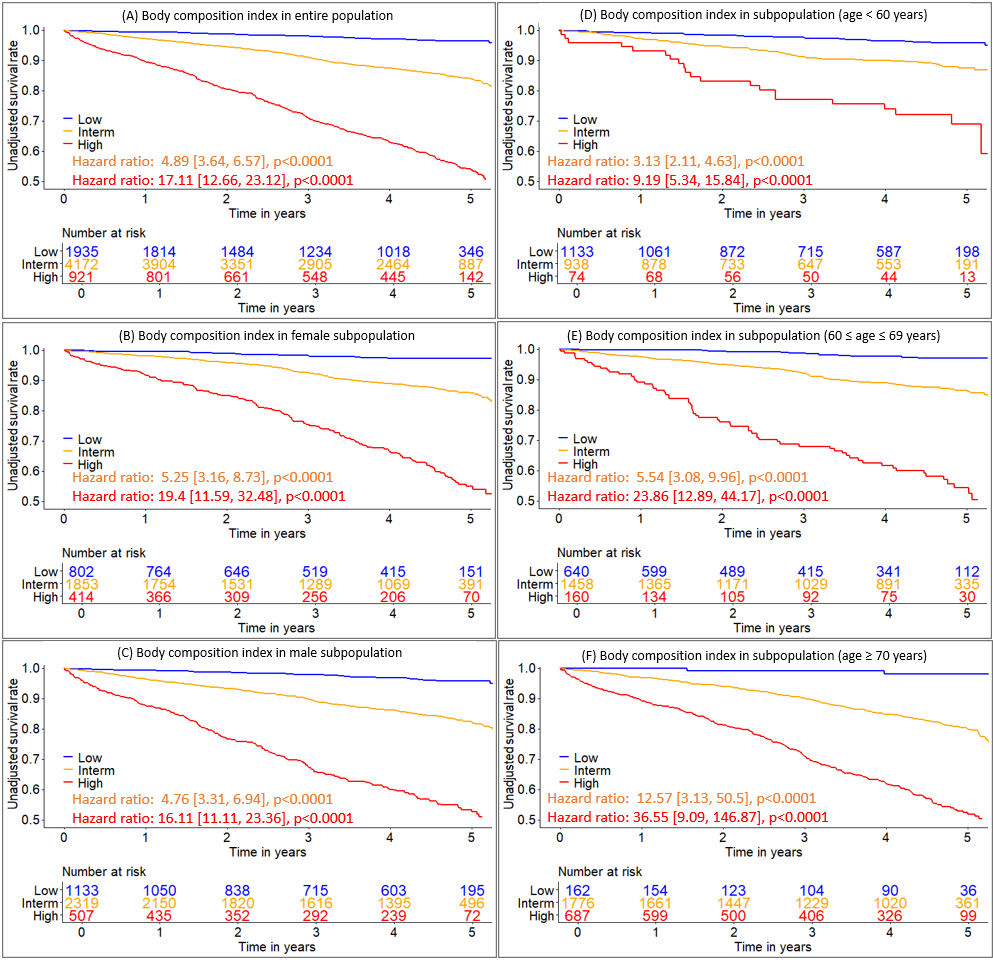

Body composition index integrated body composition metrics, coronary artery calcium score, and demographics.

**Supplementary Figure 7. Distributions of body composition index (BCI) risk groups and their corresponding death rate in external testing subpopulations.**

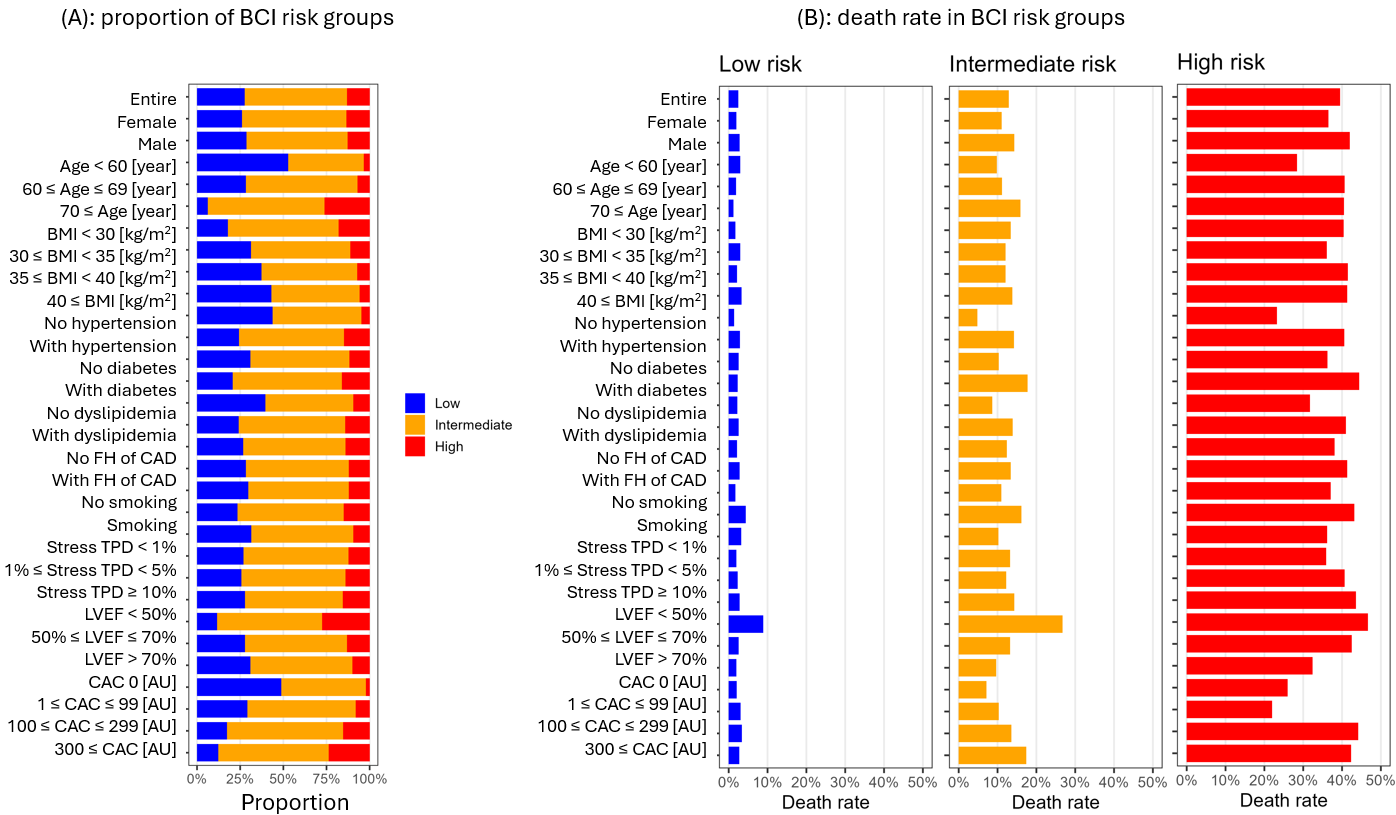

BCI integrated body composition metrics, coronary artery calcium (CAC) score, and demographics. Bold indicates statistical significance. Subpopulations stratification by body mass index (BMI), age, stress total perfusion deficit (TPD), left ventricular ejection fraction (LVEF), and coronary artery calcium (CAC) score follows existing literature.^11-16^ The patients with low body composition index value was reference group. FH – family history, CAD – coronary artery disease, AU – Agatston unit

**Supplementary Figure 8. Body composition index achieved better net clinical benefits than baseline risk index model.**

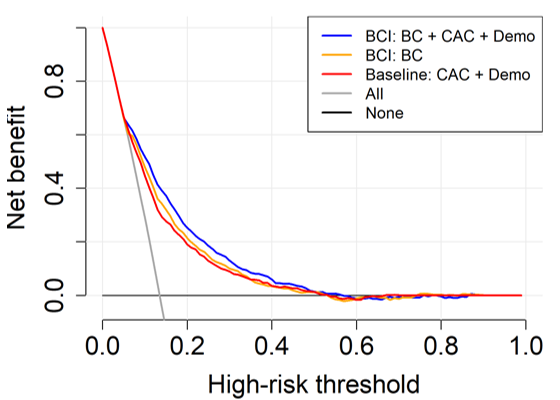

The baseline model took demographic (Demo) variables (including sex, age, and body mass index) and CT-derived coronary artery calcium (CAC) score as input. Two body composition index models were considered, with one taking only body composition (BC) metrics as input and the other taking BC metrics, Demo variables, and CAC score as input.

**Supplementary Figure 9: Model-based simulation of estimated mortality risk reduction through targeted improvement in visceral adipose tissue attenuation across patient subgroups.** (A): in female non-smokers stratified by hypertension (HTN), diabetes mellitus (DM), and dyslipidemia (DLD); (B): in male non-smokers stratified by HTN, DM, and DLD; (C): in female smokers stratified by HTN, DM, and DLD; (D): in male smokers stratified by HTN, DM, and DLD**.**

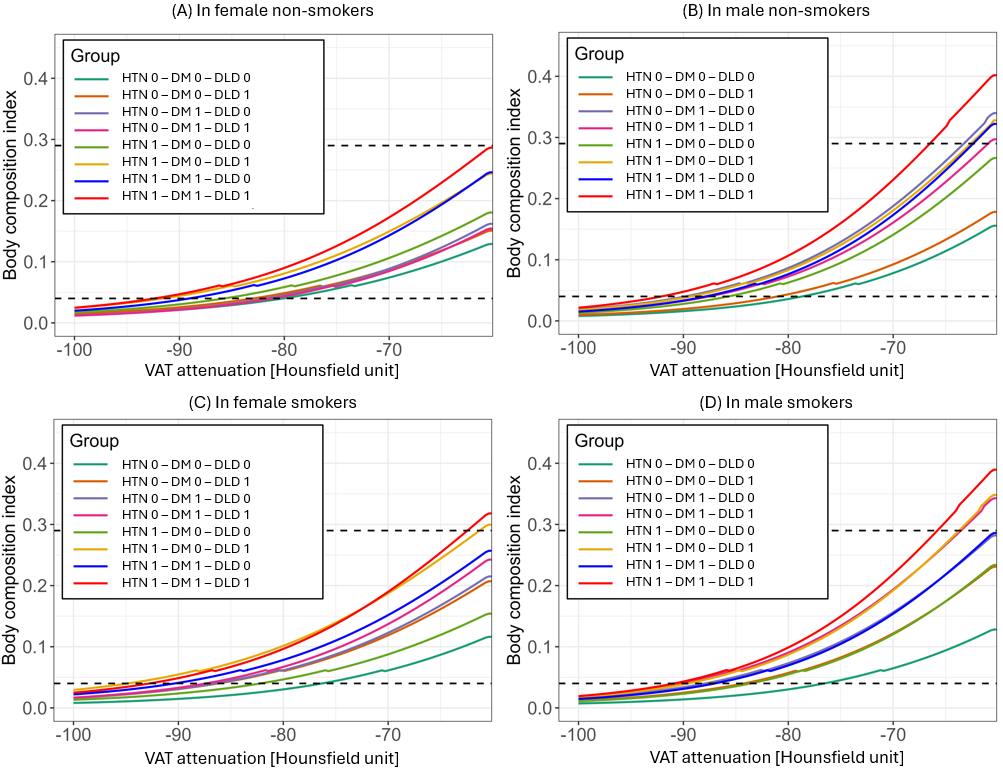

BCI integrated body composition metrics, coronary artery calcium (CAC) score, and demographics. HTN 0 indicated patients without hypertension while HTN 1 indicated patients with hypertension. Similarly, DM and DLD were used to indicate whether diabetes and dyslipidemia were present or not, respectively. Body composition index (BCI) was estimated as a function of simulated decreases in visceral adipose tissue (VAT) attenuation, stratified by sex, smoking status, and cardiometabolic comorbidities. All other body composition metrics were held at subgroup means. Two horizontal dashed lines indicate BCI thresholds of 0.04 and 0.29, corresponding to the low-risk (BCI <0.04), intermediate-risk (0.04 ≤ BCI ≤ 0.29), and high-risk (BCI >0.29) categories. This is a model-based simulation; results should be interpreted as hypothesis-generating and do not imply causal therapeutic efficacy. HTN — hypertension; DM — diabetes mellitus; DLD — dyslipidemia; BCI — body composition index; CAC — coronary artery calcium.

**Supplementary Figure 10: Increasing skeletal muscle (SM) volume index can significantly reduce the body composition index (BCI) in external testing cohort. (A): within female non-smokers; (B): within male non-smokers; (C): within female smokers; (D): within male smokers.**

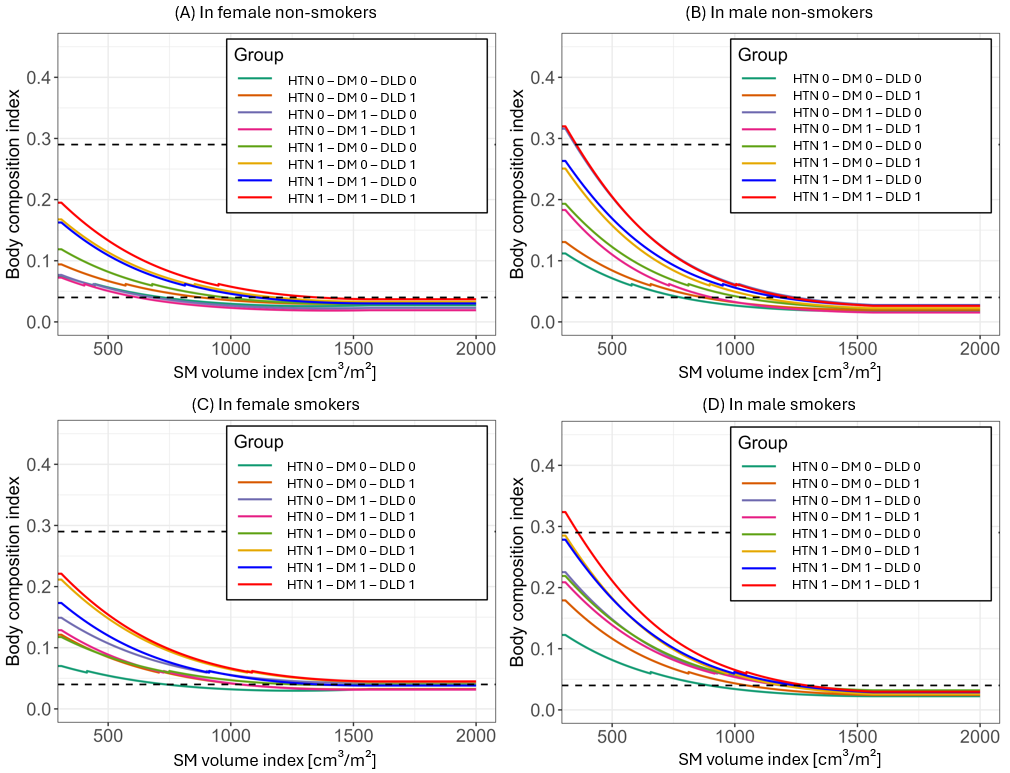

BCI integrated body composition metrics, coronary artery calcium (CAC) score, and demographics. Two dashed horizontal lines corresponded to cutoffs 0.04 and 0.29 which were used to define low-risk (BCI < 0.04), intermediate-risk (0.04 ≤ BCI ≤ 0.29), and high-risk (BCI > 0.29) groups. HTN 0 indicated patients without hypertension while HTN 1 indicated patients with hypertension. Similarly, DM and DLD were used to indicate whether the diabetes and the dyslipidemia were present or not, respectively.

**Supplementary Figure 11: Modifying body mass index (BMI) cannot significantly reduce the body composition index (BCI) and the mortality risk. (A): within female non-smokers; (B): within male non-smokers; (C): within female smokers; (D): within male smokers.**

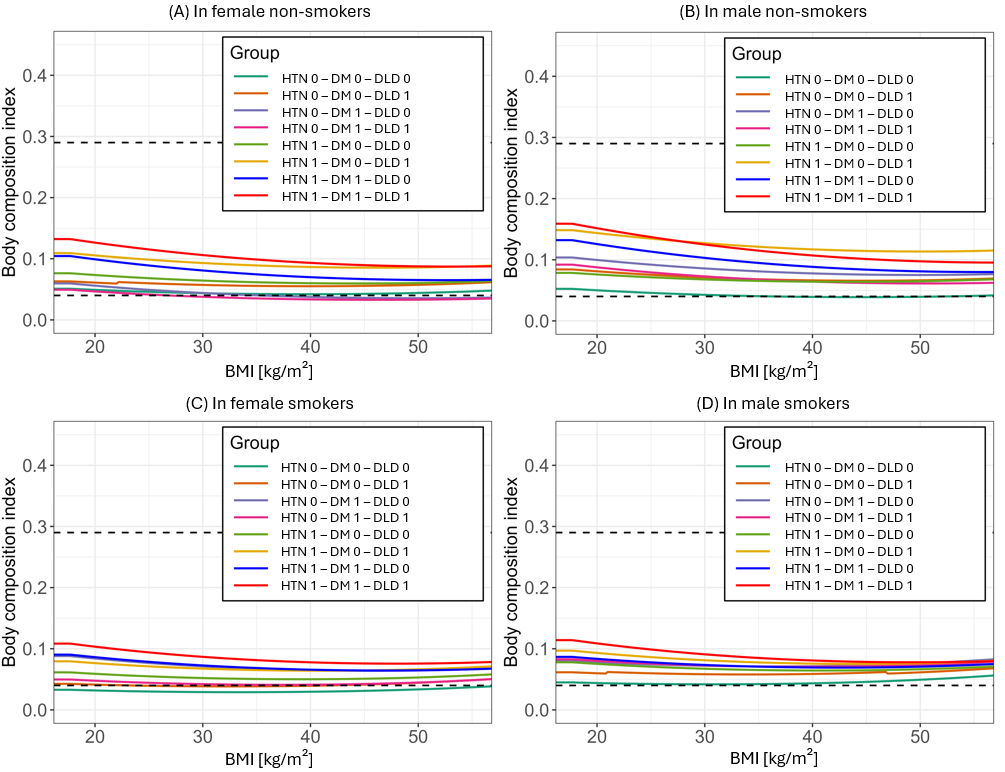
 BCI integrated body composition metrics, coronary artery calcium (CAC) score, and demographics. Two dashed horizontal lines corresponded to cutoffs 0.04 and 0.29 which were used to define low-risk (BCI < 0.04), intermediate-risk (0.04 ≤ BCI ≤ 0.29), and high-risk (BCI > 0.29) groups. HTN 0 indicated patients without hypertension while HTN 1 indicated patients with hypertension. Similarly, DM and DLD were used to indicate whether the diabetes and the dyslipidemia were present or not, respectively.

**Supplementary Figure 12. Comparison of area under receiver operating characteristic (ROC) curve (AUC) for mortality prediction using body composition index (BCI,** which integrated body composition metrics, CT-based coronary artery calcium (CAC) score, and demographical variables) in development, internal, and external testing cohorts.

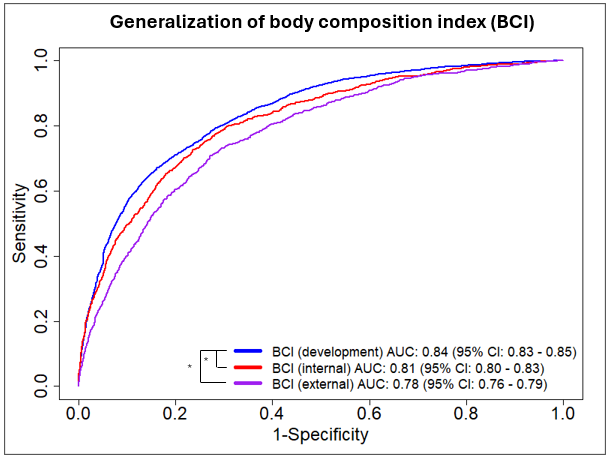

* p<0.05. NS – not statistically significant. BC – 20 body composition metrics, Demo – three demographic variables, CAC – CT-based coronary artery calcium score, Med – five medical histories variables, MPI – three myocardial perfusion variables.

**Supplementary Figure 13. Kaplan-Meier curve of body composition index (BCI) for mortality risk stratification in the entire internal testing data and its different subpopulations.** (A): in entire internal testing cohort; (B): in Black subpopulation of internal testing cohort; (C): in White subpopulation of internal testing cohort; (D): in internal testing cohort patients with age < 60 years; (E): in internal testing cohort patients with 60 ≤ age ≤ 69 years; (F): in internal testing cohort patients with 70 years ≤ age.

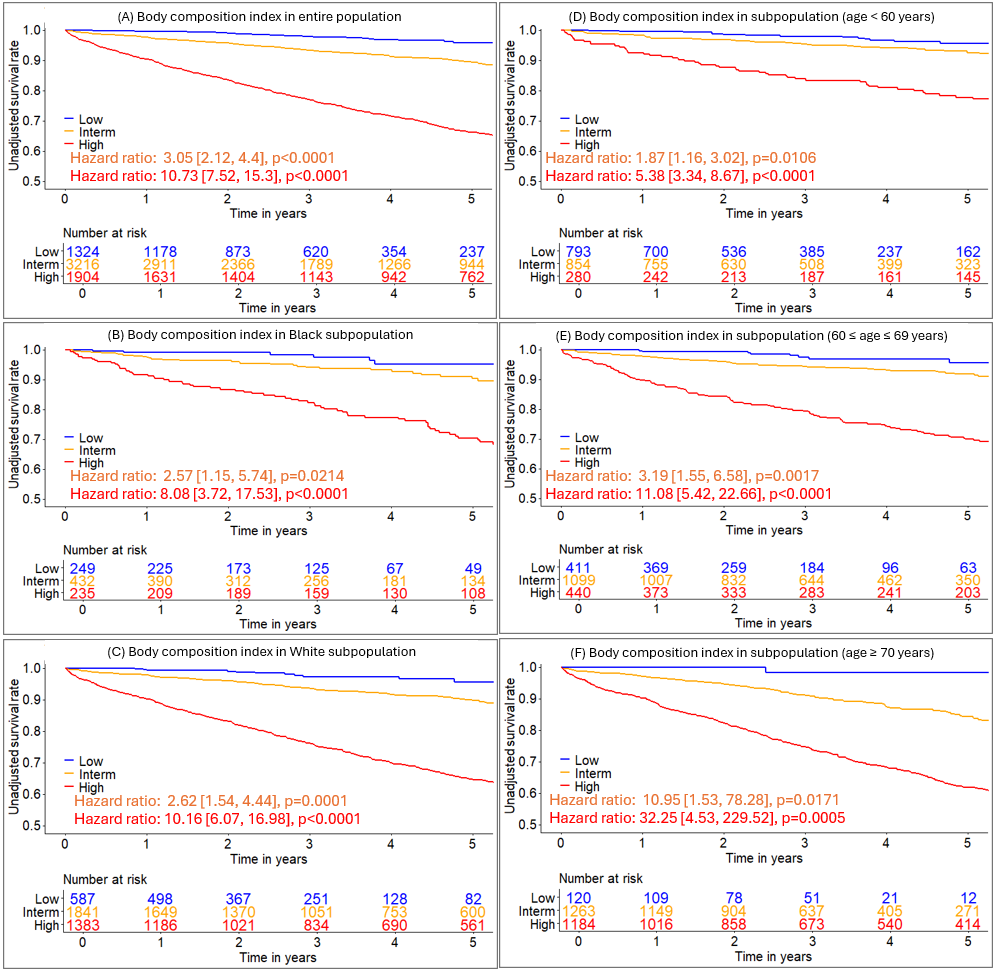

BCI integrating body composition metric, coronary artery calcium score, and demographics
